# Effects of cognitive-behavioural therapy for insomnia on cognitive and emotional functioning in adults with insomnia: behavioural and neuroimaging evidence

**DOI:** 10.64898/2026.10.06.26364880

**Authors:** Kirsten Gong, Emma-Maria Phillips, Aurore A. Perrault, Florence Borgetto, Nyissa A. Walsh, Mathilde Reyt, Antonia Maltezos, James Garneau, Lukia Tarelli, Jennifer J. McGrath, Josée Savard, Sophie Schwartz, Jean-Philippe Gouin, Thien Thanh Dang-Vu

**Author notes:** Corresponding authors, Concordia University’s School of Health, 7200 Sherbrooke Street West, H4B 1R6, Montreal, QC, Canada, Fax: N/A.

## Abstract

**Study Objectives:** To characterize neural correlates of memory and emotion reactivity in individuals with insomnia disorder (INS) compared to good sleepers (GS) and to investigate associated short- and long-term changes following cognitive-behavioural therapy for insomnia (CBTi).

**Methods:** This randomized controlled trial included 56 INS and 20 GS. At baseline, participants underwent resting-state and task-based functional magnetic resonance imaging, including working memory, emotion reactivity, and face-name associative memory tasks. They filled out sleep diaries and self-report questionnaires about sleep, cognition, and emotions, and underwent in-lab polysomnography. Subsequently, participants with INS were randomized to either the 3-month treatment group (TX) or wait-list control group (WL). Both TX and WL groups repeated baseline assessments at three months post-randomization and completed sleep diaries and questionnaires one year after finishing CBTi.

**Results:** At baseline, the INS group reported higher insomnia severity, greater subjective cognitive impairment, and perceived sleep-related stimuli as more negative than GS. Contrasting with these subjective evaluations, no group differences were found in memory performance, task-based brain activity, or resting-state functional connectivity. At three months post-randomization and one-year follow-up, CBTi significantly improved subjective, but not objective, cognitive functioning and sleep in adults with INS. No significant treatment-related changes were observed in valence ratings of sleep images, task-based brain activity, or resting-state functional connectivity.

**Conclusions:** Beyond its primary role in improving insomnia symptoms, CBTi may have long-term benefits for individuals with INS who experience concerns about their cognitive functioning, even in the absence of objective changes in neural processing of cognitive and emotional tasks.

**Clinical trial:** Impact of Insomnia Treatment on Brain Responses During Resting-state and Cognitive Tasks. ClinicalTrials.gov identifier: NCT04024787; https://clinicaltrials.gov/study/NCT04024787

**Statement of Significance:** Insomnia disorder impairs sleep, cognitive, and emotional functioning, imposing a significant clinical burden and highlighting the need for effective interventions to address these impairments. Our study used self-reports, task performance, and neuroimaging techniques to comprehensively investigate the effectiveness of cognitive-behavioural therapy for insomnia on subjective and objective measures of sleep, cognitive, and emotional functions in adults with insomnia disorder. Our findings show that this treatment has long-lasting benefits on subjective, but not objective, cognitive functioning and sleep measures. No changes were found in valence ratings of sleep images, memory performance, or brain function. Future research should identify neural substrates and psychological processes underlying these subjective cognitive deficits as well as the dissociation between subjective and objective cognitive functioning in insomnia.

## INTRODUCTION

Insomnia disorder is a debilitating condition impacting around 16.2% of adults and is characterized by complaints about difficulty falling asleep, staying asleep, and/or undesired earlier awakenings.^1–3^ These symptoms occur at least three times per week, persist for at least three months, and are generally accompanied by daytime impairments in memory, attention, and/or mood.^1,2^ Cognitive-behavioural therapy for insomnia (CBTi) is the first line treatment for this disorder, combining both cognitive and behavioural components to modify individuals’ dysfunctional beliefs about sleep and maladaptive habits that perpetuate insomnia.^4,5^ While CBTi reliably improves insomnia severity and self-reported sleep measures,^6–10^ recent meta-analyses have reported inconsistent effects on objective sleep measures from actigraphy and/or polysomnography (PSG).^6,11–13^

In terms of cognitive functioning, individuals with insomnia disorder commonly report having memory difficulties as well as trouble processing and regulating their emotions.^14–20^ This has been shown by neuropsychological tests in which they tend to be less accurate and make more errors on working memory tasks, yet their reaction times are comparable to good sleepers.^20–24^ Although these individuals also show reduced overnight consolidation of declarative memory relative to good sleepers on a word-pair associates task,^25^ performance on other ecologically relevant tasks, such as memorizing face-name pairs, remains largely unexamined in this population. As for emotional processing, individuals with insomnia disorder have been shown to be slower and less accurate at recognizing facial emotions than good sleepers.^26^

These subjective and objective difficulties in memory and emotional processing are also evident at the neural level. Functional magnetic resonance imaging (fMRI) studies on working memory in insomnia reveal a complex pattern of both hyper- and hypo-activation across different brain regions.^27–33^ In contrast, emotional processing, particularly of sleep-related stimuli that can carry heightened threat or anxiety for individuals with insomnia disorder, is more consistently characterized by hyper-activation in brain regions associated with negative emotions, such as the amygdala and insula.^17,34–37^ Importantly, altered communication both within and between multiple resting-state networks, including the Default Mode, Salience, and Frontoparietal networks, could be linked to poorer self-reported sleep quality, prolonged sleep onset latency, and greater severity of both insomnia and depressive symptoms.^38–48^ Together, these findings highlight that insomnia affects memory and emotional processing at both the behavioural and brain levels, which naturally leads to the question of whether insomnia treatment may restore these functions.

Given the cognitive and emotional toll of insomnia, recent studies have investigated the extent to which CBTi ameliorates these deficits across subjective, objective, or neural indices. While current evidence indicates small-to-moderate improvements in subjective cognitive functioning, these gains do not consistently translate into significant improvements in objective cognitive performance.^7–8,49–50^ Furthermore, despite prevalent memory complaints in this population,^22,27^ changes in memory circuits following CBTi have yet to be explored via fMRI. One fMRI study previously found hypo-activation in medial and inferior prefrontal regions, but during verbal fluency tasks, in a sample of older individuals with insomnia disorder (*Mage* = 61±6.2 years) compared to age-, sex-, and education-matched good sleepers (*Mage* = 60±8.2 years). These neural deficits in individuals with insomnia disorder were partly resolved only in the inferior prefrontal region following sleep therapy that consisted of CBT, body temperature and bright light interventions, sleep hygiene, and physical activity counseling (compared to a wait-list period).^51^ Conversely, heightened limbic activity in response to sleep stimuli has been shown to attenuate following CBTi. However, this study lacked a wait-list control group, emphasizing the need to validate these results in more controlled trials.^52,53^

While the effect of CBTi on sleep has been studied extensively, our understanding of its impact on the brain at rest is still relatively novel. Most intervention studies investigating changes in resting-state networks among individuals with insomnia disorder has commonly used transcranial magnetic stimulation (TMS) instead of psychological interventions.^48,54–55^ To date, the only study evaluating the effect of CBTi on resting-state connectivity focused on specific regions of interest rather than on brain networks and reported decreased functional connectivity between the amygdala and lingual gyrus following CBTi.^28^ This result was interpreted as a decoupling of emotional and visual-processing circuits, potentially reducing hyperarousal. Yet, it is still unclear whether these neural changes also translate into improvements in how individuals with insomnia disorder perceive sleep-related emotional stimuli.

To our knowledge, no study has simultaneously assessed the effects of CBTi on both subjective and objective measures of memory and emotion reactivity using fMRI in adults with insomnia disorder. Most CBTi studies focus on subjective measures of cognitive or emotional functioning (e.g., self-reported questionnaires) or behavioural measures (e.g., accuracy and reaction time), often overlooking the underlying neural correlates.^7–8,50^ While some studies have examined resting-state functional connectivity, changes in connectivity following CBTi in individuals with insomnia disorder are sparse.^28–29,31,33,39^ Here, we used fMRI within a randomized controlled trial (RCT) primarily to investigate whether there are baseline differences in brain activity at rest or during memory and emotion tasks between individuals with insomnia disorder and good sleepers as well as whether CBTi normalizes these changes relative to a wait-list control group. The secondary aim was to evaluate whether there were baseline differences between these two groups in subjective measures of memory, emotion, and sleep using self-reported questionnaires as well as objective measures of sleep using PSG as well as improvement in these measures following CBTi. The primary outcome measures are brain responses during a resting-state sequence and three MRI tasks assessing working and associative memory performance and valence ratings of emotional images. The secondary outcome measures consist of subjective measures (self-reported questionnaires) of sleep (i.e., sleep diaries), cognition, and emotions, and objective measures of sleep (i.e., PSG recordings). We hypothesized that brain responses associated with memory and emotions as well as memory performance and valence ratings would be altered in participants with insomnia disorder compared to good sleepers at baseline. As a result, we hypothesized a larger recovery in these brain responses, memory performance, and valence ratings among participants with insomnia disorder who received the CBTi compared to the three-month wait period. Overall, these analyses will elucidate the effects of CBTi on both cognitive and emotional functioning, extending the current literature beyond its primary focus on sleep outcomes.

## METHODS

### Participants

This RCT was conducted at the Concordia School of Health in Montreal, Canada and included adults between 25 and 65 years old with insomnia disorder (INS) and good sleepers (GS) with no sleep complaints.

Participants underwent a clinical interview and those who met the diagnostic criteria of insomnia disorder (according to the International Classification of Sleep Disorders [ICSD-3]) were included in the INS group.^56^ Those who did not meet the diagnostic criteria nor reported any sleep complaints were classified as GS. To look at the effect of CBTi, INS participants were randomized into one of two groups: an immediate treatment group (TX) or a wait-list control group (WL).

The exclusion criteria consisted of the following: 1) age (< 25 or > 65 years); 2) presence of or a history of medical conditions: current neurological disorder (e.g., epilepsy with any seizure in the past year, concussion in the past 3 months, multiple sclerosis, Parkinson’s disease), past history of brain lesion (e.g., brain hemorrhage, brain tumor, any condition requiring brain surgery), untreated thyroid disorder, chronic pain syndrome self-reported as interfering with sleep (e.g., migraine, fibromyalgia, rheumatoid arthritis), recent and severe infection in the past three months (e.g., pneumonia, kidney infection), active cancer or remitted cancer with cancer treatment within the last two years, renal insufficiency, major cardiovascular events (e.g., stroke, myocardial infarct, heart failure causing limitation of ordinary physical activity), or interventions (e.g., arterial bypass, angioplasty, pacemaker, major surgery requiring general anesthesia in the past three months); 3) sleep disorders other than insomnia (e.g., sleep apnea with an apnea-hypopnea index > 5/hour, restless legs syndrome with symptoms three days or more per week, periodic limb movements during sleep with index > 15/hour, REM-sleep behavior disorder, narcolepsy and other central disorders of hypersomnolence, sleepwalking more than once per month) based on PSG and/or interview; 4) pregnancy/breastfeeding; 5) night or rotating shifts for more than two weeks in the past three months or expecting to do so during the study period; 6) severe cognitive impairments (diagnosed dementia), diagnosed mental disorder (e.g., bipolar disorder (Type I), schizophrenia, psychosis, major depressive disorder, generalized anxiety disorder, substance use disorder, post-traumatic stress disorder) based on the Mini International Neuropsychiatric Interview (MINI),^57^ or current suicidality; 7) regular alcohol or drug consumption (> 10 glasses of alcohol per week, smoking > 10 cigarettes per day, cannabis more than once per week, or illicit drugs more than once per month); 8) current psychotherapy, past CBTi, and/or current use of medications that can impact sleep (e.g., hypnosedatives, antidepressants, anxiolytics); and 9) contraindications to MRI (e.g., metallic implants, claustrophobia). Any participants who had any cardiac, breathing or sleep-related anomalies detected at the PSG screening were also excluded.

Participants were recruited through flyers, journals, newspapers, websites, radio, Concordia University School of Health advertisements, and participant pools from the Centre de Recherche de l’Institut de Gériatrie de Montréal (CRIUGM). The study was approved by the Comité Central d’Éthique de la Recherche (CCER 19-20-04). All participants signed an informed consent form and were financially compensated for their participation.

### Study protocol

Participant eligibility assessment was done through a phone interview, an in-person semi-structured screening interview, and then an in-lab PSG recording during a habituation night (V0) at the Concordia University School of Health sleep laboratory (see Figure 1 for the study design). Following V0, participants completed sleep diaries every morning for two weeks.

**Figure 1.**
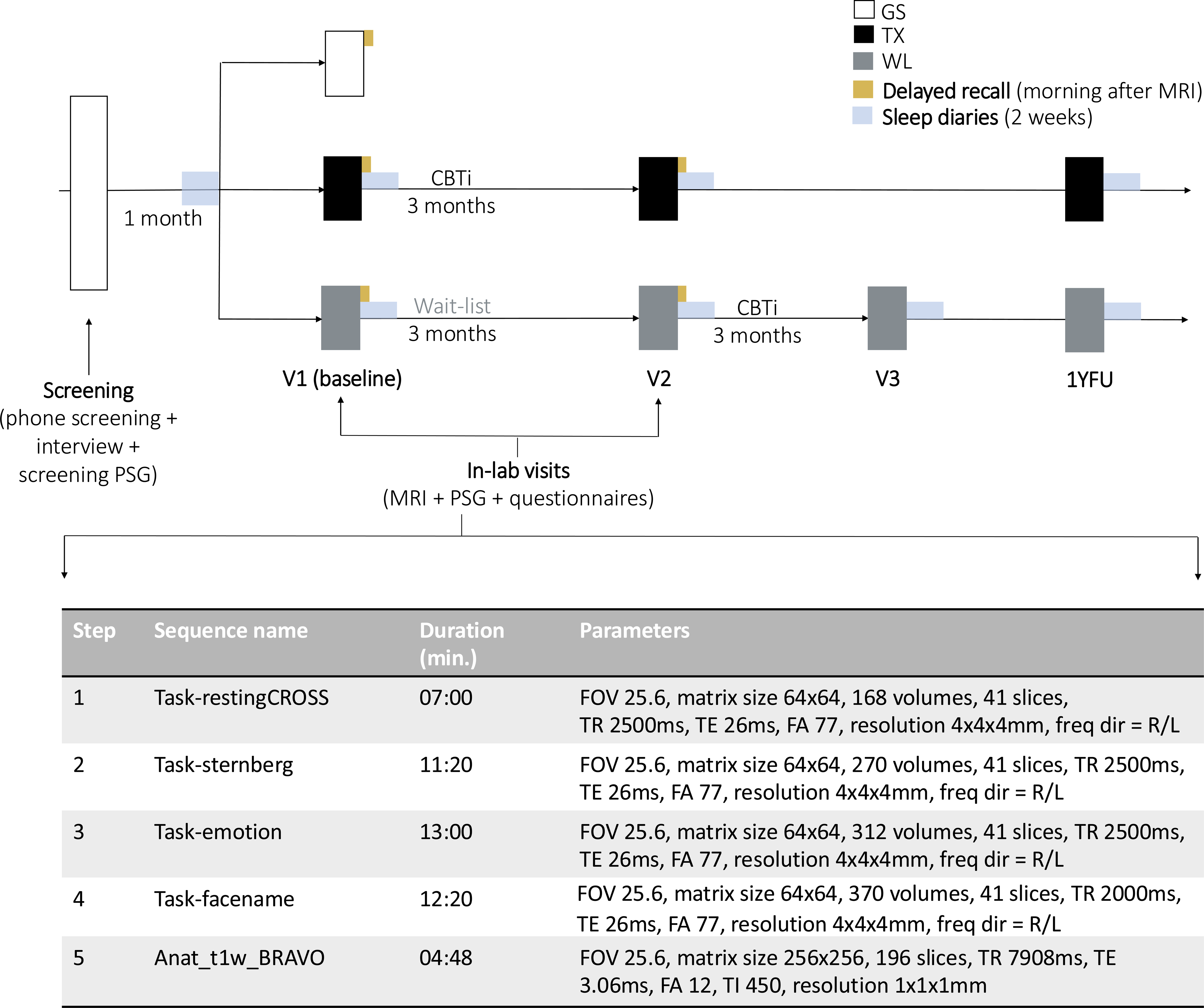
Study design

Eligible participants returned for a second overnight visit (V1) within a month after the initial PSG screening, which consisted of cognitive, neuroimaging, and sleep (PSG) assessments. Upon arrival, participants first performed a practice session outside the MRI consisting of a short version of the resting-state, Sternberg working memory, emotion reactivity, and face-name associative memory tasks that were subsequently performed during the MRI session. The chosen fMRI tasks were selected to target cognitive and emotional domains that are often impaired in insomnia disorder. In between each task, participants were asked to provide their level of sleepiness within the previous five minutes using the Karolinska Sleepiness Scale (KSS).^58^

During the MRI session (∼6PM to 8PM), participants performed the task in a fixed order: a resting-state session followed by one of two versions of the Sternberg working memory,^59^ emotion reactivity,^34^ and then face-name associative memory task.^60–62^ Following this session, participants filled out online clinical questionnaires assessing sleep, emotional arousal, daytime functioning, and cognition and underwent a second PSG sleep assessment. The morning after the MRI session, all participants performed the delayed recall portion of the face-name associative memory task outside of the MRI scanner. To assess CBTi-related changes in sleep measures, the insomnia group completed sleep diaries each morning for two additional weeks.

The morning after the MRI session, participants with insomnia were randomized in a 1:1 ratio into either the TX or WL group. The randomization was stratified according to age (above or below 45), sex, and education level to ensure similar distribution between groups. To maintain allocation concealment, a computer-generated stratified randomization schedule was prepared prior to the start of the study using the permuted block randomization procedure with variable block size (4, 8, 12 participants per block). To also maintain research personnel blind to each participant’s group allocation, sealed envelopes were opened sequentially in the presence of the participant and the research assistant. The TX group started eight 50-minute one-on-one CBTi sessions with a licensed psychologist or a clinical psychology graduate trainee supervised by a licensed psychologist. Over 12 weeks, sessions occurred weekly for four weeks followed by four bi-weekly sessions. These CBTi sessions consisted of psychoeducation about sleep and circadian rhythms, stimulus control, sleep restriction, sleep hygiene, cognitive therapy, and relaxation. The WL group was placed on a 12-week long wait period. After the 12 weeks, participants from both groups returned to the imaging suite for a second MRI session and laboratory within the Concordia University School of Health for their third overnight visit (V2) during which they completed the same assessments as at V1.

After completion of V2, participants in the WL group were then offered the opportunity to access the same CBTi protocol as the TX group. Upon completion of the CBTi, they filled out online self-reported questionnaires at home and filled out sleep diaries every morning for two weeks (V3) without returning to the laboratory for a post-treatment PSG.

All participants with insomnia had a final at-home assessment one year after the end of their respective treatment (one-year follow-up), in which they completed online self-reported questionnaires at home and filled out sleep diaries every morning for two weeks without returning to the laboratory for another neuroimaging or PSG assessment (see Figure 2 for the CONSORT flowchart).

**Figure 2.**
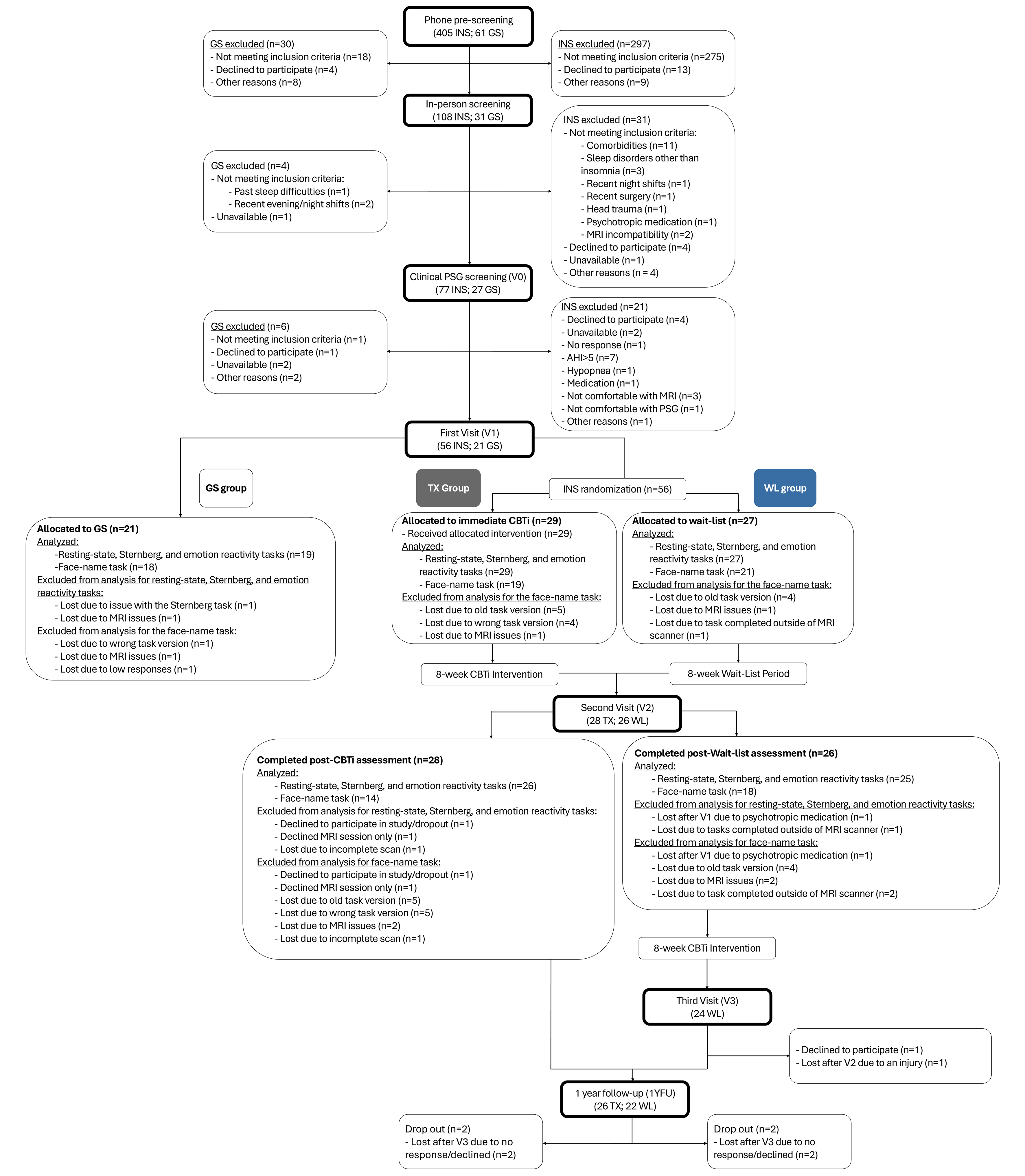
CONSORT flowchart

This study was registered as a clinical trial on ClinicalTrials.gov (NCT number: NCT04024787; https://clinicaltrials.gov/study/NCT04024787).

### Data acquisition and preprocessing

#### MRI data acquisition

MRI data were acquired using a 3-Tesla GE Discovery MR750 scanner with a 32-channel head coil at the Concordia University School of Health imaging suite. Functional scans were acquired using T2*-weighted echo-planar imaging sequences with the following parameters: repetition time (TR) = 2500ms, echo time (TE) = 26ms, flip angle = 77°, field of view = 25.6cm, matrix size = 64×64, 41 axial slices, and voxel resolution 4×4×4mm. The *resting-state* sequence consisted of 168 volumes. The *Sternberg* sequence consisted of 270 volumes and the *emotion reactivity* sequence consisted of 312 volumes, while the *face-name associative memory* sequence used a TR of 2000ms and comprised 370 volumes. High-resolution T1-weighted structural images were obtained using a BRAVO sequence with the following parameters: 196 axial slices, TR = 7908ms, TE = 3.06ms, flip angle = 12°, field of view = 25.6cm, voxel resolution = 1×1×1mm, and matrix size = 256×256 (Figure 1). During all functional scans, the Eyelink 1000 Plus eye tracker (SR Research, Canada) was used to monitor or detect eye closures or signs of sleepiness.

#### Resting-state

Participants completed a 7-minute resting-state scan while fixating on a white fixation cross in the middle of a black screen. They were instructed to avoid closing their eyes or falling asleep. Wakefulness was constantly monitored via real-time eye tracking throughout this task.

#### Sternberg working memory task

Participants memorized sequences of five, six, or seven white digits that were presented one after the other on the black screen (10 trials per difficulty level lasting, on average, 10.5 seconds with pseudo-random inter-stimulus-interval between 2 and 4 seconds). After each sequence, they were given 3.5 seconds to determine if one probe digit was presented during previous sequence of numbers. This was followed by a fixation cross with pseudo-random inter-stimulus-interval between 4 and 8 seconds. At the end of this task, participants rated their perceived performance level, motivation level, and task difficulty on a 10-point Likert scale. The Sternberg working memory task lasted 11 minutes and 20 seconds.^59^ The task design is presented in Supplemental Material (Figure S1A).

#### Emotion reactivity task

The emotion-reactivity task assessed immediate negative emotional valence of 60 images: 20 neutral, 20 negative, and 20 sleep-related images (see Database of emotional images in Supplemental Material).^34,63^ Participants were presented with images (total presentation: 3 seconds; pseudo-random inter-stimulus-interval: between 1 and 3 seconds) and had 5 seconds to rate the extent to which the image evoked a negative emotion (1= not negative to 5 = very negative). A second fixation cross was presented for a pseudo-random duration between 1.5 and 12 seconds before the next image presentation. The emotion-reactivity task lasted 13 minutes. The task design with examples of emotional stimuli is presented in Supplemental Material (Figure S1B).

#### Face-name associative memory task

The face-name associative memory task assessed associative memory between images of individuals’ faces and first names. During the encoding phase of the task that was performed in the MRI, participants were presented with 95 face-name pairs, including 15 control trials. To assess the role of extrinsic motivation, half of the encoded faces were associated with a gold coin/high reward (one dollar), and the other half was associated with a silver coin/low reward (five cents) if memorized correctly. Control faces, which were composed of two repeatedly presented pairs with which the participants had grown familiar during the practice session, were not rewarded. To ensure sustained attention throughout the task, participants were also asked whether they believed the name matched the face (total presentation: 5 seconds; pseudo-random inter-stimulus-interval: between 1.5 and 12 seconds). The face-name associative memory task lasted 12 minutes and 20 seconds.^60–62^

Immediate and delayed (next morning) recalls were performed outside of the scanner. Due to time constraints, the number of stimuli presented during the immediate recall was reduced by half (from 100 to 50) for all participants. Participants were presented with encoded faces (40 at immediate, 80 at delayed) and novel faces (10 at immediate, 20 at delayed). They were asked to select the correct name out of three choices or indicate that the face was new (i.e., not part of the encoding phase). Recall performance corresponded to hits (selecting the correct name), misses (selecting the wrong name or New for an encoded face), correct rejections (correctly selecting New for a novel face; CR), and false alarms (selecting a name for a novel face; FA). The task design is presented in Supplemental Material (Figure S1C).

#### Quality check and pre-processing for task-based fMRI

All anatomical and fMRI data were first converted from DICOM to NIfTI format with dcm2niix (version 1.0.20181125).^64^ Initial visual inspection of task-based fMRI data was performed using FSLeyes version 0.34.2 and Mango version 4.1 software. Task-based data were then preprocessed using Statistical Parametric Mapping 25 (SPM25; version 25.01) within MATLAB (R2020a). The T1-weighted structural images in native space were normalized into standard space using the Montreal Neurological Institute (MNI) 152 template (ICBM 152).^65^ Regarding the functional scans, the first two volumes for each cognitive task were discarded for each participant to provide enough time for the longitudinal magnetization to reach a stable state. Slice timing correction (41 slices; reference slice: 3) was performed to correct for delays in slice acquisition. To correct for any motion, each slice was temporally aligned to the reference slice using a least squares approach as well as spatial transformation using six parameters: rotations along the x, y, and z-axes and left-right, up-down, and forward-backward translations. This was followed by co-registration in which functional images were overlayed with the T1-weighted structural image with the goal of maximizing mutual information. Next, the transformation matrix that was defined for the T1 normalization was applied on the functional images. Lastly, the normalized images underwent spatial smoothing (8mm full width at half maximum [FWHM] Gaussian kernel) to improve the signal-to-noise ratio and to minimize spatial interindividual variability.

#### Quality check and pre-processing for resting-state fMRI

Using the eye tracker, we identified participants who were fighting sleep or fell asleep during the resting-state fMRI session and ran analyses with and without these participants. Nevertheless, all anatomical and resting-state fMRI data were also converted from DICOM to NIfTI format with dcm2niix (version 1.0.20181125).^64^ Initial visual inspection of resting-state fMRI data was then performed using FSLeyes version 0.34.2 and Mango version 4.1 software. Resting-state fMRI data were subsequently preprocessed with fMRIPrep (version 23.2.0) and briefly consisted of bias field correction, skull stripping, slice timing correction, head motion correction, co-registration to the T1-weighted structural image, and normalization.^66^ More details on the exact anatomical and fMRI data pre-processing steps can be found in Supplemental Material.

### Data analyses

#### Statistical power analysis

To determine the sample size needed to detect a change in brain activity during either memory or emotion tasks using fMRI in individuals with insomnia disorder, an a priori power analysis was conducted using G*Power (version 3.1.9.7).^67^ In Kim et al.’s study that included emotional images (*N* = 14),^52^ the effect size for reduced brain activity following CBTi was Cohen’s *d* = 0.61 or Hedges’ *g* = 0.57. Considering a two-tailed alpha level of .05, a power analysis for a repeated-measures effect indicated that a total of 24 participants would be needed to have a statistical power of .80. However, to account for our mixed-factorial design, which includes the TX and WL groups assessed pre- and post-intervention, and to accommodate for any potential attrition, we aimed to include at least 24 participants per group.

### Primary outcomes

#### Task-based fMRI analyses

In terms of the primary behavioural outcomes derived from the fMRI tasks, performance (i.e., percentage of correct answers) was extracted as the main outcome variable of interest for the Sternberg working memory task. The main outcome variable of interest for the emotion reactivity task was the average valence ratings for each image category. Lastly, the main outcome variables of interest for the face-name associative memory task were raw performance, including hits, accuracy ([hit+CR - miss+FA] to measure correct vs. incorrect responses), and recognition (hit-FA) as well as performance change overnight (i.e., delayed vs. immediate recall).

All task-based fMRI analyses were conducted in SPM25 (version 25.01) within MATLAB (R2020a). First-level analyses used a general linear model to assess whole-brain brain activation for each participant during each cognitive task. In the case of the Sternberg working memory task, the condition of interest consisted of correct encoding, pooling all difficulty levels. For the emotion reactivity task, the conditions of interest were neutral, negative, and sleep-related images. Lastly, for the face-name associative memory task, the conditions included hits, misses, and control.

Onsets and duration times were extracted from the PsychoPy files that were generated for each fMRI task (standalone version 3.0.7). The timing of each condition was convolved with the timing of the canonical hemodynamic response function. The six motion parameter estimates were included as nuisance regressors. The main first-level contrasts of interest for the Sternberg, emotion reactivity, and face-name associative memory tasks consisted of (1) correct encoding; (2) sleep-related condition > neutral condition; (3) hit (i.e., correctly memorized pairs) + miss (i.e., incorrectly memorized pairs) > control condition. The contrasts of interest for both memory tasks were selected to maximize statistical power and to keep as many trials as possible.

Second-level analyses were conducted in two steps for each task. First, the main first-level contrasts of interest were compared between the INS and GS groups at V1 using two-sample *t*-tests. Then, to assess the effect of CBTi among participants with insomnia, a factorial design was implemented to determine the impact of Group and Visit factors and their interaction. Age and sex were added as covariates in all analyses. To correct for multiple comparisons, the Benjamini-Hochberg procedure was used with a false discovery rate (FDR) set at .05.

#### Resting-state functional connectivity analyses

The resting-state task was used to assess resting-state networks, mainly the Default Mode Network (DMN), Salience Network (SN), and Frontoparietal Network (FPN). Resting-state functional connectivity analyses were performed using Nilearn (version 0.10.2).^68^ The 100-parcel Schaefer atlas organized into seven resting-state networks was used to independently define the DMN, SN, and FPN.^69,70^ The DMN consisted of the temporal cortex, parietal cortex, prefrontal cortex (ventral, dorsal, and medial), and precuneus-posterior cingulate cortex in both hemispheres. The SN was composed of the left parietal operculum, left frontal-operculum-insula, left prefrontal cortex (lateral), left medial nodes, right temporo-parieto-occipital region, right frontal-operculum-insula, and right medial nodes. Lastly, the FPN included the right posteromedial cortex and bilateral prefrontal cortex, parietal cortex, cingulate cortex, and precuneus.

For each parcel, the blood oxygen level dependent (BOLD) signal was averaged across all voxels within that parcel. Global signal, white matter, framewise displacement, and six motion parameters were included as confound regressors. To assess within-network connectivity, a correlation matrix was generated for each participant by computing the correlation coefficients between each parcel and all other parcels within a given resting-state network. Each correlation coefficient was converted to *z*-scores using the Fisher’s *r*-to-*z* transform. Group-level differences in resting-state functional connectivity within a given resting-state network were assessed using a two-sample *t*-test. To assess the effectiveness of CBTi on resting-state functional connectivity, linear mixed-effects models were performed with Group (TX vs. WL) as the between-subjects factor and Visit (V1 vs. V2) as the within-subjects factor. Age and sex were added as covariates in both types of models. To correct for multiple comparisons, the Benjamini-Hochberg procedure was used with a false discovery rate (FDR) set at .05.

### Secondary outcomes

To evaluate secondary measures of cognition, emotions, and sleep, all participants completed a battery of validated self-report questionnaires at V1. Participants in the TX and WL groups also completed them at V2 and one-year follow-up. Self-reported cognitive functioning was evaluated using the Sahlgrenska Academy Self-reported Cognitive Impairment Questionnaire (SASCI-Q).^71^ Self-reported emotional functioning was assessed using the Beck Depression Inventory (BDI-II),^72^ Positive Affect Negative Affect Scale (PANAS),^73^ and State-Trait Inventory for Cognitive and Somatic Anxiety (STICSA).^74^ Subjective sleep measures were tracked through sleep diaries. Sleep variables of interest, averaged over the two weeks, included subjective total sleep time (TST), sleep onset latency (SOL), wake after sleep onset (WASO), and sleep efficiency (SE)^75,76^ alongside the following questionnaires: Insomnia Severity Index (ISI),^77^ Pittsburgh Sleep Quality Index (PSQI),^78^ the abbreviated version of the Dysfunctional Beliefs and Attitudes about Sleep (DBAS-16),^79^ Daytime Insomnia Symptom Response Scale (DISRS),^80^ and Karolinska Sleepiness Scale (KSS).^58^ The following objective sleep measures were extracted from full-night PSG recordings: TST, SOL, WASO, and SE. Comprehensive details regarding questionnaire subscales, scoring interpretation, internal consistency as assessed with Cronbach’s alpha, or test-retest reliability using intraclass correlation coefficients (ICC) are provided in the Supplemental Material.

### Statistical analyses for demographic, behavioural, and questionnaire data

Demographic, primary behavioural outcomes derived from the fMRI tasks, and self-reported questionnaire data were analyzed using RStudio (2025.09.2+418). Linear models with Group as a fixed effect were used for each outcome derived from these questionnaires to determine if there were differences between the INS and GS groups at V1. Once the models were fitted, the assumption of normality of residuals for each outcome was assessed visually using histograms. The Multivariate difference in fits (MDFFITS) function was also used to identify any influential points for each outcome.^81^ Post-hoc comparisons were assessed with estimated marginal means (emmeans) from the fitted models.^82^ To examine the effect of CBTi on these measures among participants with insomnia, linear mixed-effects models (lmer) were implemented with Group, Visit, and their interaction included as fixed effects. To account for repeated measures, the intercept for each participant was added as a random effect. Age and sex were added as covariates in both types of models.^83^ To correct for multiple comparisons, the Benjamini-Hochberg procedure was also used with an FDR set at .05. Missing data in the dependent variables of interest were handled under the Missing at Random assumption, keeping all available observations per participant in the analyses. Standardized regression coefficients (*β*) were obtained by refitting the models on *z*-scored continuous predictors and outcomes. Effect sizes were interpreted using the following general benchmarks: *β* ≈ 0.10–0.29 = small, 0.30–0.49 = medium, ≥ 0.50 = large.^84^

## RESULTS

### Participant characteristics

In this RCT, 56 adults with insomnia disorder (*Mage* = 43.71±10.93 years, range 25–65 years; 66% female) were randomized to receive CBTi immediately (*n*TX = 29) or to a three-month wait-period (*n*WL = 27; Table 1). Additionally, 20 adults with no sleep complaints were included in the GS group (*M*age = 37.40±11.86 years, range 25–64 years; 50% female). Most of the participants self-identified as White (66%), had a university degree (89%), and were right-handed (89%). Of the 43 women who completed a questionnaire about women’s health, 18 were in perimenopause, with four taking menopause hormone therapy containing both estrogen and progesterone. On average, participants with insomnia disorder were significantly older than good sleepers (*t*(74) = 2.17, *p* = .03, Cohen’s *d* = 0.57). There were no differences in IQ (from Raven’s Progressive Matrices test; 10 items) between INS and GS groups at V1. Although the duration of insomnia varied widely among INS participants, there were no statistically significant differences between TX and WL groups (*t*(50) = 1.43, *p* = .16, Cohen’s *d* = 0.40). The demographic characteristics of the sample at baseline are presented in Table 1. Although there were initially 21 good sleepers, one was excluded from all analyses due to MRI issues. One participant from the TX group and another from the WL group dropped out after V1. Thus, the total attrition rate for this study was 3.9%. The other reasons for excluding participants from analyses for each MRI task at each visit are listed in Figure 2.

**Table 1.**
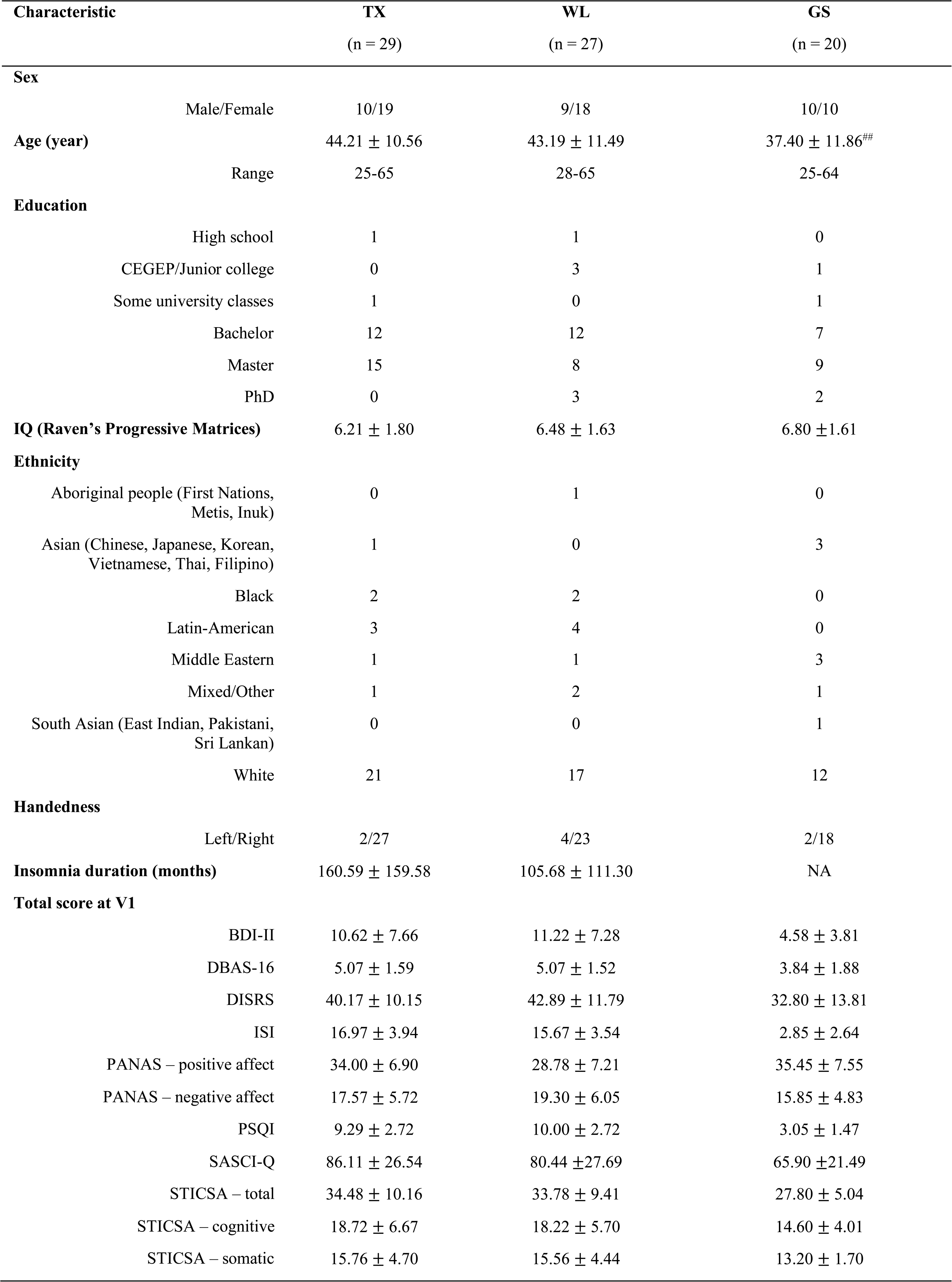
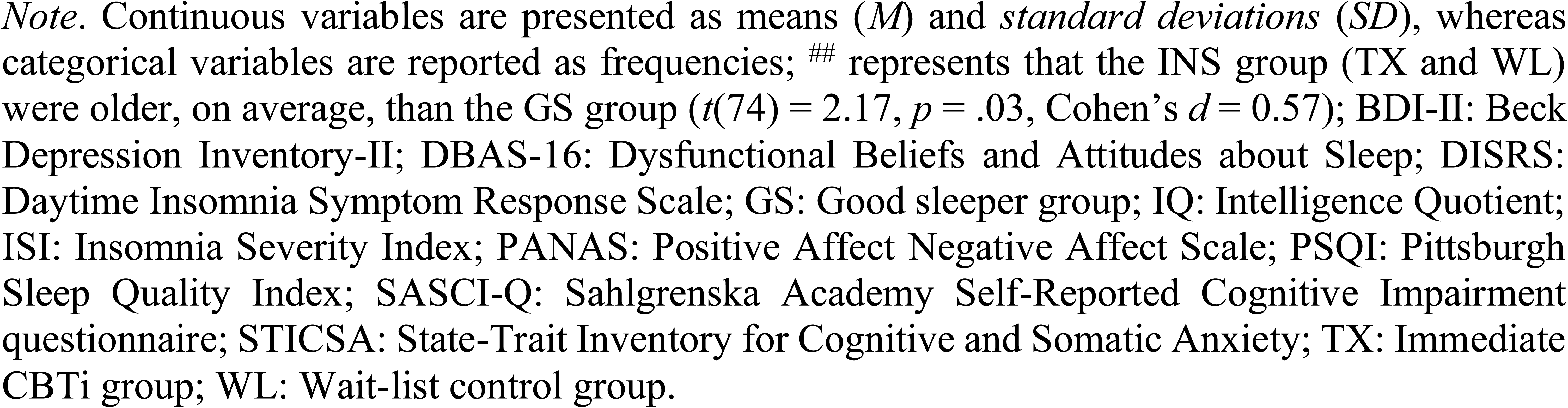
Descriptive statistics of the sample at baseline.

### Comparison between individuals with insomnia and good sleepers

#### Primary outcomes

##### Cognitive performance

At V1, the performance on the Sternberg (i.e., percentage of correct answers) and face-name tasks did not differ between participants with insomnia and good sleepers (all *p* > .05). No statistically significant group differences in Sternberg task performance were found across the five-, six-, or seven-digit difficulty levels (all *p* > .05). Therefore, these levels were pooled together in subsequent analyses. For the emotion reactivity task, the INS group rated the sleep-related images as more negative than GS (*p* = .03). The standardized effect size was large (*β* = .70, 95% CI [0.18, 1.23]). There was no Group effect for neutral or negative images (all *p* > .05; Table S1). Regarding the face-name task, no statistically significant differences in number of hits, accuracy, or recognition were found for both immediate and delayed recall between groups. Similarly, there were no statistically significant differences in the number of hits, accuracy, or recognition at delayed recall compared to immediate recall between the INS and GS groups (all *p* > .05; Table S1). Although group differences did not achieve statistical significance for all cognitive performance measures except for the valence rating of sleep images, the remaining standardized effect sizes were consistently small-to-medium in magnitude (*β* = 0.07 to 0.40). Results regarding lack of group differences in performance for high-vs. low-reward face-name pairs between INS and GS groups can be found in Supplemental Material (Table S2).

##### Brain activity

No statistically significant differences in brain activity between the INS and GS groups at V1 were detected during the Sternberg, emotion reactivity, and face-name tasks (all *p* > .05; Table S3).

Similarly, there were no statistically significant differences in within- and between-network resting-state functional connectivity involving regions within the DMN, SN, or FPN when comparing the two groups at V1 (all *p* > .05; Figures S2–S3). These results did not differ whether we included or excluded four INS and two GS participants who were fighting sleep or fell asleep at any point during the resting-state fMRI session at V1 (Figures S2–S3).

#### Secondary outcomes

##### Self-reported cognition measure

Compared to the GS, the INS group scored higher on the SASCI-Q at baseline, indicating greater self-reported cognitive difficulties (*p* = .02; Table S1).

##### Self-reported emotion measures

In terms of emotional states or traits at baseline, the INS group reported having less positive and more negative affect than the GS group (*p* = .02). The INS group also had higher total BDI-II and STICSA (total, cognitive, and somatic) scores, reflecting greater depressive (*p* = .001) and anxiety symptoms (*p* = .01), respectively (Table S1).

##### Self-reported measures of sleep

The KSS scores before each MRI task between the INS and GS groups at baseline did not differ (all *p* > .05).

At V1, the INS group reported longer SOL, more and longer awakenings during the night, reduced total sleep duration, and poorer SE than good sleepers (*p* < .05). The INS group also had more dysfunctional beliefs and attitudes about sleep (DBAS-16) as well as greater daytime ruminative thoughts when feeling tired (DISRS), worse insomnia severity and sleep quality (ISI and PSQI) than the GS group (*p* < .05; Table S1).

##### Objective measures of sleep

At V1, the INS group exhibited more WASO than the GS group. However, no statistically significant group differences emerged for PSG-derived SOL, TST, and SE (Table S4).

### Effects of CBTi on brain activity, cognition, emotions, and sleep

#### Primary outcomes

##### Cognitive performance

No statistically significant Group*Visit interaction or main effects of Group or Visit were found for performance on the Sternberg task between the TX and WL groups (all *p* > .05; Figure 3B and Table S5). For the emotion reactivity task (Figure 3C and Table S5), no statistically significant Group*Visit interactions or main effect of Group were observed for the mean valence rating of neutral, negative, and sleep-related images (all *p* > .05). There was a statistically significant main effect of Visit on valence rating for neutral and negative images (*p* = .02), with these images being rated, on average, as less negative at three months post-randomization than at V1. Additionally, the main effect of Visit on valence rating for sleep images was marginally significant (*p* = .05). Regarding performance on the delayed portion of the face-name task with 100 stimuli (Figure 3D and Table S5), there was no statistically significant Group*Visit interaction, main effect of Group, or main effect of Visit for number of hits, accuracy, and recognition (all *p* > .05). Results regarding no statistically significant effects of CBTi on performance for the immediate recall (50 stimuli), performance for the delayed recall (50 stimuli), change in performance from delayed to immediate recall (overnight consolidation with 50 stimuli each), and performance for high-vs. low-reward face-name pairs can be found in Supplemental Material (Tables S5–6).

**Figure 3.**
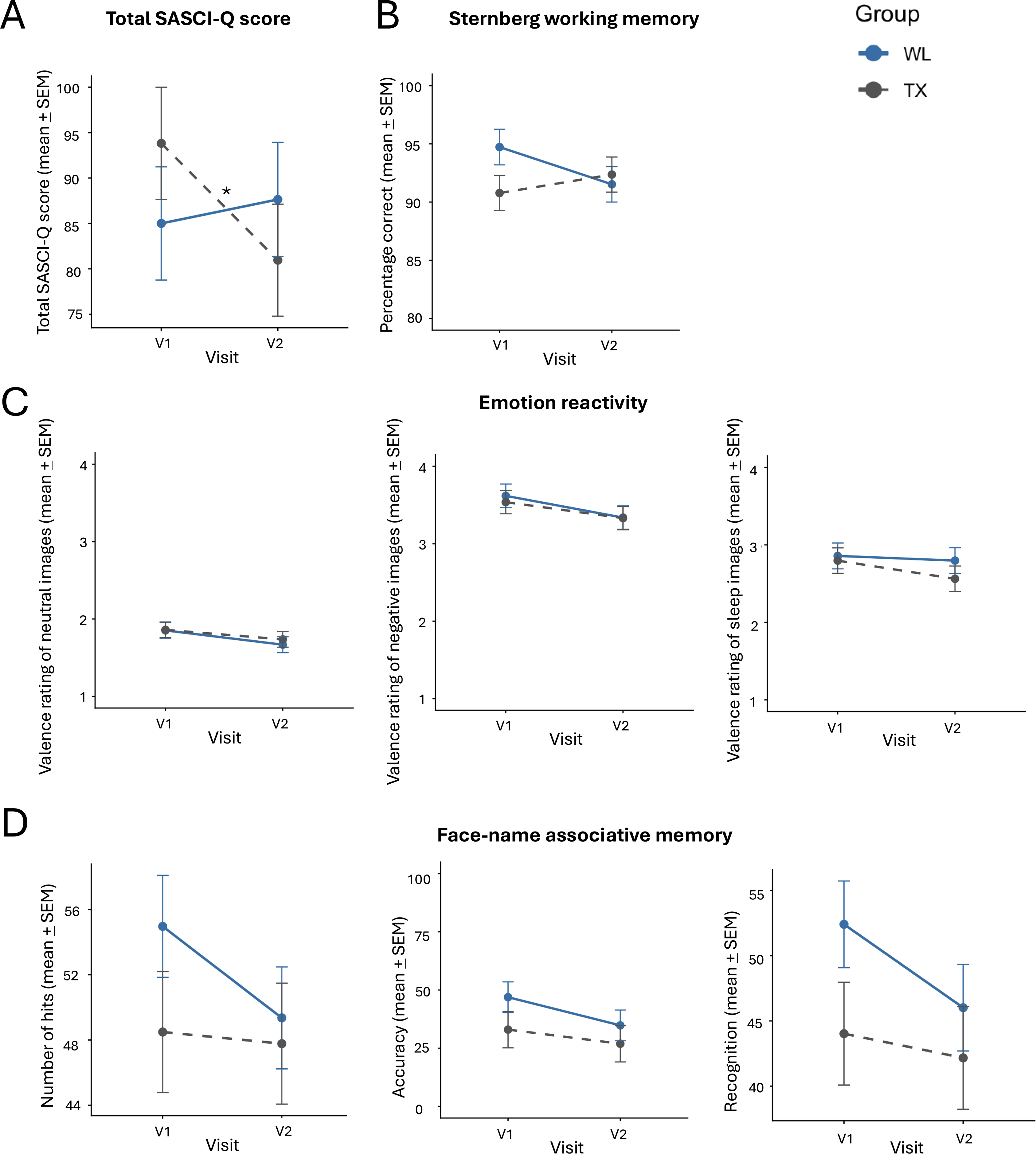
Effects of CBTi on total SASCI-Q score and cognitive performance (A) Total Sahlgrenska Academy Self-reported Cognitive Impairment questionnaire (SASCI-Q) score at baseline (V1) and three months post-randomization (V2) (B) Percentage correct on the Sternberg working memory task at V1 and V2 (C) Valence rating of neutral, negative, and sleep images for the emotion reactivity task at V1 and V2 (D) Number of hits, accuracy [Hit + Correct Rejection (CR) – Miss + False Alarm (FA)], and recognition (Hit – FA) for the face-name associative memory task (delayed recall with 100 stimuli) at V1 and V2 The TX group is shown in grey with a dashed line and the WL group in blue with a solid line. Error bars represent standard error of the mean (SEM). Asterisks represent statistical significance (*p*): * <.05.

##### Brain activity

At the whole-brain level, no statistically significant Group*Visit interaction or main effects of Group or Visit were found for brain activity during all three fMRI tasks. More precisely, analyses revealed no overall group differences or changes from pre-to post-intervention in brain activity while the TX and WL groups correctly encoded the digits during the Sternberg task, viewed sleep vs. neutral images during the emotion reactivity task, or engaged in the face-name task during hit and miss vs. control conditions (all *p* > .05; Figure 4 and Table S3).

**Figure 4.**
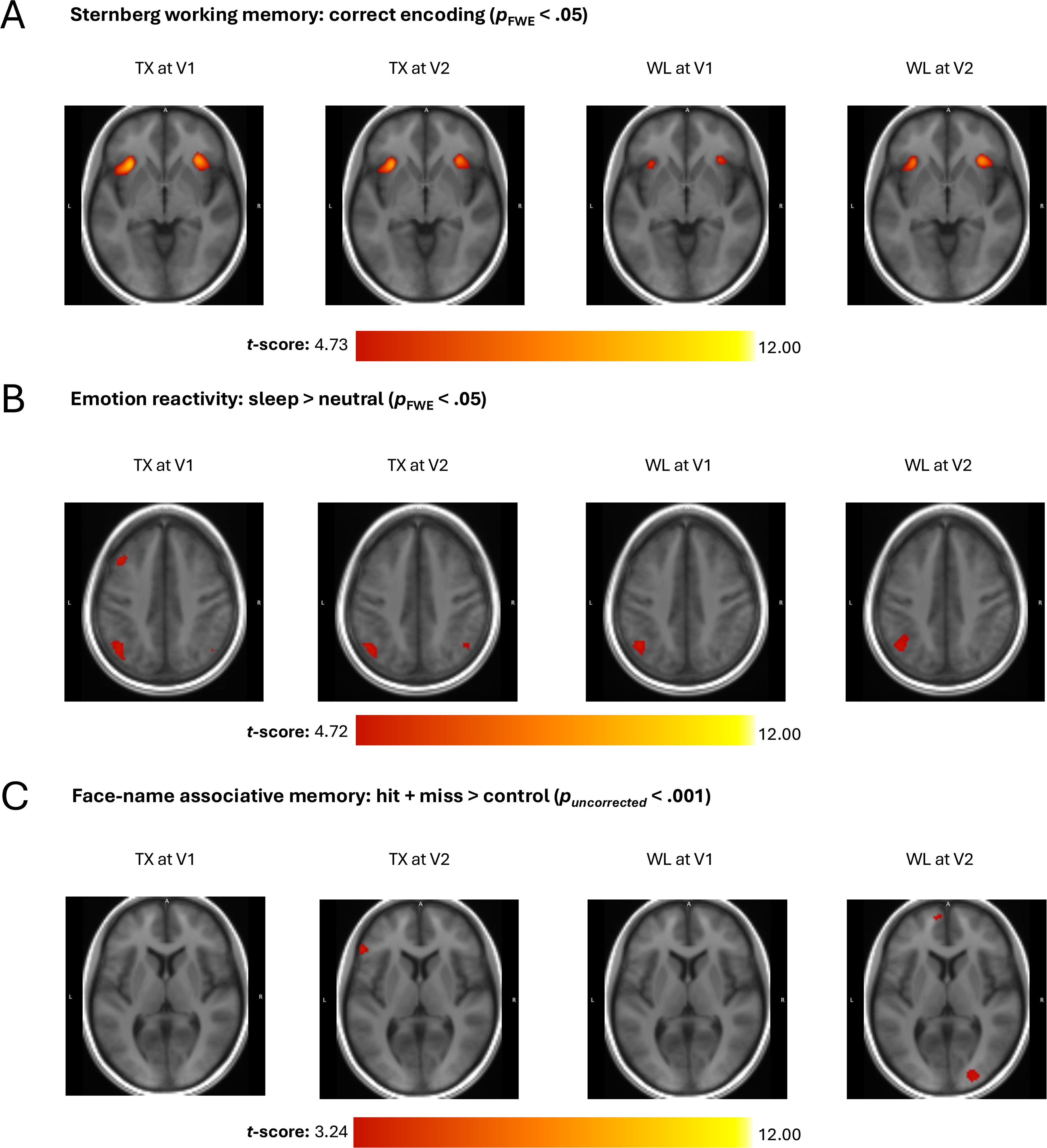
Effects of CBTi on brain activity during memory and emotion tasks (A) Brain activity in the TX and WL groups at V1 and V2 while correctly encoding digits during the Sternberg working memory task (B) Brain activity in the TX and WL groups at V1 and V2 in response to sleep relative to neutral images during the emotion reactivity task (C) Brain activity in the TX and WL groups at V1 and V2 during hit and miss trials relative to control trials for the face-name associative memory task As no voxels survived a family-wise error (FWE) correction set at *p* < .05 for the face-name associative memory task, the accompanying images in panel C display activations at an uncorrected threshold (*puncorrected* < .001).

Analyses revealed no statistically significant Group*Visit interaction or main effects of Group or Visit on within-network resting-state functional connectivity between regions within the DMN, SN, and FPN (all *p* > .05; Figure S4). These results did not differ whether 6 TX and 7 WL participants who were fighting sleep or fell asleep at any point during the resting-state fMRI session at V1 or at V2 were included or excluded (Figure S4). When comparing regions between DMN vs. SN, DMN vs. FPN, or SN vs. FPN, there was only a statistically significant main effect of Group. More precisely, the WL group exhibited lower connectivity between the right dorsal anterior cingulate cortex (SN region) and right dorsolateral prefrontal cortex (FPN region), on average, than the TX group (*z* = -3.63, *pFDR* = .04; Figure S5).

#### Secondary outcomes

Descriptive statistics (i.e., number of participants, means, and standard deviations) for self-reported cognition, emotion, and sleep measures of both TX and WL groups at V1, V2, or one-year follow-up can be found in Supplemental Material (Table S7). Results on the effects of CBTi on these self-reported measures at V1, V2, and one-year follow-up can also be found in Supplemental Material (Figure S6 and Table S8).

##### Self-reported cognition measure

Regarding cognition, there was a statistically significant Group*Visit interaction (*F*(1, 49.79) = 8.52; *p* = .02), but not main effects of Group or Visit, on total SASCI-Q scores. Post-hoc analyses revealed that while the WL group showed no change from V1 to V2 (*t*(51.39) = 0.69, *p* = .67), the TX group had improvements in self-reported cognitive impairment immediately after CBTi than before (*t*(51.77) = -3.46, *p* = .004; Figure 3A and Table 2). These improvements were also maintained one-year after completion of the CBTi intervention in the TX group (V1 vs. one-year follow-up: *t*(98.34) = -3.30, *p* = .01; Figure S6 and Table S8).

**Table 2.**
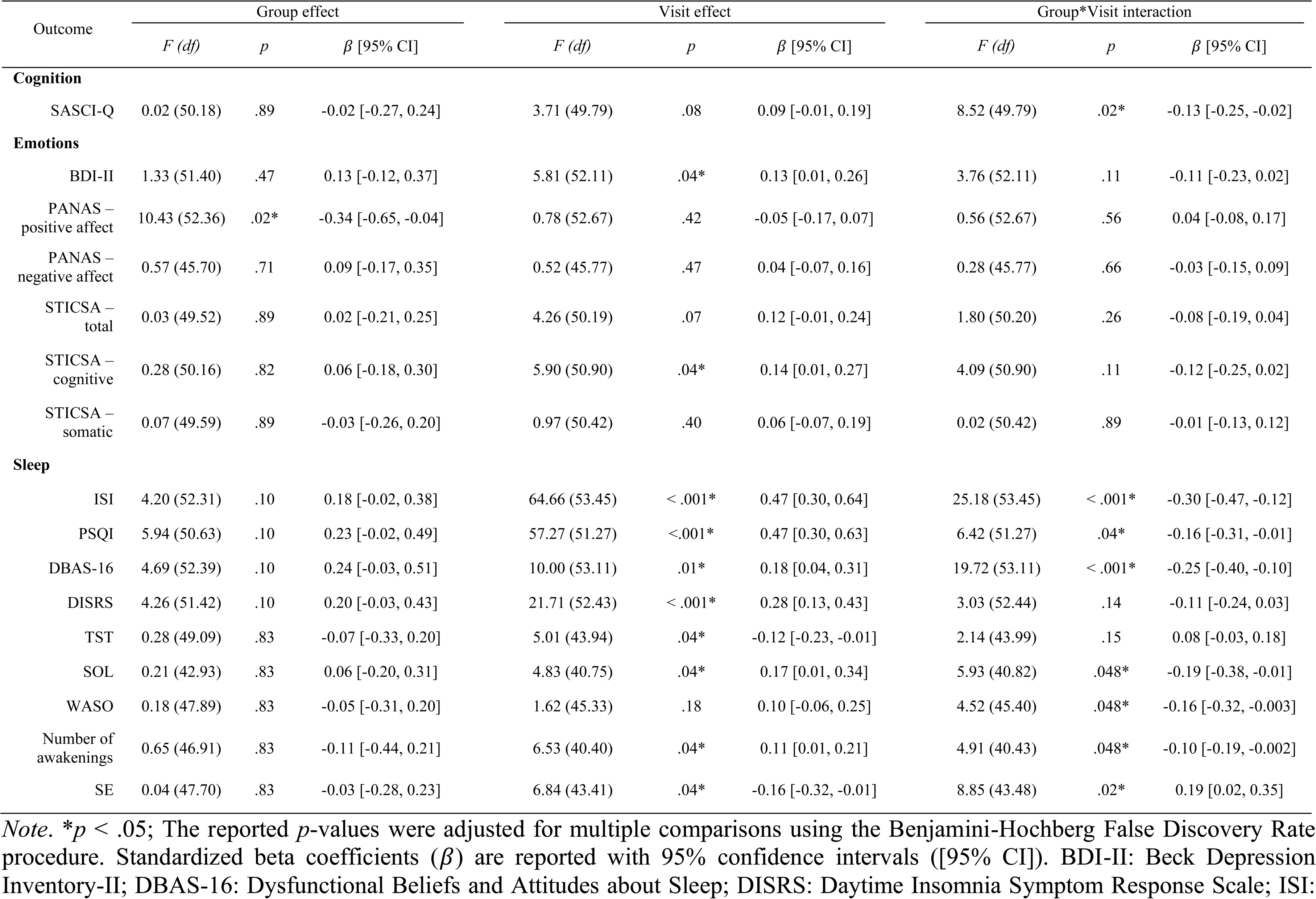
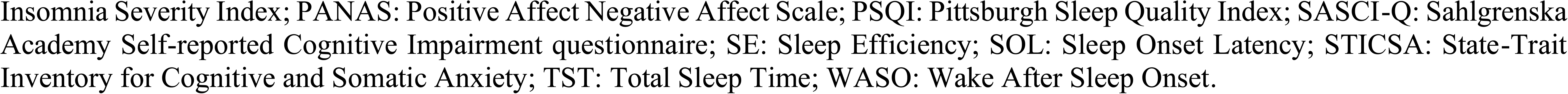
Self-reported measures of cognition, emotions, and sleep (V1 versus V2)

| Outcome | Group effect |  |  | Visit effect |  |  | Group*Visit interaction |  |  |
| --- | --- | --- | --- | --- | --- | --- | --- | --- | --- |
| | *F* (*df*) | *p* | $\beta$ [95% CI] | *F* (*df*) | *p* | $\beta$ [95% CI] | *F* (*df*) | *p* | $\beta$ [95% CI] |
| **Cognition** |  |  |  |  |  |  |  |  |  |
| SASCI-Q | 0.02 (50.18) | .89 | -0.02 [-0.27, 0.24] | 3.71 (49.79) | .08 | 0.09 [-0.01, 0.19] | 8.52 (49.79) | .02* | -0.13 [-0.25, -0.02] |
| **Emotions** |  |  |  |  |  |  |  |  |  |
| BDI-II | 1.33 (51.40) | .47 | 0.13 [-0.12, 0.37] | 5.81 (52.11) | .04* | 0.13 [0.01, 0.26] | 3.76 (52.11) | .11 | -0.11 [-0.23, 0.02] |
| PANAS – positive affect | 10.43 (52.36) | .02* | -0.34 [-0.65, -0.04] | 0.78 (52.67) | .42 | -0.05 [-0.17, 0.07] | 0.56 (52.67) | .56 | 0.04 [-0.08, 0.17] |
| PANAS – negative affect | 0.57 (45.70) | .71 | 0.09 [-0.17, 0.35] | 0.52 (45.77) | .47 | 0.04 [-0.07, 0.16] | 0.28 (45.77) | .66 | -0.03 [-0.15, 0.09] |
| STICSA – total | 0.03 (49.52) | .89 | 0.02 [-0.21, 0.25] | 4.26 (50.19) | .07 | 0.12 [-0.01, 0.24] | 1.80 (50.20) | .26 | -0.08 [-0.19, 0.04] |
| STICSA – cognitive | 0.28 (50.16) | .82 | 0.06 [-0.18, 0.30] | 5.90 (50.90) | .04* | 0.14 [0.01, 0.27] | 4.09 (50.90) | .11 | -0.12 [-0.25, 0.02] |
| STICSA – somatic | 0.07 (49.59) | .89 | -0.03 [-0.26, 0.20] | 0.97 (50.42) | .40 | 0.06 [-0.07, 0.19] | 0.02 (50.42) | .89 | -0.01 [-0.13, 0.12] |
| **Sleep** |  |  |  |  |  |  |  |  |  |
| ISI | 4.20 (52.31) | .10 | 0.18 [-0.02, 0.38] | 64.66 (53.45) | < .001* | 0.47 [0.30, 0.64] | 25.18 (53.45) | < .001* | -0.30 [-0.47, -0.12] |
| PSQI | 5.94 (50.63) | .10 | 0.23 [-0.02, 0.49] | 57.27 (51.27) | < .001* | 0.47 [0.30, 0.63] | 6.42 (51.27) | .04* | -0.16 [-0.31, -0.01] |
| DBAS-16 | 4.69 (52.39) | .10 | 0.24 [-0.03, 0.51] | 10.00 (53.11) | .01* | 0.18 [0.04, 0.31] | 19.72 (53.11) | < .001* | -0.25 [-0.40, -0.10] |
| DISRS | 4.26 (51.42) | .10 | 0.20 [-0.03, 0.43] | 21.71 (52.43) | < .001* | 0.28 [0.13, 0.43] | 3.03 (52.44) | .14 | -0.11 [-0.24, 0.03] |
| TST | 0.28 (49.09) | .83 | -0.07 [-0.33, 0.20] | 5.01 (43.94) | .04* | -0.12 [-0.23, -0.01] | 2.14 (43.99) | .15 | 0.08 [-0.03, 0.18] |
| SOL | 0.21 (42.93) | .83 | 0.06 [-0.20, 0.31] | 4.83 (40.75) | .04* | 0.17 [0.01, 0.34] | 5.93 (40.82) | .048* | -0.19 [-0.38, -0.01] |
| WASO | 0.18 (47.89) | .83 | -0.05 [-0.31, 0.20] | 1.62 (45.33) | .18 | 0.10 [-0.06, 0.25] | 4.52 (45.40) | .048* | -0.16 [-0.32, -0.003] |
| Number of awakenings | 0.65 (46.91) | .83 | -0.11 [-0.44, 0.21] | 6.53 (40.40) | .04* | 0.11 [0.01, 0.21] | 4.91 (40.43) | .048* | -0.10 [-0.19, -0.002] |
| SE | 0.04 (47.70) | .83 | -0.03 [-0.28, 0.23] | 6.84 (43.41) | .04* | -0.16 [-0.32, -0.01] | 8.85 (43.48) | .02* | 0.19 [0.02, 0.35] |

*Note.* \* $p < .05$ ; The reported $p$ -values were adjusted for multiple comparisons using the Benjamini-Hochberg False Discovery Rate procedure. Standardized beta coefficients ( $\beta$ ) are reported with 95% confidence intervals ([95% CI]). BDI-II: Beck Depression Inventory-II; DBAS-16: Dysfunctional Beliefs and Attitudes about Sleep; DISRS: Daytime Insomnia Symptom Response Scale; ISI:
Insomnia Severity Index; PANAS: Positive Affect Negative Affect Scale; PSQI: Pittsburgh Sleep Quality Index; SASCI-Q: Sahlgrenska Academy Self-reported Cognitive Impairment questionnaire; SE: Sleep Efficiency; SOL: Sleep Onset Latency; STICSA: State-Trait Inventory for Cognitive and Somatic Anxiety; TST: Total Sleep Time; WASO: Wake After Sleep Onset.

##### Self-reported emotion measures

In terms of measures of emotional states or traits, there were no statistically significant Group*Visit interaction, main effect of Group, or main effect of Visit on total negative affect, total STICSA, and STICSA-somatic scores (all *p* > .05; Table 2). There was a statistically significant main effect of Group on total positive affect scores, with the TX group having higher positive affect, on average, than the WL group. There was also statistically significant main effect of Visit on total BDI-II and STICSA-cognitive scores, with depressive and cognitive anxiety symptoms improving, on average, from V1 to V2. When including the one-year follow-up, there was still a statistically significant main effect of Group on total positive affect scores, with the TX group having higher positive affect, on average, than the WL group. There was also a statistically significant main effect of Visit on the total BDI-II, total STICSA, and STICSA-cognitive scores, reflecting improvements in depressive and anxiety symptoms in both TX and WL groups from V1 to the one-year follow-up. No statistically significant Group*Visit, main effect of Group, or main effect of Visit on total negative affect scores were found (Figure S6 and Table S8).

##### Self-reported sleep measures

There were no statistically significant Group*Visit interaction or main effects of Group or Visit on KSS scores before the resting-state fMRI task (all *p* > .05). There was also no statistically significant Group*Visit interaction or main effect of Group on KSS scores before the Sternberg, emotion reactivity, and face-name tasks. A statistically significant main effect of Visit on KSS scores before these three tasks showed that participants reported more sleepiness, on average, at V2 than at V1.

Based on sleep diaries, there was statistically significant Group*Visit interactions on SOL, number of awakenings, WASO, and SE (all *p* < .05), with the TX group specifically reporting improvements in these self-reported sleep measures at post-intervention. No statistically significant Group*Visit interaction or main effects of Group or Visit on TST were observed. When including the one-year follow-up, there was a marginally significant Group*Visit interaction (*F*(2, 86.43) = 4.85, *p* = .05) and a statistically significant main effect of Visit on SE (*F*(2, 86.31) = 9.67, *p* < .001). Post-hoc analyses revealed improvements in SE specifically from V1 to the one-year follow-up (*t*(88.89) = 4.39, *p* < .001). No statistically significant Group*Visit interaction or main effects on the other self-reported sleep measures were found (Table S8).

In terms of subjective sleep measures from the questionnaires, there were statistically significant Group*Visit interactions on total ISI (Figure S7), PSQI, and DBAS-16 scores. While the total ISI and DBAS-16 scores did not differ across visits for the WL group, the TX group had greater improvements at post-intervention compared to the WL group (*p* < .001). Moreover, these improvements in total ISI, PSQI, and DBAS-16 scores were maintained one-year after completion of the CBTi intervention in the TX and WL groups (Figure S6 and Table S8). Although there was no statistically significant Group*Visit interaction or main effect of Group on the total DISRS scores, there was a statistically significant main effect of Visit with ruminative thoughts when feeling tired decreasing, on average, from V1 to V2 (Table 2). When including the one-year follow-up, there was a statistically significant main effect of Visit, with DISRS scores, on average, improving from V1 to the one-year follow-up. Moreover, the Group*Visit interaction on the total DISRS scores was marginally significant (*F*(2, 99.59) = 3.68, *p* = .06; Figure S6 and Table S8).

Based on the total ISI scores, 61% of participants in the TX group were considered as treatment responders (total ISI score decreasing by at least 8 points) following CBTi. Among those responders, 76% of them achieved remission (total ISI score lower than 8). On the contrary, only 19% of participants in the WL group showed clinically significant improvement, though 80% of those responders reached remission at V2. Overall, the TX group demonstrated both a significantly higher number of responders (*χ*^2^(1, *N* = 54) = 7.97, *p* = .005) and a higher total remission rate compared to the WL group (46% vs. 15%, respectively; *χ*^2^(1, *N* = 54) = 4.67, *p* = .03). Results showing no statistically significant effects of CBTi on cognitive performance and associated neural correlates during the MRI tasks stratified by group (CBTi responders vs. non-responders) are presented in Supplemental Material (Tables S9–S10).

##### Objective measures of sleep

No statistically significant Group*Visit interaction, main effects of Group or Visit on PSG-derived TST, SOL, WASO, and SE were found (Table S11).

## DISCUSSION

This RCT primarily investigated the effects of CBTi on cognitive and emotional functioning, assessed subjectively and objectively, in individuals with insomnia disorder and baseline differences with good sleepers. At baseline, individuals with insomnia disorder did not differ from good sleepers in resting-state functional connectivity, performance, or brain activity during memory tasks. Despite no differences in brain activity during the emotion reactivity task, individuals with insomnia disorder perceived sleep-related, but not neutral or negative images, as more negative than good sleepers. Furthermore, individuals with insomnia disorder reported poorer sleep, cognitive, and emotional functioning than good sleepers. No statistically significant treatment-related changes in valence ratings of sleep images or brain activity during cognitive tasks or at rest were detected. Despite the absence of changes in underlying brain function or objective cognitive performance, CBTi modifies subjective perceptions of sleep and of cognitive functioning that are maintained at the one-year follow-up.

### Comparison between insomnia and good sleeper participants

At baseline, behavioural differences between individuals with insomnia disorder and good sleepers emerged selectively during the emotion reactivity task, while performance remained similar across both memory tasks. Replicating findings from previous studies,^85,86^ participants with insomnia disorder only perceived sleep-related images as more negative than good sleepers. These sleep images, mainly depicting scenes of insomnia (e.g., a person lying awake in bed), may generate feelings of anxiety or emotional distress, promoting selective attention to sleep-related threat cues, and subsequently reinforcing excessive worrying and rumination, as proposed in the cognitive model of insomnia.^87^ Consistent with the work of Drummond and colleagues,^27^ participants with insomnia disorder and good sleepers had comparably high performance levels (> 90%) during the Sternberg working memory task. Furthermore, despite a growing interest in socially relevant memory, such as face-name associative memory, research in insomnia remains scarce. Although one previous study noted poorer accuracy on a face-matching task that does not rely on recognition memory in individuals with sleep disturbances,^88^ these findings did not extend to the present study that evaluated associative memory in individuals with insomnia disorder.

Brain activity during memory and emotion tasks did not differ between participants with insomnia disorder and good sleepers at baseline. Despite subjective cognitive complaints among those with insomnia disorder, both memory tasks may not have been sensitive enough to detect differences nor were they challenging enough for participants to require additional neural resources. Additionally, this discrepancy can suggest that individuals with this disorder may experience an increased sense of perceived effort despite utilizing neural resources comparable to those of good sleepers.^27^ Furthermore, our sample was younger than those in a previous study using verbal fluency tasks.^51^ As insomnia can increase the risk of memory decline in older individuals,^89^ age differences may account for the lack of group differences in memory performance observed in the current study. Regarding emotion reactivity, previous studies found that individuals with insomnia disorder exhibited heightened amygdala activity in response to insomnia-related images and greater functional connectivity involving the amygdala while viewing humorous films compared to good sleepers.^34^, However, one key difference in task design is that the arousal levels were consistent across all three image categories in the present study to isolate baseline differences driven solely by image content (i.e., whether they were sleep-related or not). In contrast, Baglioni and colleagues’^34^ study compared different image categories with varying arousal levels against neutral images with low arousal levels. As the current study focused specifically on moderate arousal levels, our findings provide valuable methodological insight for future studies, suggesting that more intense sleep-related or emotional stimuli may be required to significantly increase activity in the emotional neural circuitry, including the amygdala, in this population.

Moreover, no group differences in resting-state functional connectivity involving the DMN, FPN, or SN at baseline were detected. While one previous study did report lower resting-state functional connectivity between DMN subregions in individuals with insomnia disorder compared to good sleepers, these alterations were not linked to any clinical measures.^91^ As the DMN and FPN support memory function, including maintaining and manipulating information in working memory,^92,93^ the intact functional connectivity within these networks aligns with the preserved behavioural performance observed among the participants with insomnia disorder. The SN has also been a focal point in understanding the pathophysiology of insomnia disorder, largely due to its involvement in reactivity to new, important, or emotional stimuli.^94^ Our findings regarding the SN reinforce the growing evidence from a few studies,^46,95^ showing comparable resting-state functional connectivity between participants with insomnia disorder and good sleepers at baseline. While these studies along with ours primarily assessed mean functional connectivity strength within minutes rather than fluctuations in functional connectivity at finer time scales, it is possible that group differences may only emerge when examined at a more granular level.^46^ Moreover, one study found greater functional connectivity between the anterior insula and the SN only while participants with insomnia disorder were falling asleep compared to good sleepers.^95^ As our participants were instructed to stay awake, this procedural difference could account for these divergent findings and can mean that functional connectivity patterns may be state-dependent or scale with the severity of insomnia symptoms, such as when individuals are trying to fall asleep or are experiencing elevated hyperarousal. Importantly, the clinical presentation of insomnia disorder is highly heterogeneous in symptom profiles, etiology, and causes across individuals, which also may have impacted group analyses.^96–99^ Collectively, the absence of group differences in task-based brain activity, resting-state functional connectivity, and memory performance at baseline align with the perspective that insomnia disorder is defined more by subjective complaints of sleep and altered perception of cognitive functioning than by objective physiological deficits alone.

In accordance with previous research and reports from people living with insomnia disorder,^7–8,10,90,100–106^ participants with insomnia disorder had more subjective cognitive impairment, emotional disturbances, and sleep difficulties than good sleepers at baseline. Consistent with previous PSG studies,^107–110^ participants with insomnia disorder had greater WASO than good sleepers at baseline. However, no significant group differences emerged for any other sleep measure.^8,111^ Many individuals complain about the impacts of their sleep disturbance on daytime functioning.^7,87,112^ However, the objective tests used in the study did not fully capture the patient experience. This may be due to some literature suggesting those with insomnia disorder show excessive worry and catastrophizing about sleep.^87^ Nevertheless, these findings highlight the importance of using both subjective and objective measures to accurately assess sleep disturbances.

### Effects of CBTi

Brain activity during the memory and emotion tasks as well as resting-state functional connectivity within the DMN, FPN, or SN remained unchanged following CBTi. This may indicate that the tasks were not sensitive enough to detect treatment-specific neural changes, especially given that no group differences with good sleepers were observed at baseline. As this is the first study to assess the effects of CBTi on both working and associative memory in individuals with insomnia disorder, more research is warranted to clarify potential neural changes associated with the treatment in the long-term. Regarding emotions, previous research found increased amygdala activity in response to negative facial expressions in individuals with insomnia disorder who have a higher risk of developing depression after receiving both CBTi and internet-based circadian rhythm support (CRS) compared to no treatment, CBTi alone, or CRS alone.^113^ Conversely, individuals with insomnia disorder had less activity in various brain regions in response to sleep-related images or sounds after receiving CBTi alone.^52,53^ Nevertheless, variations in task design, stimuli type, treatment protocols, and clinical characteristics may account for these opposing findings. These results may also offer insight, indicating that CBTi may lead to clinical improvements by targeting sleep or cognitive pathways rather than by directly modifying the neural circuitry underlying emotion reactivity. Only one prior resting-state fMRI study reported increased functional connectivity in other memory-related regions, such as the hippocampus, as well as reduced functional connectivity in emotion-related regions following CBTi.^28^ This lack of effect in our study may be attributed to the fact that we did not exclusively recruit participants with psychophysiological insomnia, as was done by Lee and colleagues.^28^ As this previous study did not monitor alertness or sleepiness as was done in the current study, their findings may have also been reporting state rather than trait effects.^28^

In this present study, no changes in working or associative memory performance in participants with insomnia disorder were found following CBTi. Our finding is consistent with the only other study using the Sternberg working memory task specifically in adults with insomnia disorder and good sleepers taking eszopiclone or placebo as well as with another digital CBTi study using a different working memory task.^114,115^ Although the task was fairly easy for individuals with insomnia disorder and good sleepers, it is possible that the patients may have required more effort to perform equally well. On the contrary, our TX and WL groups, on average, were less accurate on the face-name memory associative task and had higher KSS scores at three months post-randomization. One explanation for this is that this cognitive task was performed last, potentially leading to fatigue. Mental fatigue can lead to both task disengagement and decreased memory performance.^116^ Alternatively, participants may have been less motivated at V2 than at V1 due to the tasks being less novel. It is also possible that no effects of CBTi were observed because there were no baseline differences between INS and GS groups. Consequently, these fMRI tasks may not be sensitive to detect subtle treatment-specific changes.

Although we anticipated improvements in valence rating for sleep images following CBTi, participants with insomnia disorder did not habituate to these images more than to neutral or negative ones. This distinct pattern may reflect that attentional biases toward sleep stimuli are more persistent in individuals with insomnia disorder.^117–123^ Different cognitive models of insomnia have been proposed to explain this bias.^87,124^ In addition to the aforementioned cognitive model of insomnia,^87^ Espie and colleagues^124^ later proposed the attention-intention-effort pathway that characterizes the sleep-wake process as normally automatic. However, in the case of insomnia disorder, this process is inhibited through selective attention to sleep or efforts to control it. Consequently, the shift from automaticity to an attentional bias to sleep can create a self-perpetuating cycle of distress that maintains the disorder.

Participants with insomnia disorder in the current study showed significant improvements in subjective cognitive impairment following CBTi, whereas no changes were observed following the wait-list period. Consistent with the literature on cognitive function in insomnia,^7^ our findings confirm that CBTi ameliorates subjective rather than objective cognitive functioning. This discrepancy is analogous to sleep-state misperception, the discrepancy between subjective and objective sleep measures, that has been noted throughout the literature in this population.^7,125^ These self-reported cognitive improvements were also maintained at the one-year follow-up, highlighting the long-lasting benefits of CBTi. By learning to use cognitive restructuring to replace negative thoughts about sleep with more adaptive ones,^126^ individuals receiving CBTi may develop broader cognitive flexibility, enabling them to apply these skills to negative thoughts about their cognition or other daytime impairments. These findings extend our earlier study with a different cohort,^7^ which noted a trend for improvements in subjective cognitive impairment after group CBTi. The more robust results observed here likely stems from a more homogeneous sample in terms of demographic characteristics and stricter exclusion criteria regarding psychiatric comorbidities. As such, while the baseline depressive and anxiety scores were higher in the INS group compared to the GS group, they remained below clinical significance.^72,74^ Nevertheless, both TX and WL groups had self-reported improvements in depressive and anxiety symptoms from baseline to three months post-randomization and were maintained at the one-year follow-up, potentially showing long-lasting benefits of the intervention on these mental health outcomes. Furthermore, positive and negative affect remained stable over time in both insomnia groups, suggesting that the PANAS may reflect stable personality traits rather than acute shifts in affective states in individuals with insomnia disorder.^73,102^

The observed improvements in self-reported insomnia severity, sleep quality, dysfunctional beliefs and attitudes about sleep, and daytime ruminative thoughts in participants with insomnia disorder following CBTi and one-year after completion of the CBTi intervention align with the treatment effects documented in recent RCTs and meta-analyses.^7,127–129^ Similar to our previous work with a different cohort,^7^ we also found statistically significant group-by-visit interactions on certain sleep diary measures (e.g., SOL, number of awakenings, time spent awake after sleep onset, and SE) immediately after CBTi. Consistent with prior studies,^6–7,130^ we found no significant changes in objective sleep microarchitecture measures in individuals with insomnia disorder immediately after CBTi. Thus, this finding provides further evidence that CBTi primarily ameliorates self-reported rather than objective measures of sleep.

### Strengths and Limitations

The most notable strength of the study was its longitudinal, multimodal, and multilevel RCT design with the use of self-report questionnaires, PSG, and MRI (during tasks and rest) to capture a comprehensive picture of both subjective and objective sleep, cognition, and emotional functioning. Moreover, during fMRI tasks, we objectively monitored alertness and sleepiness using the KSS unlike other studies, allowing us to isolate the effects of the chronic condition (i.e., insomnia disorder) from transient states (i.e., sleepiness). In addition to having two control groups (i.e., good sleepers and the WL group), participants were matched based on age, sex, and education level. Another strength of this study is that individual CBTi was employed rather than CBTi in a group setting. Although both individual and group CBTi were previously shown to be beneficial for sleep, individual CBTi may yield better subjective and objective sleep outcomes than group CBTi.^131^

Some limitations should be taken into consideration when interpreting the current findings. First, the overall sample was highly educated and middle-aged, which could reflect better cognitive reserve than the general population.^132^ This may explain why participants performed exceptionally well on the Sternberg working memory task. Another potential limitation is that the generalizability of the findings may be limited by our stringent exclusion criteria, such as individuals with comorbidities or those who used sleep medications. Lastly, due to the heterogeneity of insomnia,^96–99^ larger sample sizes might be required for more sensitive analyses including insomnia profiling.

## Conclusion

The current findings reveal that CBTi has clinical implications beyond just sleep, especially when it comes to subjective cognitive functioning. Improvement in self-reported cognitive difficulties may represent a meaningful daytime benefit of CBTi, even when changes are not detected on laboratory memory tasks. Importantly, this intervention can be optimal for individuals with insomnia disorder who particularly have greater worries about their cognitive functioning or experience more misperception. To better address emotional distress brought on by insomnia, clinicians could incorporate more emotion regulation strategies that target sleep stimuli to the standard CBTi protocol, as a combined CBTi and emotion regulation intervention has recently been shown to be effective for improving insomnia, depressive, and anxiety symptoms.^133^ Future research should replicate this study with more extended follow-up sessions to determine whether there are functional brain changes or objective cognitive improvements that emerge as participants with insomnia disorder further consolidate and integrate the learned CBTi strategies. Future research should also delineate the neural substrates and psychological processes underlying these subjective cognitive deficits and clarify the mechanisms driving the dissociation between subjective and objective cognitive functioning in insomnia disorder. In conclusion, this study demonstrates that the long-lasting benefits of CBTi are primarily evidenced through subjective rather than objective improvements in sleep and cognitive functioning. These improvements suggest that CBTi strategies effectively shift subjective perception of sleep and cognitive functioning, despite a lack of change in corresponding objective measures.

## Acknowledgments

We acknowledge the contributions of the following students who assisted with participants’ recruitment, data collection, and data preprocessing: Jonah Miller-Smith, Saba Sarani, Tinhinane Kessai, Mila Goulon, Victoria D’Amours, Kazem Habibi-Tanha, Zehra Güngör, Ilona Scellier-Dekens, Franck Vilcourt, Alesar Hamati, Kimia Motevalli, Dania Takach, Claire Dyment, Ophélia Fontaine, Quin Lagden, and Clara Vo-Dignard. We also acknowledge the contribution of the psychologists who provided CBTi: Caroline Desrosiers, Rosemarie Perrault, Chantal Paquin, Warren Caldwell, Nyissa Walsh, and Oren Weiner. We thank the MRI technicians, Antonys Melek and Dwayne Thompson, who helped acquire the MRI data and collect blood samples. We also thank our sleep technologists Madeline Dickson and Elinah Mozhentiq for their contribution to the setup of PSG recordings and overnights. Finally, we would like to thank all the participants who generously gave their time to participate in the study.

## List of abbreviations

BDI-II: Beck Depression Inventory-II
CBTi: Cognitive-Behavioural Therapy for insomnia
CR: Correct Rejection
CRS: Circadian Rhythm Support
DBAS-16: abbreviated version of the Dysfunctional Beliefs and Attitudes about Sleep (16 items)
DISRS: Daytime Insomnia Symptom Response Scale
DMN: Default Mode Network
EEG: Electroencephalography
EMG: Electromyography
Emmeans: Estimated Marginal Means
EOG: Electrooculography
FA: False Alarm
fMRI: Functional Magnetic Resonance Imaging
FDR: False Discovery Rate
FPN: Frontoparietal Network
FWE: Family-Wise Error
FWHM: Full Width at Half Maximum
GS: Good Sleeper
ICC: Intraclass Correlation Coefficient
ICSD: International Classification of Sleep Disorders
INS: Insomnia Disorder
IQ: Intelligence Quotient
ISI: Insomnia Severity Index
KSS: Karolinska Sleepiness Scale
lmer: Linear Mixed-Effects Models
MDFFITS: Multivariate Difference in Fits
MINI: Mini International Neuropsychiatric Interview
MNI: Montreal Neurological Institute
MRI: Magnetic Resonance Imaging
MTN: Medial Temporal Network
PANAS: Positive Affect Negative Affect Scale
PSG: Polysomnography
PSQI: Pittsburgh Sleep Quality Index
RCT: Randomization Controlled Trial
SASCI-Q: Sahlgrenska Academy Self-reported Cognitive Impairment Questionnaire
SE: Sleep Efficiency
SEM: Standard Error of the Mean
SN: Salience Network
SOL: Sleep Onset Latency
STICSA: State-Trait Inventory for Cognitive and Somatic Anxiety
TMS: Transcranial Magnetic Stimulation
TST: Total Sleep Time
TX: Treatment group
WASO: Wake After Sleep Onset
WL: Wait-list control group

## Author contributions

Conceptualization: TDV, JPG, AAP, FB; Data collection: KG, EMP, AAP, FB, NAW, AM, JG, LT; Data curation: KG, EMP, AAP, NAW; Formal analysis: KG, EMP; Methodology: AAP, FB; Funding acquisition: TDV, JPG; Project coordination/administration: LT, AAP, FB; Visualization: KG; Roles/Writing-original draft: KG, FB, EMP, TDV; Writing-review & editing: KG, FB, EMP, TDV, AAP, NAW, MR, AM, JG, JPG, JJM, JS, SS.

## Funding

This research was supported by grants to KG, TDV, and JPG from the Canadian Institutes of Health Research (reference number: 236-2021-2022-Q3-00089; MOP 142191, PJT153115), Concordia University, Center for Studies in Behavioral Neurobiology, and Quebec Bio-Imaging Network.

AAP received grants from the formerly called PERFORM Center, Concordia University and Centre de Recherche de l’Institut Universitaire de Gériatrie de Montréal.

## Disclosure Statement

Financial disclosure and non-financial disclosure: None to report.

## Data availability

The data underlying this article will be shared on reasonable request to the corresponding author.

## Notes

### Competing Interest Statement

The authors have declared no competing interest.

### Clinical Trial

NCT04024787

### Author Declarations

Ethics committee/IRB of Concordia University gave ethical approval for this work.

