## Supplemental file for "Effects of cognitive-behavioural therapy for insomnia on cognitive and emotional functioning in adults with insomnia: behavioural and neuroimaging evidence"

### **Supplementary Methods**

#### **Secondary outcomes**

*Sahlgrenska Academy Self-reported Cognitive Impairment Questionnaire (SASCI-Q)*

The Sahlgrenska Academy Self-reported Cognitive Impairment Questionnaire (SASCI-Q) is a 29-item measure that assesses subjective cognitive functioning within the past two weeks.^[1]^ Cognitive domains include memory, attention/speed, executive functioning, and language. Items are rated on a 7-point Likert scale (1 = never to 7 = almost all the time), with a higher score indicating more self-report of cognitive impairment. The internal consistency, as assessed with Cronbach’s alpha, of the SASCI-Q scores at V1 was .92 and .94 for the GS and INS groups, respectively. At V2, the internal consistency was .96 for the INS group. The internal consistency for the INS group was .96 both at V2 and 1YFU. Test-retest reliability was assessed only in the WL group using intraclass correlation coefficients (ICC),^[2]^ given that no changes were expected between V1 and V2 in the absence of CBTi. Total SASCI-Q scores from V1 to V2 in the WL group showed good test-retest reliability (ICC(3,1) = .82, 95% CI [.64, .92]).

*Beck Depression Inventory (BDI-II)*

The Beck Depression Inventory (BDI-II) is a 21-item measure on a 4-point Likert scale that assesses the severity of depressive symptoms within the past two weeks. The total BDI-II score ranges from 0 to 63 and are classified as minimal (0–13), mild (14–19), moderate (20–28), and severe (29–63).^[3]^ The internal consistency of the BDI-II scores at V1 was .98 and .87 for the GS and INS groups, respectively. The internal consistency for the INS group was .94 at V2 and .87 at the 1YFU. Total BDI-II scores from V1 to V2 in the WL group demonstrated moderate test-retest reliability (ICC(3,1) = .54, 95% CI [.20, .77]).

*Positive Affect Negative Affect Scale (PANAS)*

The Positive Affect Negative Affect Scale (PANAS) measures affective state over the past two weeks. 10 positive (e.g., proud or inspired) and 10 negative (e.g., distressed or scared) emotions or feelings are assessed on a 5-point Likert scale (1 = not at all to 5 = very much). Each subscale is independently scored from 10 and 50. Higher scores indicate greater positive or negative affect, depending on the subscale.^[4]^ The internal consistency of the PANAS positive affect scores at V1 was .91 for both the GS and INS groups. The internal consistency of the PANAS positive affect scores for the INS group was .93 at V2 and .92 at the 1YFU. Furthermore, the internal consistency of the PANAS negative affect scores at V1 was .82 and .84 for the GS and INS groups, respectively. The internal consistency of the PANAS negative affect scores for the INS group was .92 at V2 and .89 at the 1YFU. PANAS positive affect scores (ICC(3,1) = .70, 95% CI [.44, .86]) and negative affect scores (ICC(3,1) = .70, 95% CI [.44, .85]) from V1 to V2 in the WL group demonstrated moderate test-retest reliability.

*State-Trait Inventory for Cognitive and Somatic Anxiety (STICSA)*

The State-Trait Inventory for Cognitive and Somatic Anxiety (STICSA) is a 21-item measure that assesses both cognitive and somatic anxiety within the past two weeks. The items are rated on a 4-point Likert scale (1 = almost never to 4 = almost always). The total STICSA score ranges from 21 to 84. Subscale scores were computed by summing the 10 cognitive symptom items (i.e., STICSA-cognitive) and 11 somatic symptom items (i.e., STICSA-somatic). For all scales, higher scores indicate greater anxiety.^[5]^ The internal consistency of the STICSA-cognitive scores at V1 was .80 and .88 for the GS and INS groups, respectively. The internal consistency of the STICSA-cognitive scores for the INS group was .89 at V2 and .88 at the 1YFU. The internal consistency of the STICSA-somatic scores at V1 was .48 and .82 for the GS and INS groups, respectively. The internal consistency of the STICSA-somatic scores for the INS group was .90 at V2 and .70 at the 1YFU. STICSA cognitive scores (ICC(3,1) = .67, 95% CI [.39, .84]) and somatic scores (ICC(3,1) = .70, 95% CI [.44, .85]) from V1 to V2 in the WL group demonstrated moderate test-retest reliability.

*Insomnia Severity Index (ISI)*

The Insomnia Severity Index (ISI) is a seven-item questionnaire evaluating the severity of insomnia symptoms within the past two weeks.^[6]^ Items are rated on a 5-point Likert scale (0 = no problem to 4 = very severe problem), yielding a total score out of 28 points with higher scores indicating greater insomnia severity. At V1, the internal consistency of the ISI scores was .70 and .63 for the GS and INS groups, respectively. The internal consistency for the INS group was .90 at V2 and .83 at the 1YFU. Total ISI scores from V1 to V2 in the WL group demonstrated moderate test-retest reliability (ICC(3,1) = .50, 95% CI [.15, .74]).

*Pittsburgh Sleep Quality Index (PSQI)*

The Pittsburgh Sleep Quality Index (PSQI) is a measure that assesses general sleep quality within the past month across seven components: subjective sleep quality, sleep latency, sleep duration, sleep efficiency, sleep disturbance, use of sleep medication, and daytime dysfunction. The total PSQI score ranges from 0 to 21, with higher scores indicating worse sleep quality.^[7]^ The internal consistency of the PSQI scores at V1 was .27 and .37 for the GS and INS groups, respectively. The internal consistency for the INS group was .75 at V2 and .56 at the 1YFU. Total PSQI scores from V1 to V2 in the WL group indicated poor test-retest reliability (ICC(3,1) = .49, 95% CI [.13, .73]).

*Dysfunctional Beliefs and Attitudes about Sleep (DBAS-16)*

The abbreviated version of the Dysfunctional Beliefs and Attitudes about Sleep (DBAS-16) assesses personal beliefs and attitudes about sleep using 16 items rated on a 0–10 scale (0 = strongly disagree to 10 = strongly agree). The total score is calculated as the mean of all 16 items and ranges from 0 to 10, with higher scores reflecting greater dysfunctional cognitions about sleep.^[8]^ At V1, the internal consistency of the DBAS-16 scores was .94 and .85 for the GS and INS groups, respectively. The internal consistency for the INS group was .93 at V2 and .89 at the 1YFU. Total DBAS-16 scores from V1 to V2 in the WL group demonstrated moderate test-retest reliability (ICC(3,1) = .70, 95% CI [.44, .86]).

*Daytime Insomnia Symptom Response Scale (DISRS)*

The Daytime Insomnia Symptom Response Scale (DISRS) is a 20-item measure on a 4-point Likert scale (1 = almost never to 4 = almost always) that assesses the severity of daytime impairment or ruminative thoughts when feeling tired. The total DISRS score ranges from 20 to 80, with higher scores indicating more rumination.^[9]^ At V1, the internal consistency of the DISRS scores was .97 and .92 for the GS and INS groups, respectively. The internal consistency for the INS group was .96 at V2 and .95 at the 1YFU. Total DISRS scores from V1 to V2 in the WL group demonstrated moderate test-retest reliability (ICC(3,1) = .57, 95% CI [.24, .78]).

*Sleep diaries*

Computerized daily sleep diaries, based on the Consensus Sleep Diary and Social Rhythm Metric, were used to assess treatment efficacy on sleep and stability of daily routine at each timepoint.^[10],^^[11]^ For two weeks, participants answered 19 questions every morning about sleep and daily activity. Sleep variables of interest, averaged over the two weeks, included subjective total sleep time (TST), sleep onset latency (SOL), wake after sleep onset (WASO), and sleep efficiency (SE). Only sleep diary data with at least seven days completed per timepoint were included in the analyses.

*Karolinska Sleepiness Scale (KSS)*

The Karolinska Sleepiness Scale (KSS) is a single item measure on a 9-point Likert scale (1 = extremely alert to 9 = very sleepy, fighting sleep) that assesses momentary alertness or sleepiness within the previous five minutes.^[12]^

**Polysomnographic (PSG) recording**

Full-night PSG was recorded in-lab using the 32-channel SOMNOscreen plus device with synchronized video (SOMNOmedics GmbH, Germany). Data were acquired with a sampling rate of 512 Hz. PSG included EEG, EOG, EMG, ECG. Eleven scalp EEG electrodes (F3, Fz, F4, C3, Cz, C4, Pz, O1, O2, M1, and M2) were placed according to the international 10-20 system. During the first overnight visit (V0) only, nasal-oral thermocouple airflow, transcutaneous finger pulse oximeter, and thoracic-abdominal belts were added to assess breathing and screen for sleep apnea. Electrodes were also placed on both legs to screen for periodic limb movement. EEG signals were referenced online to Pz and re-referenced offline to the linked mastoids (M1 and M2). All sleep scoring and analyses were conducted using the Wonambi python toolbox (https://wonambi-python.github.io). For each recording, two scorers blind to allocation determined the different sleep stages (NREM 1, 2, 3, REM sleep and wake) according to the AASM rules for each recorded night of sleep.^55^ From the scoring of the sleep architecture, we computed TST, SOL, WASO, and SE using the seapipe python toolbox (<https://github.com/nathanecross/seapipe>).

#### **Figure S1. Design of the cognitive tasks**

(A) Task design for the Sternberg working memory task.

(B) Task design for the emotion reactivity task.

(C) Task design for the face-name associative memory task. The encoding portion was done in the MRI scanner. The recall portion was done immediately after the task and the following morning both outside of the MRI scanner.

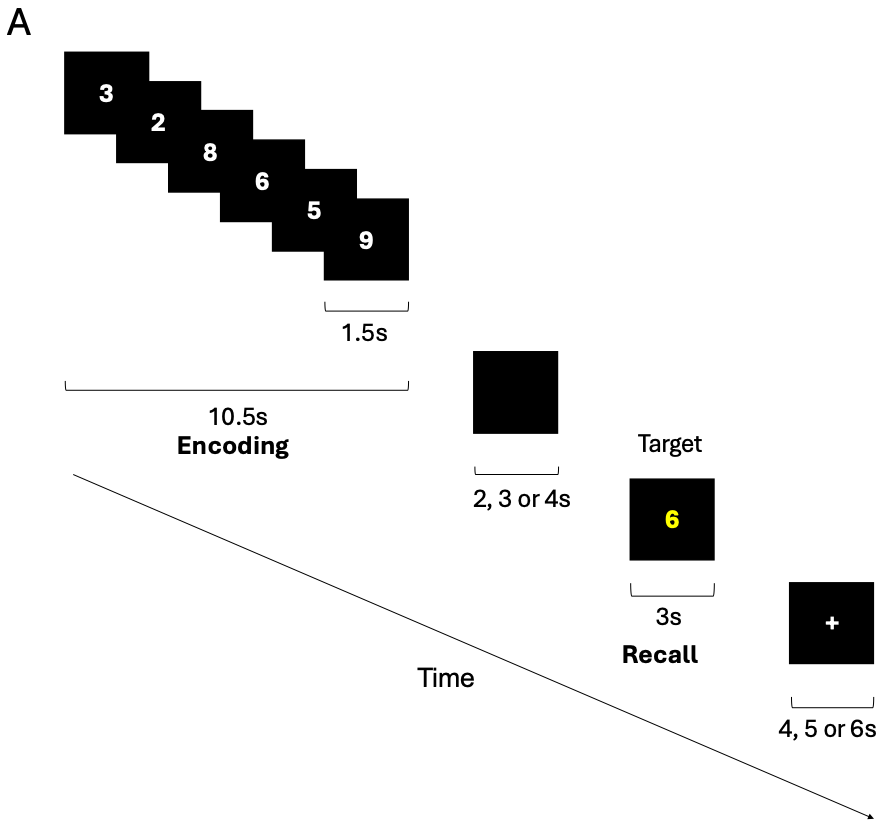

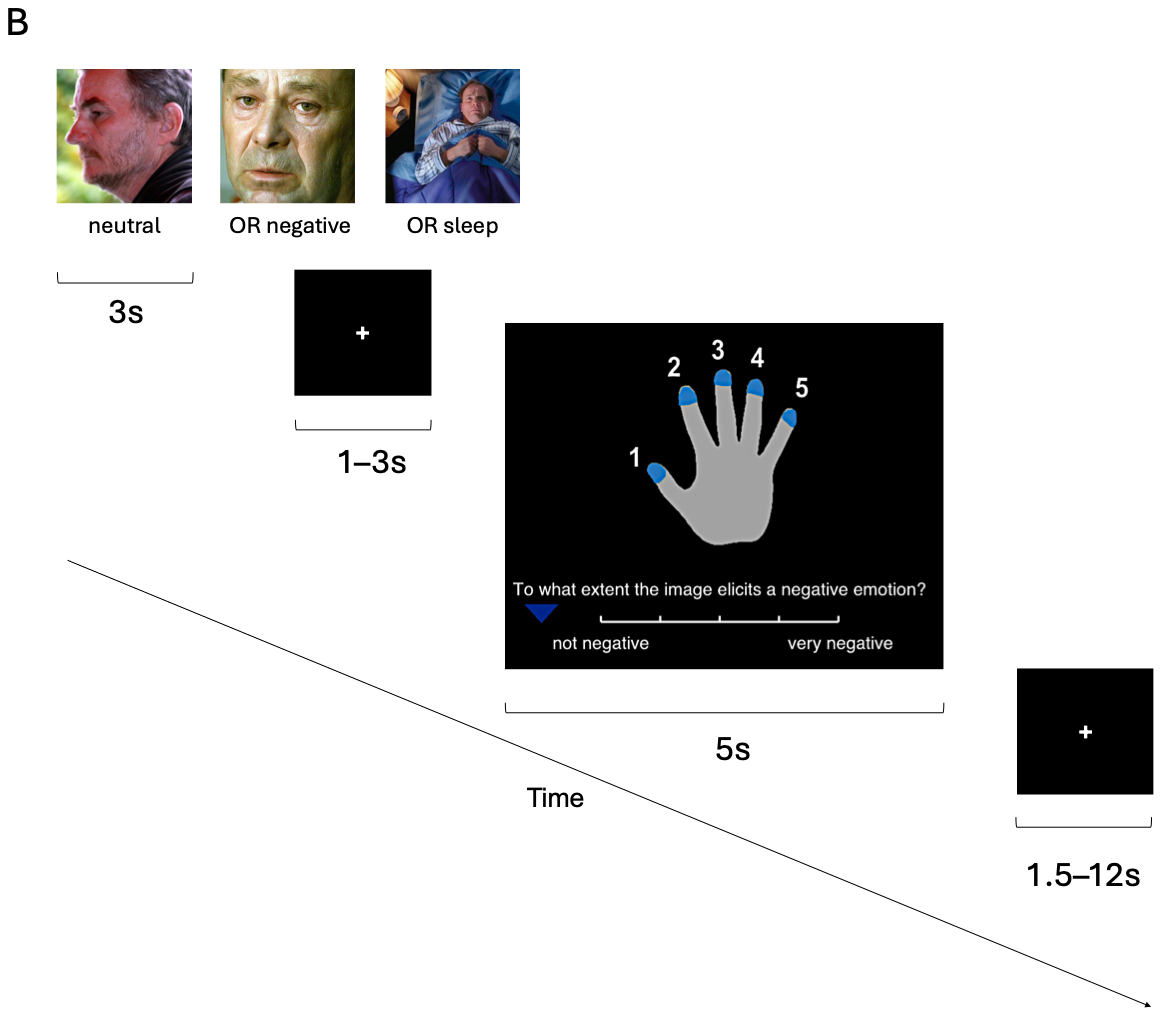

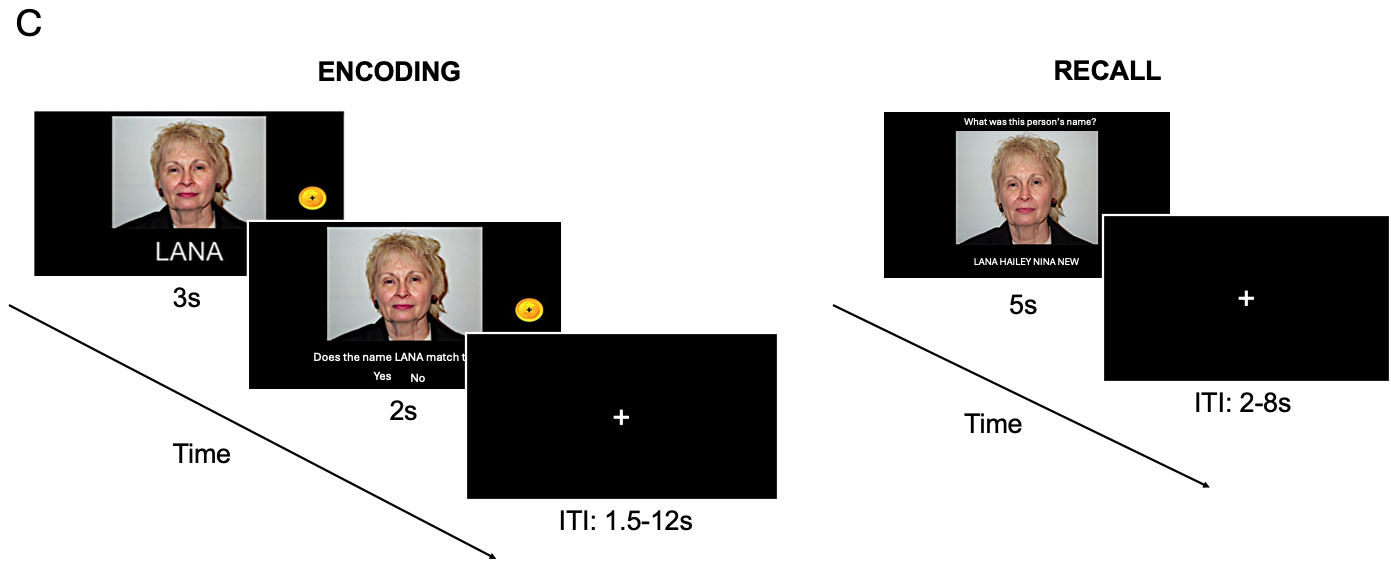

#### **Database of emotional images**

Out of the 120 images that were used in either the first or second version of the emotion reactivity task, 40 negative and 40 neutral images came from the Nencki Affective Picture System (NAPS) database.^[13]^ Ten sleep-related images came from the database that was used by Baglioni et al.^[14]^ and 30 were obtained from the Internet. Prior to the start of the study, these sleep-related images were validated on Amazon Mechanical Turk by good sleepers (*n* = 30; ISI < 7) and individuals with insomnia (*n* = 29; ISI > 14) along three different dimensions: valence (1=very negative to 9=very positive, with 5=neutral), sleep-relatedness (1=not sleep-related to 9=very sleep-related, with 5=ambivalent), and arousal intensity (1=relaxed to 9=aroused, with 5=neutral/ambivalent). These participants were compensated 12 dollars per hour.

#### **fMRIPrep pre-processing steps**

Results included in this manuscript come from preprocessing performed using fMRIPrep 23.2.0 (Esteban et al. (2019); Esteban et al. (2018); RRID:SCR_016216), which is based on Nipype 1.8.6 (K. Gorgolewski et al. (2011); K. J. Gorgolewski et al. (2018); RRID:SCR_002502).

##### *Anatomical data preprocessing*

The T1w image was corrected for intensity non-uniformity (INU) with N4BiasFieldCorrection (Tustison et al. 2010), distributed with ANTs 2.5.0 (Avants et al. 2008, RRID:SCR_004757), and used as T1w-reference throughout the workflow. The T1w-reference was then skull-stripped with a Nipype implementation of the antsBrainExtraction.sh workflow (from ANTs), using OASIS30ANTs as target template. Brain tissue segmentation of cerebrospinal fluid (CSF), white-matter (WM) and gray-matter (GM) was performed on the brain-extracted T1w using fast (FSL (version unknown), RRID:SCR_002823, Zhang, Brady, and Smith 2001). Volume-based spatial normalization to two standard spaces (MNI152NLin6Asym, MNI152NLin2009cAsym) was performed through nonlinear registration with antsRegistration (ANTs 2.5.0), using brain-extracted versions of both T1w reference and the T1w template. The following templates were were selected for spatial normalization and accessed with TemplateFlow (23.1.0, Ciric et al. 2022): FSL’s MNI ICBM 152 non-linear 6th Generation Asymmetric Average Brain Stereotaxic Registration Model [Evans et al. (2012), RRID:SCR_002823; TemplateFlow ID: MNI152NLin6Asym], ICBM 152 Nonlinear Asymmetrical template version 2009c [Fonov et al. (2009), RRID:SCR_008796; TemplateFlow ID: MNI152NLin2009cAsym].

##### *Functional data preprocessing*

For each of the BOLD runs found per subject (across all tasks and sessions), the following preprocessing was performed. First, a reference volume was generated, using a custom methodology of fMRIPrep, for use in head motion correction. Head-motion parameters with respect to the BOLD reference (transformation matrices, and six corresponding rotation and translation parameters) are estimated before any spatiotemporal filtering using mcflirt (FSL, Jenkinson et al. 2002). The BOLD reference was then co-registered to the T1w reference using mri_coreg (FreeSurfer) followed by flirt (FSL, Jenkinson and Smith 2001) with the boundary-based registration (Greve and Fischl 2009) cost-function. Co-registration was configured with six degrees of freedom. Several confounding time-series were calculated based on the preprocessed BOLD: framewise displacement (FD), DVARS and three region-wise global signals. FD was computed using two formulations following Power (absolute sum of relative motions, Power et al. (2014)) and Jenkinson (relative root mean square displacement between affines, Jenkinson et al. (2002)). FD and DVARS are calculated for each functional run, both using their implementations in Nipype (following the definitions by Power et al. 2014). The three global signals are extracted within the CSF, the WM, and the whole-brain masks. Additionally, a set of physiological regressors were extracted to allow for component-based noise correction (CompCor, Behzadi et al. 2007). Principal components are estimated after high-pass filtering the preprocessed BOLD time-series (using a discrete cosine filter with 128s cut-off) for the two CompCor variants: temporal (tCompCor) and anatomical (aCompCor). tCompCor components are then calculated from the top 2% variable voxels within the brain mask. For aCompCor, three probabilistic masks (CSF, WM and combined CSF+WM) are generated in anatomical space. The implementation differs from that of Behzadi et al. in that instead of eroding the masks by 2 pixels on BOLD space, a mask of pixels that likely contain a volume fraction of GM is subtracted from the aCompCor masks. This mask is obtained by thresholding the corresponding partial volume map at 0.05, and it ensures components are not extracted from voxels containing a minimal fraction of GM. Finally, these masks are resampled into BOLD space and binarized by thresholding at 0.99 (as in the original implementation). Components are also calculated separately within the WM and CSF masks. For each CompCor decomposition, the k components with the largest singular values are retained, such that the retained components’ time series are sufficient to explain 50 percent of variance across the nuisance mask (CSF, WM, combined, or temporal). The remaining components are dropped from consideration. The head-motion estimates calculated in the correction step were also placed within the corresponding confounds file. The confound time series derived from head motion estimates and global signals were expanded with the inclusion of temporal derivatives and quadratic terms for each (Satterthwaite et al. 2013). Frames that exceeded a threshold of 0.5 mm FD or 1.5 standardized DVARS were annotated as motion outliers. Additional nuisance timeseries are calculated by means of principal components analysis of the signal found within a thin band (crown) of voxels around the edge of the brain, as proposed by (Patriat, Reynolds, and Birn 2017). All resamplings can be performed with a single interpolation step by composing all the pertinent transformations (i.e. head-motion transform matrices, susceptibility distortion correction when available, and co-registrations to anatomical and output spaces). Gridded (volumetric) resamplings were performed using nitransforms, configured with cubic B-spline interpolation.

Many internal operations of fMRIPrep use Nilearn 0.10.2 (Abraham et al. 2014, RRID:SCR_001362), mostly within the functional processing workflow. For more details of the pipeline, see the section corresponding to workflows in fMRIPrep’s documentation.

##### *Copyright Waiver*

The above boilerplate text was automatically generated by fMRIPrep with the express intention that users should copy and paste this text into their manuscripts unchanged. It is released under the CC0 license.

##### *References for fMRIPrep*

Abraham, Alexandre, Fabian Pedregosa, Michael Eickenberg, Philippe Gervais, Andreas Mueller, Jean Kossaifi, Alexandre Gramfort, Bertrand Thirion, and Gael Varoquaux. 2014. “Machine Learning for Neuroimaging with Scikit-Learn.” Frontiers in Neuroinformatics 8. https://doi.org/10.3389/fninf.2014.00014.

Avants, B. B., C. L. Epstein, M. Grossman, and J. C. Gee. 2008. “Symmetric Diffeomorphic Image Registration with Cross-Correlation: Evaluating Automated Labeling of Elderly and Neurodegenerative Brain.” Medical Image Analysis 12 (1): 26–41. https://doi.org/10.1016/j.media.2007.06.004.

Behzadi, Yashar, Khaled Restom, Joy Liau, and Thomas T. Liu. 2007. “A Component Based Noise Correction Method (CompCor) for BOLD and Perfusion Based fMRI.” NeuroImage 37 (1): 90–101. https://doi.org/10.1016/j.neuroimage.2007.04.042.

Ciric, R., William H. Thompson, R. Lorenz, M. Goncalves, E. MacNicol, C. J. Markiewicz, Y. O. Halchenko, et al. 2022. “TemplateFlow: FAIR-Sharing of Multi-Scale, Multi-Species Brain Models.” Nature Methods 19: 1568–71. https://doi.org/10.1038/s41592-022-01681-2.

Esteban, Oscar, Ross Blair, Christopher J. Markiewicz, Shoshana L. Berleant, Craig Moodie, Feilong Ma, Ayse Ilkay Isik, et al. 2018. “fMRIPrep 23.2.0.” Software. https://doi.org/10.5281/zenodo.852659.

Esteban, Oscar, Christopher Markiewicz, Ross W Blair, Craig Moodie, Ayse Ilkay Isik, Asier Erramuzpe Aliaga, James Kent, et al. 2019. “fMRIPrep: A Robust Preprocessing Pipeline for Functional MRI.” Nature Methods 16: 111–16. https://doi.org/10.1038/s41592-018-0235-4.

Evans, AC, AL Janke, DL Collins, and S Baillet. 2012. “Brain Templates and Atlases.” NeuroImage 62 (2): 911–22. https://doi.org/10.1016/j.neuroimage.2012.01.024.

Fonov, VS, AC Evans, RC McKinstry, CR Almli, and DL Collins. 2009. “Unbiased Nonlinear Average Age-Appropriate Brain Templates from Birth to Adulthood.” NeuroImage 47, Supplement 1: S102. https://doi.org/10.1016/S1053-8119(09)70884-5.

Gorgolewski, K., C. D. Burns, C. Madison, D. Clark, Y. O. Halchenko, M. L. Waskom, and S. Ghosh. 2011. “Nipype: A Flexible, Lightweight and Extensible Neuroimaging Data Processing Framework in Python.” Frontiers in Neuroinformatics 5: 13. https://doi.org/10.3389/fninf.2011.00013.

Gorgolewski, Krzysztof J., Oscar Esteban, Christopher J. Markiewicz, Erik Ziegler, David Gage Ellis, Michael Philipp Notter, Dorota Jarecka, et al. 2018. “Nipype.” Software. https://doi.org/10.5281/zenodo.596855.

Greve, Douglas N, and Bruce Fischl. 2009. “Accurate and Robust Brain Image Alignment Using Boundary-Based Registration.” NeuroImage 48 (1): 63–72. https://doi.org/10.1016/j.neuroimage.2009.06.060.

Jenkinson, Mark, Peter Bannister, Michael Brady, and Stephen Smith. 2002. “Improved Optimization for the Robust and Accurate Linear Registration and Motion Correction of Brain Images.” NeuroImage 17 (2): 825–41. https://doi.org/10.1006/nimg.2002.1132.

Jenkinson, Mark, and Stephen Smith. 2001. “A Global Optimisation Method for Robust Affine Registration of Brain Images.” Medical Image Analysis 5 (2): 143–56. https://doi.org/10.1016/S1361-8415(01)00036-6.

Patriat, Rémi, Richard C. Reynolds, and Rasmus M. Birn. 2017. “An Improved Model of Motion-Related Signal Changes in fMRI.” NeuroImage 144, Part A (January): 74–82. https://doi.org/10.1016/j.neuroimage.2016.08.051.

Power, Jonathan D., Anish Mitra, Timothy O. Laumann, Abraham Z. Snyder, Bradley L. Schlaggar, and Steven E. Petersen. 2014. “Methods to Detect, Characterize, and Remove Motion Artifact in Resting State fMRI.” NeuroImage 84 (Supplement C): 320–41. https://doi.org/10.1016/j.neuroimage.2013.08.048.

Satterthwaite, Theodore D., Mark A. Elliott, Raphael T. Gerraty, Kosha Ruparel, James Loughead, Monica E. Calkins, Simon B. Eickhoff, et al. 2013. “An improved framework for confound regression and filtering for control of motion artifact in the preprocessing of resting-state functional connectivity data.” NeuroImage 64 (1): 240–56. https://doi.org/10.1016/j.neuroimage.2012.08.052.

Tustison, N. J., B. B. Avants, P. A. Cook, Y. Zheng, A. Egan, P. A. Yushkevich, and J. C. Gee. 2010. “N4ITK: Improved N3 Bias Correction.” IEEE Transactions on Medical Imaging 29 (6): 1310–20. https://doi.org/10.1109/TMI.2010.2046908.

Zhang, Y., M. Brady, and S. Smith. 2001. “Segmentation of Brain MR Images Through a Hidden Markov Random Field Model and the Expectation-Maximization Algorithm.” IEEE Transactions on Medical Imaging 20 (1): 45–57. https://doi.org/10.1109/42.906424.

#### **Descriptive analyses**

Descriptive analyses were performed to examine the correlation between dysfunctional beliefs and attitudes about sleep using the DBAS-16 and valence ratings of emotional images across and between groups at V1. The relation between insomnia severity using the ISI and valence ratings across groups at V1 was also assessed. Additionally, the relation between subjective cognitive complaints, using the SASCI-Q, and performance on the Sternberg and face-name tasks across and between groups at V1 was also examined. To identify brain regions that were activated during each MRI task, we performed a one-sample *t*-test with age and sex as covariates. Additional descriptive analyses consisted of correlations between brain activity and performance on each MRI task across groups at V1. Additional descriptive analyses were performed to assess the correlation between the change in total DBAS-16, ISI, and SASCI-Q scores and change in task performance on the memory and emotion tasks. To correct for multiple comparisons, *p*-values were adjusted using the Benjamini-Hochberg procedure with an FDR set at .05.

### **Supplementary Results**

#### *Descriptive results*

While adjusting for multiple comparisons, we found a statistically significant positive correlation between total DBAS-16 scores and valence ratings for neutral (*r*(73) = .29, *p* = .02), negative (*r*(73) = .29, *p* = .02), and sleep-related (*r*(73) = .24, *p* = .04) images across INS and GS groups at V1. However, while adjusting for multiple comparisons, these correlations between total DBAS-16 scores and valence ratings for all images were no longer statistically significant for each group at V1 (all *p* > .05). Additionally, there was a statistically significant positive correlation between total ISI scores and valence ratings for neutral (*r*(73) = .38, *p* = .002), negative (*r*(73) = .25, *p* = .03), and sleep-related (*r*(73) = .37, *p* = .002) images across INS and GS groups at V1. Multiple comparison adjustment revealed there was only a significant positive correlation between ISI and valence ratings for neutral (*r*(54) = .40, *p* = .01) and negative (*r*(54) = .39, *p* = .01) images for the INS group at V1. Lastly, subjective complaints of memory were not correlated with performance on both the Sternberg and face-name tasks across and between groups at V1 (all *p* > .05).

We identified brain regions that were activated during each MRI task among all participants. After adjusting for multiple comparisons, the bilateral lingual gyri and right anterior insula were significantly activated during the Sternberg task (*p* < .05). The emotion reactivity task led to activation in the bilateral angular gyri, left precuneus, left middle frontal gyrus, and bilateral middle temporal gyri (*p* < .05). Lastly, the left thalamus proper, hippocampus, middle frontal gyrus, posterior cingulate gyrus, precuneus, angular gyrus as well as the right middle temporal gyrus, calcarine cortex, transverse temporal gyrus, and cuneus were found to be activated during the face-name task (*p* < .05). However, with or without correcting for multiple comparisons, there was no statistically significant correlation between task performance and activity in the corresponding brain regions.

Changes in total DBAS-16 scores and ISI scores did not significantly correlate with changes in valence ratings for neutral, negative, or sleep-related images in the TX group, WL group, or combined groups (all *p* > .05). Similarly, no statistically significant correlations between change in total SASCI-Q scores and change in performance on either memory task were observed in the TX group, WL group, or combined groups (all *p* > .05).

#### **Table S1.** Self-reported sleep, emotion, and cognition measures and cognitive performance of INS and GS groups at baseline

|  | *INS* | |  | *GS* | | *Group comparison (INS vs. GS)* | | |
| --- | --- | --- | --- | --- | --- | --- | --- | --- |
| Outcome | *n* | *M* (*SD*) |  | *n* | *M* (*SD*) | *t* (*df*) | *p* | $\beta$ [95% CI] |
| **Sleep** | | | | | | | | |
| ISI | 56 | 16.34 (3.78) |  | 20 | 2.85 (2.64) | 14.12 (72) | < .001* | 1.89 [1.63, 2.16] |
| PSQI | 55 | 9.64 (2.72) |  | 20 | 3.05 (1.47) | 9.76 (72) | < .001* | 1.69 [1.35, 2.04] |
| DBAS-16 | 56 | 5.07 (1.54) |  | 20 | 3.84 (1.88) | 2.74 (72) | .01* | 0.72 [0.20, 1.24] |
| DISRS | 56 | 41.48 (10.96) |  | 20 | 32.80 (13.81) | 2.89 (72) | .01* | 0.73 [0.23, 1.23] |
| TST (min.) | 48 | 386.45 (62.64) |  | 19 | 434.12 (34.96) | -2.55 (64) | .01* | -0.63 [-1.13, -0.14] |
| SOL (min.) | 49 | 29.12 (18.51) |  | 19 | 15.06 (9.77) | 3.33 (65) | < .001* | 0.85 [0.34, 1.36] |
| WASO (min.) | 49 | 45.48 (28.87) |  | 19 | 11.50 (6.17) | 4.64 (65) | < .001* | 1.10 [0.62, 1.57] |
| Number of awakenings | 49 | 2.33 (1.62) |  | 19 | 0.92 (0.84) | 3.35 (65) | < .001* | 0.85 [0.34, 1.36] |
| SE (%) | 48 | 77.13 (10.15) |  | 19 | 89.76 (5.83) | -4.79 (64) | < .001* | -1.10 [-1.56, -0.64] |
| **Emotions** | | | | | | | | |
| BDI-II | 56 | 10.91 (7.42) |  | 19 | 4.58 (3.81) | 3.76 (71) | .001* | 0.91 [0.43, 1.40] |
| PANAS – positive affect | 55 | 31.44 (7.47) |  | 20 | 35.45 (7.55) | -2.43 (71) | .02* | -0.63 [-1.15, -0.11] |
| PANAS – negative affect | 55 | 18.42 (5.89) |  | 20 | 15.85 (4.83) | 2.30 (71) | .02* | 0.58 [0.08, 1.09] |
| STICSA – total | 56 | 34.14 (9.72) |  | 20 | 27.80 (5.04) | 3.15 (72) | .01* | 0.77 [0.28, 1.26] |
| STICSA – cognitive | 56 | 18.48 (6.17) |  | 20 | 14.60 (4.01) | 2.90 (72) | .01* | 0.73 [0.23, 1.23] |
| STICSA – somatic | 56 | 15.66 (4.53) |  | 20 | 13.20 (1.70) | 2.68 (72) | .01* | 0.67 [0.17, 1.16] |
| **Cognition** | | | | | | | | |
| SASCI-Q | 55 | 83.33 (27.01) |  | 20 | 65.90 (21.49) | 2.47 (71) | .02* | 0.65 [0.12, 1.17] |
| **Cognitive performance** | | | | | | | | |
| **Sternberg working memory task** | | | | | | | | |
| Accuracy (% correct) | 56 | 93.50 (6.61) |  | 19 | 92.32 (9.89) | 0.89 (70) | .96 | 0.25 [-0.31, 0.82] |
| **Emotion reactivity task** | | | | | | | | |
| Valence rating of neutral images | 56 | 1.74 (0.48) |  | 19 | 1.50 (0.37) | 1.53 (71) | .19 | 0.40 [-0.12, 0.91] |
| Valence rating of negative images | 56 | 3.46 (0.70) |  | 19 | 3.35 (0.51) | 0.29 (71) | .77 | 0.08 [-0.46, 0.61] |
| Valence rating of sleep images | 56 | 2.76 (0.77) |  | 19 | 2.19 (0.59) | 2.69 (71) | .03* | 0.70 [0.18, 1.23] |
| **Face-name associative memory task** | | | | | | | | |
| **Immediate recall (50 stimuli)** | | | | | | | | |
| Number of hits | 40 | 26.10 (5.25) |  | 18 | 26.17 (7.56) | 0.06 (54) | .95 | 0.02 [-0.55, 0.59] |
| Accuracy (Hit+CR - Miss+FA) | 40 | 22.15 (11.79) |  | 18 | 20.83 (15.82) | 0.10 (54) | .95 | 0.03 [-0.53, 0.58] |
| Recognition (Hit-FA) | 40 | 24.58 (6.17) |  | 18 | 24.61 (8.08) | -0.07 (54) | .95 | -0.02 [-0.58, 0.54] |
| **Delayed recall (50 stimuli)** | | | | | | | | |
| Number of hits | 40 | 28.15 (5.80) |  | 18 | 27.22 (7.51) | 0.27 (54) | .95 | 0.08 [-0.49, 0.64] |
| Accuracy (Hit+CR - Miss+FA) | 40 | 24.25 (13.23) |  | 18 | 21.56 (15.49) | 0.34 (54) | .95 | 0.09 [-0.46, 0.65] |
| Recognition (Hit-FA) | 40 | 26.53 (6.79) |  | 18 | 25.67 (7.73) | 0.13 (54) | .95 | 0.03 [-0.52, 0.59] |
| **Delayed recall (100 stimuli)** | | | | | | | | |
| Number of hits | 40 | 47.05 (12.08) |  | 18 | 45.22 (16.37) | 0.43 (54) | .98 | 0.12 [-0.46, 0.70] |
| Accuracy (Hit+CR - Miss+FA) | 40 | 30.78 (26.15) |  | 18 | 25.72 (33.01) | 0.44 (54) | .98 | 0.12 [-0.45, 0.70] |
| Recognition (Hit-FA) | 40 | 43.60 (13.20) |  | 18 | 42.11 (16.52) | 0.26 (54) | .98 | 0.07 [-0.50, 0.65] |
| **Delayed recall (50 stimuli) – Immediate recall (50 stimuli)** | | | | | | | | |
| Change in number of hits | 40 | 2.08 (4.35) |  | 18 | 1.39 (2.12) | 0.06 (54) | .96 | 0.02 [-0.58, 0.61] |
| Change in accuracy (Hit+CR - Miss+FA) | 40 | 2.23 (8.79) |  | 18 | 1.22 (5.50) | 0.23 (54) | .96 | 0.07 [-0.54, 0.68] |
| Change in recognition (Hit-FA) | 40 | 1.98 (4.90) |  | 18 | 1.39 (2.81) | 0.05 (54) | .96 | 0.02 [-0.59, 0.62] |

*Note*. **p* < .05; Standardized beta coefficients ($\beta$) are reported with 95% confidence intervals ([95% CI]). BDI-II: Beck Depression Inventory-II; CR: Correct Rejection; DBAS-16: Dysfunctional Beliefs and Attitudes about Sleep; DISRS: Daytime Insomnia Symptom Response Scale; FA: False Alarm; GS: Good sleeper group; INS: Insomnia group; ISI: Insomnia Severity Index; PANAS: Positive Affect Negative Affect Scale; PSQI: Pittsburgh Sleep Quality Index; SASCI-Q: Sahlgrenska Academy Self-reported Cognitive Impairment questionnaire; SE: Sleep Efficiency; SOL: Sleep Onset Latency; STICSA: State-Trait Inventory for Cognitive and Somatic Anxiety; TST: Total Sleep Time; WASO: Wake After Sleep Onset; TST, and WASO are reported in minutes (min.).

#### **Table S2.** Results for high- vs. low-reward face-name pairs at baseline

|  | *INS* | |  | *GS* | | *Group comparison (INS vs. GS)* | | |
| --- | --- | --- | --- | --- | --- | --- | --- | --- |
| Outcome | *n* | *M* (*SD*) |  | *n* | *M* (*SD*) | *t* (*df*) | *p* | $\beta$ [95% CI] |
| **High reward** | | | | | | | | |
| **Immediate recall (50 stimuli)** | | | | | | | | |
| Number of hits | 40 | 12.42 (3.90) |  | 18 | 13.00 (3.90) | -0.03 (54) | .98 | -0.01 [-0.58, 0.57] |
| Accuracy (Hit+CR - Miss+FA) | 40 | 10.94 (9.73) |  | 18 | 13.61 (9.02) | -0.25 (54) | .96 | -0.07 [-0.63, 0.49] |
| Recognition (Hit-FA) | 40 | 10.09 (5.00) |  | 18 | 11.44 (4.59) | -0.22 (54) | .96 | -0.06 [-0.62, 0.50] |
| **Delayed recall (50 stimuli)** | | | | | | | | |
| Number of hits | 40 | 13.18 (4.03) |  | 18 | 13.06 (4.56) | 0.84 (54) | .96 | 0.23 [-0.33, 0.80] |
| Accuracy (Hit+CR - Miss+FA) | 40 | 11.79 (10.53) |  | 18 | 12.72 (10.11) | 0.75 (54) | .96 | 0.21 [-0.34, 0.75] |
| Recognition (Hit-FA) | 40 | 10.67 (5.34) |  | 18 | 11.50 (5.07) | 0.50 (54) | .96 | 0.14 [-0.42, 0.69] |
| **Delayed recall (100 stimuli)** | | | | | | | | |
| Number of hits | 40 | 21.91 (7.20) |  | 18 | 22.50 (8.96) | 0.62 (54) | .96 | 0.18 [-0.39, 0.75] |
| Accuracy (Hit+CR - Miss+FA) | 40 | 15.36 (17.14) |  | 18 | 18.72 (19.06) | 0.53 (54) | .96 | 0.15 [-0.41, 0.70] |
| Recognition (Hit-FA) | 40 | 17.00 (8.35) |  | 18 | 19.39 (9.54) | 0.29 (54) | .96 | 0.08 [-0.48, 0.64] |
| **Delayed recall (50 stimuli) – Immediate recall (50 stimuli)** | | | | | | | | |
| Change in number of hits | 40 | 0.76 (2.93) |  | 18 | 0.06 (1.92) | 1.26 (54) | .96 | 0.36 [-0.21, 0.92] |
| Change in accuracy (Hit+CR - Miss+FA) | 40 | 0.85 (6.82) |  | 18 | -0.89 (5.05) | 1.32 (54) | .96 | 0.39 [-0.20, 0.99] |
| Change in recognition (Hit-FA) | 40 | 0.58 (3.63) |  | 18 | 0.06 (2.44) | 0.94 (54) | .96 | 0.28 [-0.32, 0.87] |
| **Low reward** | | | | | | | | |
| **Immediate recall (50 stimuli)** | | | | | | | | |
| Number of hits | 40 | 12.30 (3.42) |  | 18 | 13.17 (4.06) | 0.14 (54) | .96 | 0.04 [-0.54, 0.63] |
| Accuracy (Hit+CR - Miss+FA) | 40 | 10.45 (9.76) |  | 18 | 13.72 (9.37) | -0.10 (54) | .96 | -0.03 [-0.59, 0.54] |
| Recognition (Hit-FA) | 40 | 9.97 (5.16) |  | 18 | 11.61 (4.73) | -0.10 (54) | .96 | -0.03 [-0.60, 0.54] |
| **Delayed recall (50 stimuli)** | | | | | | | | |
| Number of hits | 40 | 12.67 (3.77) |  | 18 | 14.17 (3.73) | -0.42 (54) | .96 | -0.12 [-0.70, 0.46] |
| Accuracy (Hit+CR - Miss+FA) | 40 | 10.88 (10.06) |  | 18 | 15.33 (7.83) | -0.44 (54) | .96 | -0.12 [-0.69, 0.44] |
| Recognition (Hit-FA) | 40 | 10.15 (5.10) |  | 18 | 12.61 (3.91) | -0.54 (54) | .96 | -0.15 [-0.72, 0.41] |
| **Delayed recall (100 stimuli)** | | | | | | | | |
| Number of hits | 40 | 21.06 (7.02) |  | 18 | 22.72 (8.19) | 0.17 (54) | .96 | 0.05 [-0.54, 0.65] |
| Accuracy (Hit+CR - Miss+FA) | 40 | 13.94 (16.70) |  | 18 | 20.22 (16.81) | -0.10 (54) | .96 | -0.03 [-0.61, 0.55] |
| Recognition (Hit-FA) | 40 | 16.15 (8.33) |  | 18 | 19.61 (8.42) | -0.12 (54) | .96 | -0.04 [-0.62, 0.55] |
| **Delayed recall (50 stimuli) – Immediate recall (50 stimuli)** | | | | | | | | |
| Change in number of hits | 40 | 0.36 (2.30) |  | 18 | 1.00 (1.85) | -0.72 (54) | .96 | -0.22 [-0.83, 0.39] |
| Change in accuracy (Hit+CR - Miss+FA) | 40 | 0.42 (6.35) |  | 18 | 1.61 (6.20) | -0.41 (54) | .96 | -0.12 [-0.74, 0.49] |
| Change in recognition (Hit-FA) | 40 | 0.18 (3.42) |  | 18 | 1.00 (3.12) | -0.50 (54) | .96 | -0.15 [-0.77, 0.46] |

*Note*. **p* < .05; Standardized beta coefficients ($\beta$) are reported with 95% confidence intervals ([95% CI]). CR: Correct Rejection; FA: False Alarm; Good sleeper group; INS: Insomnia group.

#### **Table S3.** Brain activity during the Sternberg working memory, emotion reactivity, and face-name associative memory tasks at V1 and V2

|  |  |  |  |  |  | Cluster-level | |  | Peak-level | | |
| --- | --- | --- | --- | --- | --- | --- | --- | --- | --- | --- | --- |
| Visit | Group | Contrast | Brain region  (BA) | Coordinates  (x, y, z) |  | Cluster size (voxels) | *p_FWE-corrected_* |  | *t* | *z_E_* | *p_FWE-corrected_* |
|  | **Sternberg working memory task** | | | | | | | | | | |
| V1 | GS | Correct encoding |  |  |  |  |  |  |  |  |  |
|  |  |  | Left anterior insula (BA13) | (-34, 20, 4) |  | 212 | .000 |  | 7.81 | 6.61 | .000 |
|  |  |  | Right anterior insula (BA13) | (34, 26, 0) |  | 254 | .000 |  | 6.76 | 5.92 | .000 |
|  |  |  | Right superior frontal gyrus (BA6) | (2, 18, 48) |  | 75 | .001 |  | 5.24 | 4.80 | .0013 |
| V1 | INS | Correct encoding |  |  |  |  |  |  |  |  |  |
|  |  |  | Left anterior insula (BA13) | (-34, 20, 4) |  | 639 | .000 |  | 12.69 | Inf | .000 |
|  |  |  | Right anterior insula (BA13) | (34, 26, -4) |  | 475 | .000 |  | 9.79 | 7.76 | .000 |
|  |  |  | Right lingual gyrus (BA18) | (12, -74, 0) |  | 561 | .000 |  | 7.83 | 6.63 | .000 |
|  |  |  | Right cuneus (BA17) | (14, -84, 18) |  |  |  |  | 6.46 | 5.71 | .000 |
|  |  |  | Right lingual gyrus (BA19) | (26, -68, 4) |  |  |  |  | 5.66 | 5.13 | .003 |
|  |  |  | Left medial frontal gyrus (BA32) | (-6, 18, 44) |  | 640 | .000 |  | 7.29 | 6.28 | .000 |
|  |  |  | Right medial frontal gyrus (BA8) | (6, 32, 36) |  |  |  |  | 6.02 | 5.40 | .001 |
|  |  |  | Right cingulate gyrus (BA32) | (10, 24, 30) |  |  |  |  | 5.63 | 5.10 | .003 |
|  |  |  | Left lingual gyrus (BA18) | (-16, -76, 2) |  | 447 | .000 |  | 7.26 | 6.25 | .000 |
|  |  |  | Left lingual gyrus (BA19) | (-28, -64, 2) |  |  |  |  | 4.88 | 4.52 | .042 |
|  |  |  | Left precentral gyrus (BA6) | (-38, -4, 50) |  | 417 | .000 |  | 6.79 | 5.94 | .000 |
|  |  |  | Left precentral gyrus (BA6) | (-60, 10, 30) |  |  |  |  | 6.18 | 5.51 | .000 |
|  |  |  | Left precentral gyrus (BA6) | (-46, 2, 34) |  |  |  |  | 5.64 | 5.11 | .003 |
|  |  |  | Right middle frontal gyrus (BA9) | (38, 38, 20) |  | 136 | .000 |  | 6.47 | 5.72 | .000 |
|  |  |  | Right parahippocampus (BA19) | (34, -48, -2) |  | 6 | .024 |  | 5.31 | 4.86 | .010 |
|  |  |  | Left angular gyrus (BA39) | (-28, -50, 40) |  | 7 | .022 |  | 4.98 | 4.60 | .030 |
|  |  |  | Left cuneus (BA17) | (-10, -86, 16) |  | 2 | .035 |  | 4.90 | 4.53 | .0039 |
| V1 | TX | Correct encoding |  |  |  |  |  |  |  |  |  |
|  |  |  | Left anterior insula (BA13) | (-34, 20, 2) |  | 540 | .000 |  | 10.99 | Inf | .000 |
|  |  |  | Right anterior insula (BA13) | (32, 24, -2) |  | 375 | .000 |  | 9.35 | Inf | .000 |
|  |  |  | Left precentral gyrus (BA6) | (-38, -4, 50) |  | 231 | .000 |  | 7.32 | 6.51 | .000 |
|  |  |  | Left precentral gyrus (BA6) | (-42, 0, 36) |  |  |  |  | 5.31 | 4.96 | .006 |
|  |  |  | Right lingual gyrus (BA18) | (10, -74, 2) |  | 549 | .000 |  | 7.26 | 6.47 | .000 |
|  |  |  | Right lingual gyrus (BA19) | (28, -62, 4) |  |  |  |  | 5.15 | 4.82 | .012 |
|  |  |  | Left medial frontal gyrus (BA32) | (-6, 18, 42) |  | 415 | .000 |  | 6.71 | 6.06 | .000 |
|  |  |  | Right medial frontal gyrus (BA32) | (8, 14, 44) |  |  |  |  | 5.79 | 5.35 | .001 |
|  |  |  | Right middle frontal gyrus (BA9) | (38, 36, 18) |  | 72 | .001 |  | 6.40 | 5.83 | .000 |
|  |  |  | Left lingual gyrus (BA18) | (-12, -72, 4) |  | 217 | .000 |  | 6.09 | 5.59 | .000 |
|  |  |  | Right cingulate gyrus (BA32) | (10, 26, 28) |  | 7 | .0.022 |  | 5.00 | 4.70 | .020 |
| V1 | WL | Correct encoding |  |  |  |  |  |  |  |  |  |
|  |  |  | Left anterior insula (BA13) | (-34, 20, 4) |  | 196 | .000 |  | 7.51 | 6.64 | .000 |
|  |  |  | Right anterior insula (BA13) | (32, 26, -2) |  | 137 | .000 |  | 6.17 | 5.65 | .000 |
| V2 | TX | Correct encoding |  |  |  |  |  |  |  |  |  |
|  |  |  | Left anterior insula (BA13) | (-32, 22, 0) |  | 433 | .000 |  | 9.56 | Inf | .000 |
|  |  |  | Right anterior insula (BA13) | (32, 24, 0) |  | 333 | .000 |  | 8.20 | 7.12 | .000 |
|  |  |  | Left cingulate gyrus (BA24) | (-10, 8, 48) |  | 7 | .022 |  | 4.90 | 4.61 | .029 |
| V2 | WL | Correct encoding |  |  |  |  |  |  |  |  |  |
|  |  |  | Left anterior insula (BA13) | (-32, 22, 0) |  | 269 | .000 |  | 8.91 | 7.59 | .000 |
|  |  |  | Right anterior insula (BA13) | (34, 24, -2) |  | 272 | .000 |  | 8.70 | 7.45 | .000 |
|  |  |  | Right medial frontal gyrus (BA8) | (6, 32, 34) |  | 363 | .000 |  | 6.01 | 5.52 | .000 |
|  |  |  | Left medial frontal gyrus (BA8) | (-4, 16, 48) |  |  |  |  | 5.81 | 5.37 | .001 |
|  |  |  | Right medial frontal gyrus (BA8) | (4, 24, 46) |  |  |  |  | 5.66 | 5.25 | .002 |
|  |  |  | Right middle frontal gyrus (BA10) | (38, 40, 16) |  | 142 | .000 |  | 5.78 | 5.34 | .001 |
|  |  |  | Left precentral gyrus (BA6) | (-44, 2, 50) |  | 25 | .007 |  | 5.10 | 4.78 | .014 |
|  |  |  | Left middle frontal gyrus (BA6) | (-36, 0, 58) |  |  |  |  | 4.92 | 4.64 | .026 |
|  |  |  | Right lingual gyrus (BA18) | (12, -74, 0) |  | 3 | .031 |  | 4.93 | 4.64 | .025 |
|  | **Emotion reactivity task** | | | | | | | | | | |
| V1 | GS | Sleep > Neutral |  |  |  |  |  |  |  |  |  |
|  |  |  | Left angular gyrus (BA39) | (-42, -60, 40) |  | 24 | .006 |  | 5.22 | 4.78 | .016 |
| V1 | INS | Sleep > Neutral |  |  |  |  |  |  |  |  |  |
|  |  |  | Left angular gyrus (BA39) | (-42, -60, 40) |  | 1016 | .000 |  | 9.23 | 7.46 | .000 |
|  |  |  | Right angular gyrus (BA39) | (46, -62, 44) |  | 430 | .000 |  | 7.49 | 6.41 | .000 |
|  |  |  | Right middle temporal gyrus (BA39) | (52, -60, 38) |  |  |  |  | 7.39 | 6.34 | .000 |
|  |  |  | Left precuneus (BA31) | (-12, -50, 30) |  | 720 | .000 |  | 7.40 | 6.35 | .000 |
|  |  |  | Left cingulate gyrus (BA31) | (-4, -38, 34) |  |  |  |  | 6.32 | 5.61 | .000 |
|  |  |  | Right precuneus (BA31) | (10, -46, 32) |  |  |  |  | 5.74 | 5.19 | .002 |
|  |  |  | Left middle frontal gyrus (BA8) | (-44, 22, 44) |  | 157 | .000 |  | 7.38 | 6.34 | .000 |
|  |  |  | Left middle frontal gyrus (BA46) | (-42, 50, 0) |  | 38 | .003 |  | 5.52 | 5.02 | .006 |
|  |  |  | Right cerebellum exterior | (40, -70, -46) |  | 1 | .039 |  | 5.03 | 4.64 | .028 |
|  |  |  | Left posterior cingulate gyrus (BA23) | (-4, -28, 22) |  | 1 | .039 |  | 4.93 | 4.56 | .037 |
|  |  |  | Left cingulate gyrus (BA23) | (-2, -24, 34) |  | 1 | .039 |  | 4.87 | 4.51 | .040 |
| V1 | TX | Sleep > Neutral |  |  |  |  |  |  |  |  |  |
|  |  |  | Left middle temporal gyrus (BA39) | (-42, -62, 32) |  | 379 | .000 |  | 6.88 | 6.18 | .000 |
|  |  |  | Left precentral gyrus (BA9) | (-42, 22, 40) |  | 57 | .002 |  | 5.57 | 5.17 | .002 |
|  |  |  | Left cingulate gyrus (BA31) | (-14, -46, 28) |  | 18 | .011 |  | 5.09 | 4.78 | .014 |
|  |  |  | Right angular gyrus (BA39) | (50, -62, 38) |  | 18 | .011 |  | 4.90 | 4.62 | .027 |
|  |  |  | Right precuneus (BA39) | (44, -66, 44) |  |  |  |  | 4.86 | 4.58 | .031 |
| V1 | WL | Sleep > Neutral |  |  |  |  |  |  |  |  |  |
|  |  |  | Left angular gyrus (BA39) | (-44, -58, 42) |  | 218 | .000 |  | 5.42 | 5.05 | .004 |
|  |  |  | Left middle temporal gyrus (BA39) | (-42, -58, 30) |  |  |  |  | 5.20 | 4.87 | .009 |
|  |  |  |  | (-12, -50, 30) |  | 6 | .024 |  | 4.88 | 4.60 | .029 |
| V2 | TX | Sleep > Neutral |  |  |  |  |  |  |  |  |  |
|  |  |  | Left inferior parietal lobule (BA40) | (-48, -60, 42) |  | 151 | .000 |  | 5.33 | 4.98 | .006 |
|  |  |  | Right angular gyrus (BA39) | (48, -60, 40) |  | 16 | .013 |  | 4.85 | 4.58 | .032 |
|  |  |  | Right inferior parietal lobule (BA40) | (54, -50, 46) |  | 1 | .040 |  | 4.79 | 4.52 | .040 |
| V2 | WL | Sleep > Neutral |  |  |  |  |  |  |  |  |  |
|  |  |  | Left middle temporal gyrus (BA39) | (-42, -60, 38) |  | 232 | .000 |  | 5.49 | 5.10 | .003 |
|  |  |  | Left posterior cingulate gyrus (BA23) | (-2, -26, 22) |  | 8 | .415 |  | 5.08 | 4.77 | .014 |
| V1 | GS | Negative > Neutral |  |  |  |  |  |  |  |  |  |
|  |  |  | Right lingual gyrus (BA18) | (20, -74, -10) |  | 1071 | .000 |  | 6.76 | 5.92 | .000 |
|  |  |  | Right lingual gyrus (BA18) | (12, -78, -8) |  |  |  |  | 6.33 | 5.61 | .000 |
|  |  |  | Right cuneus (BA17) | (18, -92, 14) |  |  |  |  | 6.03 | 5.40 | .001 |
|  |  |  | Left middle occipital gyrus (BA19) | (-30, -92, 16) |  | 28 | .006 |  | 5.92 | 5.32 | .001 |
|  |  |  | Left cerebellum exterior | (-38, -64, -20) |  | 49 | .002 |  | 5.65 | 5.11 | .003 |
|  |  |  | Left middle occipital gyrus (BA18) | (-42, -82, -4) |  | 4 | .028 |  | 4.98 | 4.60 | .031 |
| V1 | INS | Negative > Neutral |  |  |  |  |  |  |  |  |  |
|  |  |  | Right lingual gyrus (BA18) | (16, -78, -10) |  | 663 | .000 |  | 7.60 | 6.48 | .000 |
|  |  |  | Right lingual gyrus (BA18) | (8, -80, 0) |  |  |  |  | 6.17 | 5.50 | .000 |
|  |  |  | Right cuneus (BA17) | (22, -90, 12) |  | 1 | .040 |  | 4.95 | 4.57 | .034 |
| V1 | TX | Negative > Neutral | No suprathreshold clusters | | | | | | | | |
| V1 | WL | Negative > Neutral |  |  |  |  |  |  |  |  |  |
|  |  |  | Right lingual gyrus (BA18) | (16, -76, -12) |  | 276 | .000 |  | 6.32 | 5.77 | .000 |
|  |  |  | Right cerebellum exterior | (26, -76, -14) |  |  |  |  | 5.62 | 5.21 | .002 |
|  |  |  | Right lingual gyrus (BA18) | (22, -80, -4) |  |  |  |  | 5.24 | 4.90 | .007 |
|  |  |  | Right lingual gyrus (BA18) | (8, -80, 2) |  | 61 | .002 |  | 5.37 | 5.01 | .004 |
| V2 | TX | Negative > Neutral |  |  |  |  |  |  |  |  |  |
|  |  |  | Right occipital fusiform gyrus | (30, -76, -18) |  | 408 | .000 |  | 6.60 | 5.98 | .000 |
|  |  |  | Right lingual gyrus (BA18) | (8, -86, -4) |  |  |  |  | 4.87 | 4.60 | .026 |
|  |  |  | Right cerebellum exterior | (28, -48, -20) |  | 37 | .226 |  | 5.11 | 4.79 | .011 |
|  |  |  | Right cerebellum exterior | (30, -58, -18) |  |  |  |  | 4.90 | 4.62 | .024 |
|  |  |  | Left occipital fusiform gyrus | (-46, -68, -16) |  | 4 | .732 |  | 4.81 | 4.54 | .032 |
|  |  |  | Right middle occipital gyrus (BA18) | (36, -86, 8) |  | 2 | .732 |  | 4.77 | 4.50 | .037 |
| V2 | WL | Negative > Neutral |  |  |  |  |  |  |  |  |  |
|  |  |  | Right lingual gyrus (BA18) | (18, -78, -10) |  | 292 | .000 |  | 5.98 | 5.50 | .000 |
|  | **Face-name associative memory task** | | | | | | | | | | |
| V1 | GS | Hit + Miss > Control | No suprathreshold clusters | | | | | | | | |
| V1 | INS | Hit + Miss > Control |  |  |  |  |  |  |  |  |  |
|  |  |  | Left parahippocampus | (-28, -14, -14) |  | 43 | .003 |  | 6.12 | 5.31 | .001 |
| V1 | TX | Hit + Miss > Control | No suprathreshold clusters | | | | | | | | |
| V1 | WL | Hit + Miss > Control | No suprathreshold clusters | | | | | | | | |
| V2 | TX | Hit + Miss > Control | No suprathreshold clusters | | | | | | | | |
| V2 | WL | Hit + Miss > Control |  |  |  |  |  |  |  |  |  |
|  |  |  | Left occipital fusiform gyrus | (-34, -80, -12) |  | 2 | .036 |  | 4.97 | 4.52 | .041 |
| V1 | GS | Hit > Control |  |  |  |  |  |  |  |  |  |
|  |  |  | Left caudate | (-8, -6, 22) |  | 1 | .039 |  | 5.04 | 4.54 | .044 |
| V1 | INS | Hit > Control |  |  |  |  |  |  |  |  |  |
|  |  |  | Left parahippocampus | (-28, -14, -14) |  | 132 | .000 |  | 7.72 | 6.31 | .000 |
| V1 | TX | Hit > Control | No suprathreshold clusters | | | | | | | | |
| V1 | WL | Hit > Control | No suprathreshold clusters | | | | | | | | |
| V2 | TX | Hit > Control | No suprathreshold clusters | | | | | | | | |
| V2 | WL | Hit > Control | No suprathreshold clusters | | | | | | | | |

*Note*. Results are presented with *p_FWE-corrected_* set to .05 and extent threshold (voxels) set to 0. Coordinates are presented in mm. Good sleeper group; Inf: Infinity; INS: Insomnia group; TX: Immediate CBTi group; WL: Wait-list control group.

#### **Figure S2.** Within network resting-state functional connectivity differences between participants with insomnia and good sleepers at baseline

(A) Resting-state functional connectivity (rs-FC) differences within the default mode network (DMN)

(B) rs-FC differences within the DMN

(C) rs-FC differences within the salience network (SN)

(D) rs-FC differences within the SN

(E) rs-FC differences within the frontoparietal network (FPN)

(F) rs-FC differences within the FPN

Panels A, C, and E include participants with insomnia disorder vs. good sleepers who did not sleep during the resting-state task, whereas panels B, D, and F include all participants with insomnia disorder vs. good sleepers. Age and sex were included as covariates. As no connections survived following false discovery rate (FDR) correction set at .05, only uncorrected results are displayed.

**A**	**DMN (participants who did not fall asleep)	B**	**DMN (all participants)**

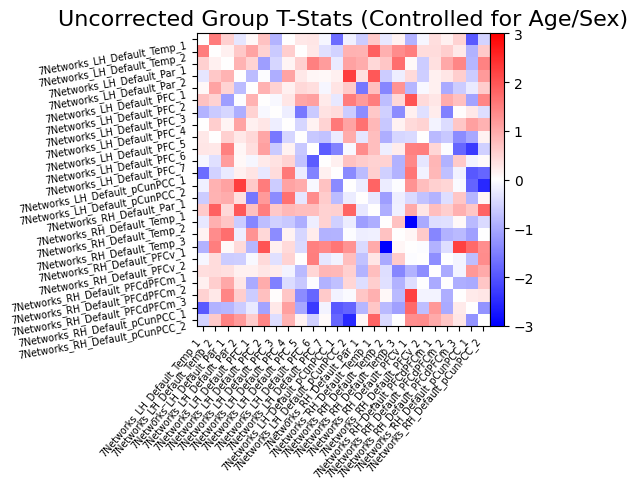

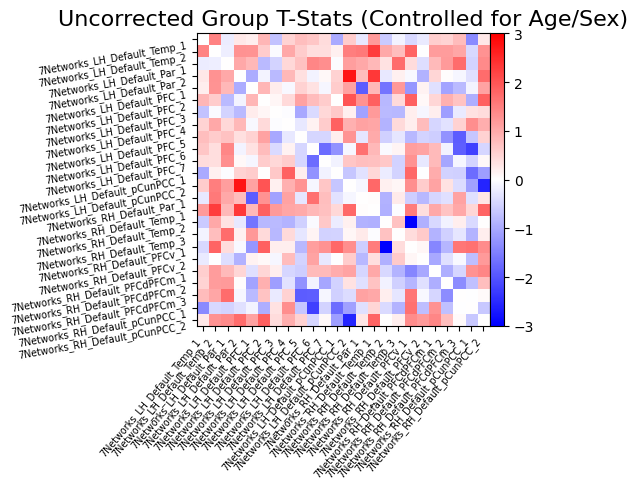

**C**	**SN (participants who did not fall asleep)	D**	**SN (all participants)**

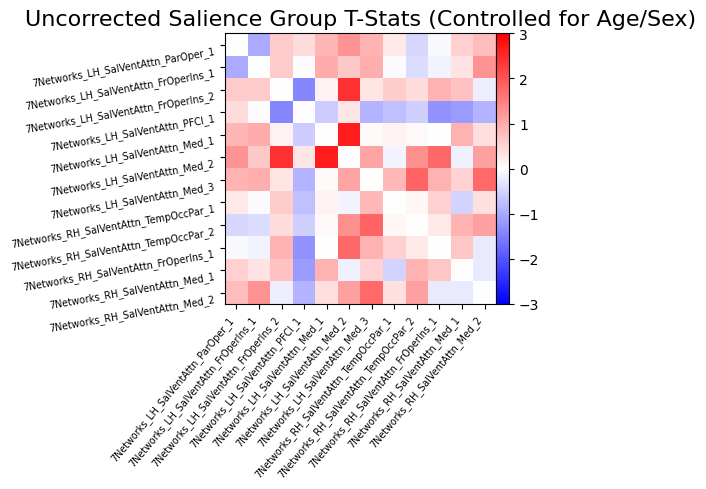

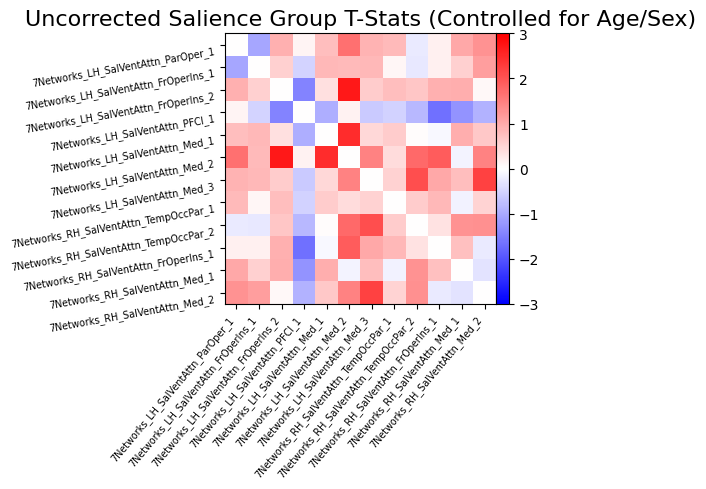

**E**	**FPN (participants who did not fall asleep)	F**	**FPN (all participants)**

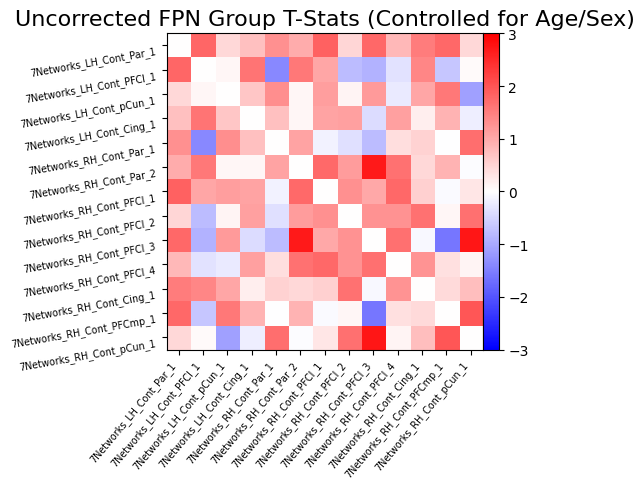

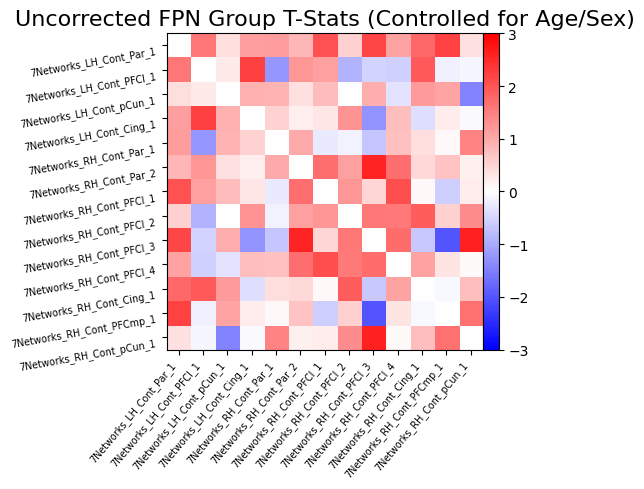

#### **Figure S3.** Between-network resting-state functional connectivity between participants with insomnia and good sleepers at baseline

(A) Default mode network (DMN) and salience network (SN) connectivity differences

(B) DMN-SN connectivity differences

(C) DMN and frontoparietal network (FPN) connectivity differences

(D) DMN-FPN connectivity differences

(E) SN-FPN connectivity differences

(F) SN-FPN connectivity differences

Panels A, C, and E include participants with insomnia disorder vs. good sleepers who did not sleep during the resting-state task, whereas panels B, D, and F include all participants with insomnia disorder vs. good sleepers. Age and sex were included as covariates. As no connections survived following false discovery rate (FDR) correction set at .05, only uncorrected results are displayed.

**A**	**DMN vs. SN (participants who did not fall asleep)	B**	**DMN vs. SN (all participants)**

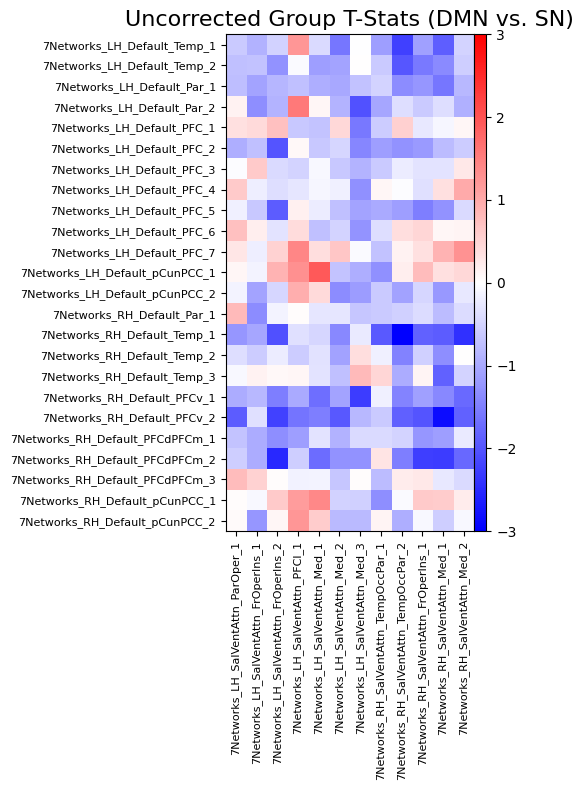

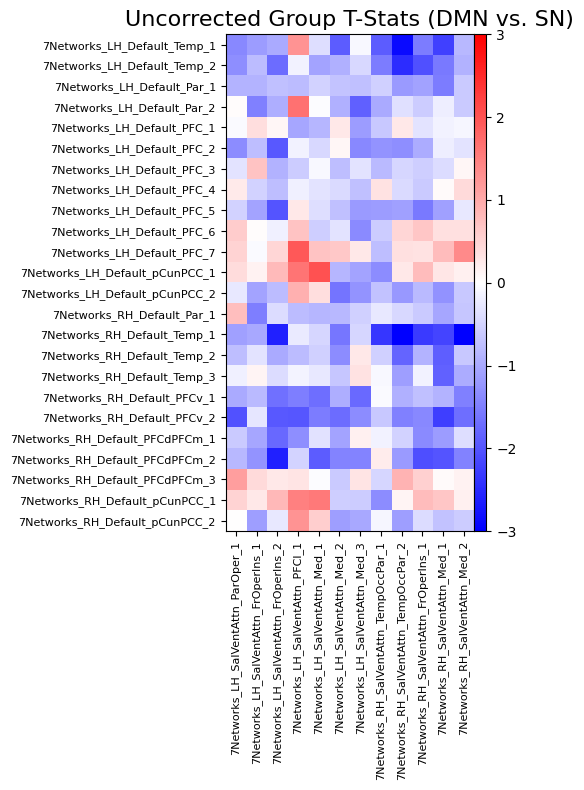

**C**	**DMN vs. FPN (participants who did not fall asleep)	D**	**DMN vs. FPN (all participants)**

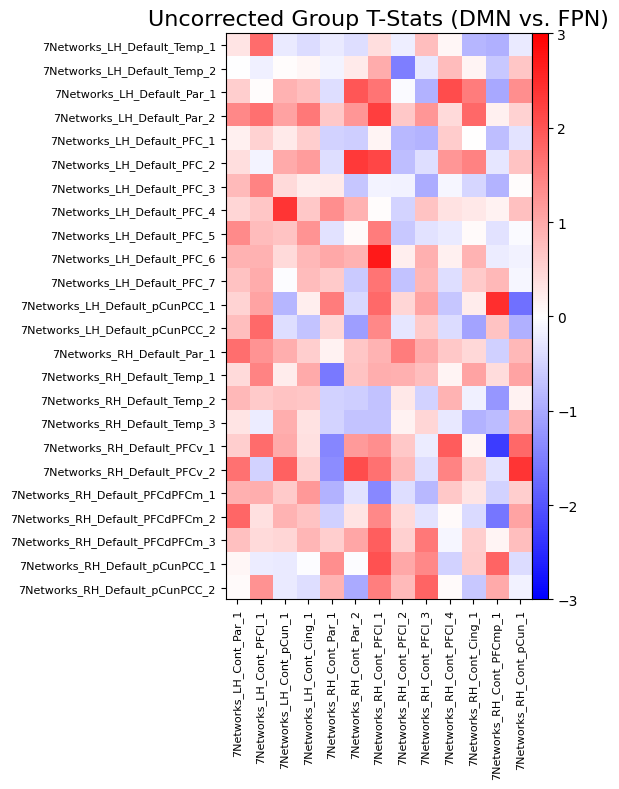

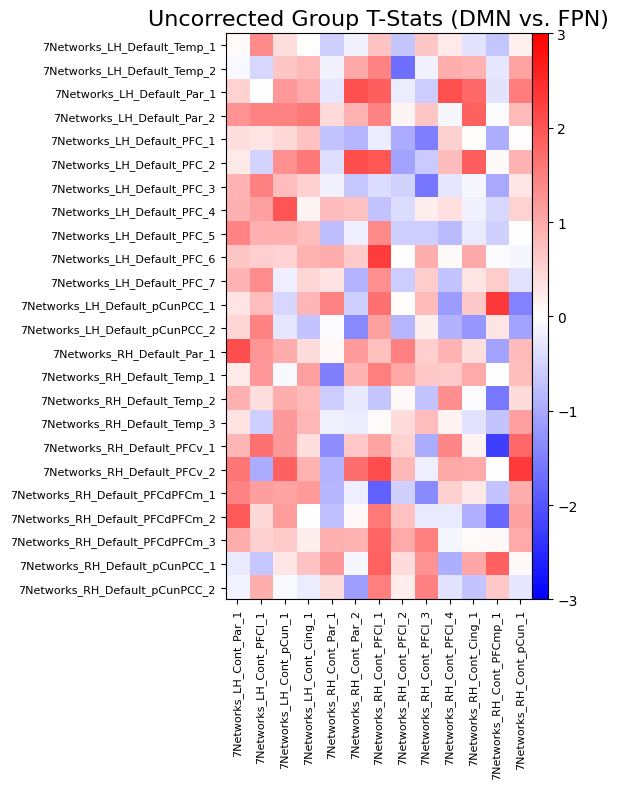

**E**	**SN vs. FPN (participants who did not fall asleep)	F**	**SN vs. FPN (all participants)**

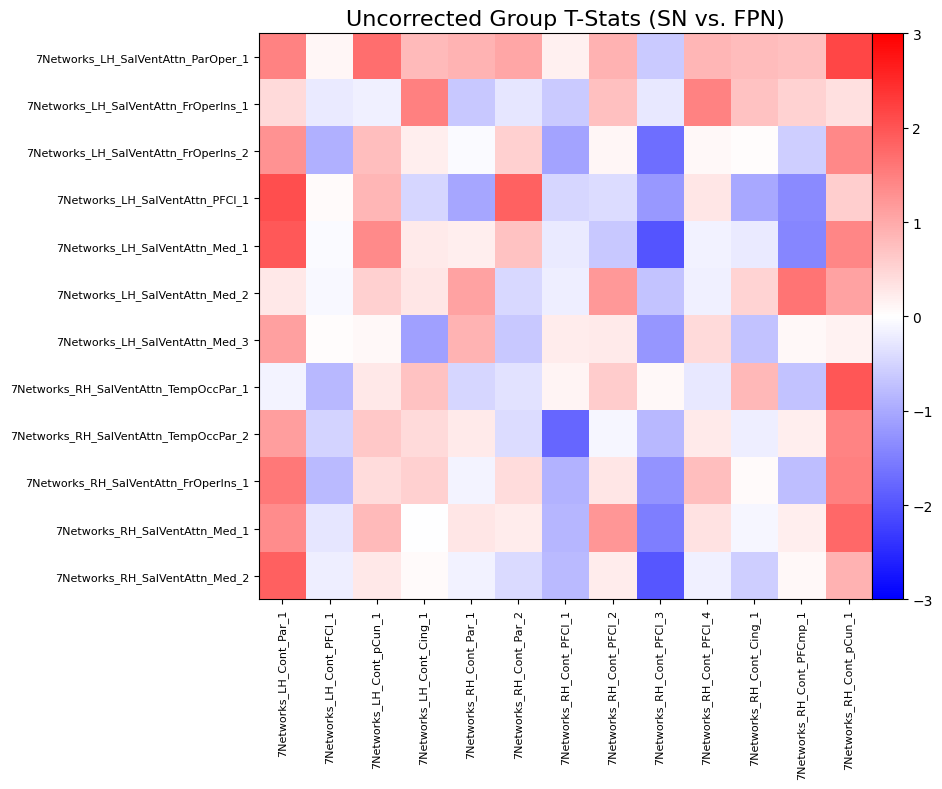

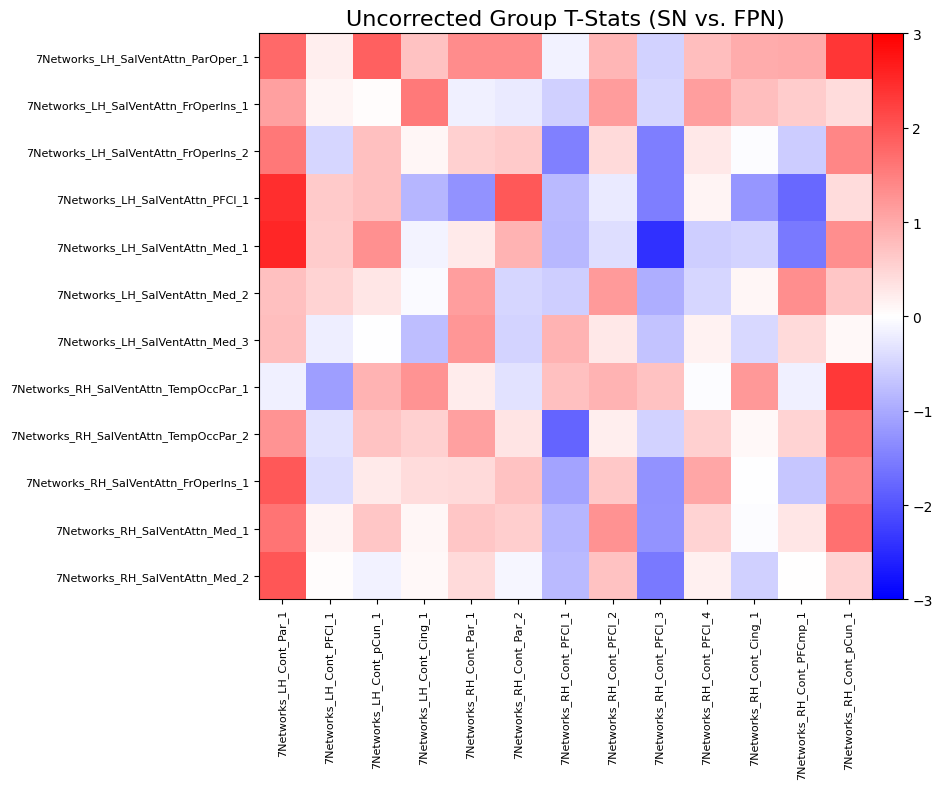

#### **Table S4.** Polysomnography-derived sleep measures between INS and GS groups at baseline

|  | *INS* | |  | *GS* | | *Group comparison (INS vs. GS)* | | |
| --- | --- | --- | --- | --- | --- | --- | --- | --- |
| Outcome | *n* | *M* (*SD*) |  | *n* | *M* (*SD*) | *t* (*df*) | *p* | $\beta$ [95% CI] |
| **Sleep** | | | | | | | | |
| TST (min.) | 56 | 346.66 (70.12) |  | 20 | 349.80 (30.92) | 0.17 (72) | .87 | 0.05 [-0.50, 0.59] |
| SOL (min.) | 56 | 15.32 (9.96) |  | 20 | 12.78 (7.41) | 1.09 (72) | .37 | 0.30 [-0.24, 0.84] |
| WASO (min.) | 56 | 47.62 (32.69) |  | 20 | 29.05 (16.55) | 2.58 (72) | .047* | 0.66 [0.15, 1.18] |
| SE (%) | 56 | 82.78 (10.75) |  | 20 | 88.20 (4.12) | -1.91 (72) | .12 | -0.50 [-1.03, 0.02] |

*Note*. **p* < .05; Standardized beta coefficients ($\beta$) are reported with 95% confidence intervals ([95% CI]). GS: Good sleeper group; INS: Insomnia group; SE: Sleep Efficiency; SOL: Sleep Onset Latency; TST: Total Sleep Time; WASO: Wake After Sleep Onset; SOL, TST, and WASO are reported in minutes (min.).

#### **Table S5.** Results for the effects of CBTi on cognitive performance

| Outcome | Group effect | | |  | Visit effect | | |  | Group*Visit interaction | | |
| --- | --- | --- | --- | --- | --- | --- | --- | --- | --- | --- | --- |
|  | *F (df)* | *p* | $\beta$ [95% CI] |  | *F (df)* | *p* | $\beta$ [95% CI] |  | *F (df)* | *p* | $\beta$ [95% CI] |
| **Sternberg working memory** | | | | | | | | | | | |
| Accuracy (% correct) | 0.97 (47.00) | .65 | 0.11 [-0.14, 0.36] |  | 0.43 (49.00) | .57 | 0.06 [-0.12, 0.23] |  | 3.79 (49.00) | .17 | 0.17 [-0.04, 0.38] |
| **Emotion reactivity** | | | | | | | | | | | |
| Valence rating for neutral images | 0.11 (47.00) | .80 | -0.04 [-0.28, 0.20] |  | 6.13 (49.00) | .02 | 0.16 [0.02, 0.30] |  | 0.23 (49.00) | .70 | 0.03 [-0.10, 0.16] |
| Valence rating for negative images | 0.07 (47.00) | .80 | 0.03 [-0.21, 0.27] |  | 6.64 (49.00) | .02 | 0.17 [0.02, 0.32] |  | 0.16 (49.00) | .70 | 0.03 [-0.11, 0.16] |
| Valence rating for sleep images | 0.55 (47.00) | .80 | 0.10 [-0.20, 0.40] |  | 3.96 (49.00) | .05 | 0.10 [0.00, 0.20] |  | 1.38 (49.00) | .70 | -0.06 [-0.18, 0.06] |
| **Face-name associative memory** | | | | | | | | | | | |
| **Immediate recall (50 stimuli)** | | | | | | | | | | | |
| Number of hits | 1.06 (28.00) | .58 | 0.15 [-0.19, 0.49] |  | 0.24 (30.00) | .67 | 0.05 [-0.15, 0.24] |  | 0.18 (30.00) | .94 | 0.04 [-0.16, 0.24] |
| Accuracy (Hit+CR - Miss+FA) | 0.85 (28.00) | .61 | -0.14 [-0.20, 0.48] |  | 3.56 (30.00) | .17 | 0.17 [-0.05, 0.38] |  | 0.06 (30.00) | .94 | -0.02 [-0.20, 0.16] |
| Recognition (Hit-FA) | 1.41 (28.00) | .58 | 0.18 [-0.18, 0.54] |  | 1.88 (30.00) | .34 | 0.12 [-0.08, 0.33] |  | 0.00 (30.00) | .99 | 0.00 [-0.18, 0.18] |
| **Delayed recall (50 stimuli)** | | | | | | | | | | | |
| Number of hits | 1.47 (28.00) | .50 | 0.19 [-0.18, 0.56] |  | 3.54 (30.00) | .17 | 0.11 [-0.03, 0.25] |  | 1.65 (30.00) | .45 | 0.07 [-0.06, 0.21] |
| Accuracy (Hit+CR - Miss+FA) | 2.39 (28.00) | .33 | 0.24 [-0.13, 0.62] |  | 5.40 (30.00) | .12 | 0.14 [-0.02, 0.29] |  | 1.02 (30.00) | .60 | 0.06 [-0.08, 0.20] |
| Recognition (Hit-FA) | 2.77 (28.00) | .32 | 0.26 [-0.13, 0.65] |  | 3.78 (30.00) | .17 | 0.12 [-0.03, 0.26] |  | 1.67 (30.00) | .45 | 0.08 [-0.06, 0.22] |
| **Delayed recall (100 stimuli)** | | | | | | | | | | | |
| Number of hits | 1.03 (28.00) | .53 | 0.16 [-1.19, 0.51] |  | 3.54 (30.00) | .13 | 0.13 [-0.03, 0.28] |  | 2.12 (30.00) | .39 | 0.10 [-0.06, 0.26] |
| Accuracy (Hit+CR - Miss+FA) | 1.70 (28.00) | .40 | 0.20 [-0.15, 0.56] |  | 6.86 (30.00) | .06 | 0.17 [0.00, 0.34] |  | 0.76 (30.00) | .73 | 0.06 [-0.09, 0.20] |
| Recognition (Hit-FA) | 2.11 (28.00) | .40 | 0.22 [-0.15, 0.59] |  | 5.37 (30.00) | .06 | 0.15 [0.00, 0.30] |  | 1.62 (30.00) | .46 | 0.08 [-0.07, 0.24] |
| **Delayed recall (50 stimuli) – Immediate recall (50 stimuli)** | | | | | | | | | | | |
| Change in number of hits | 1.07 (28.00) | .31 | 0.12 [-0.12, 0.36] |  | 0.86 (30.00) | .94 | 0.11 [-0.17, 0.38] |  | 0.14 (30.00) | .71 | 0.04 [-0.18, 0.27] |
| Change in accuracy (Hit+CR - Miss+FA) | 3.31 (28.00) | .15 | 0.23 [-0.06, 0.53] |  | 0.10 (30.00) | .94 | -0.04 [-0.29, 0.21] |  | 1.19 (30.00) | .54 | 0.13 [-0.14, 0.40] |
| Change in recognition (Hit-FA) | 2.80 (58.00) | .15 | 0.21 [-0.06, 0.47] |  | 0.01 (58.00) | .94 | -0.01 [-0.25, 0.23] |  | 0.85 (58.00) | .54 | 0.11 [-0.15, 0.38] |

*Note*. **p* < .05; The reported *p*-values were adjusted for multiple comparisons using the Benjamini-Hochberg False Discovery Rate procedure. Standardized beta coefficients ($\beta$) are reported with 95% confidence intervals ([95% CI]). CR: correct rejection; FA: false alarm.

#### **Table S6.** Results for effects of CBTi on performance for high- vs. low-reward face-name pairs

| Outcome | Group effect | | |  | Visit effect | | |  | Group*Visit interaction | | |
| --- | --- | --- | --- | --- | --- | --- | --- | --- | --- | --- | --- |
|  | *F (df)* | *p* | $\beta$ [95% CI] |  | *F (df)* | *p_._* | $\beta$ [95% CI] |  | *F (df)* | *p* | $\beta$ [95% CI] |
| **High reward** | | | | | | | | | | | |
| **Immediate recall (50 stimuli)** | | | | | | | | | | | |
| Number of hits | 0.28 (28.00) | .63 | 0.08 [-0.23, 0.39] |  | 0.10 (30.00) | .79 | -0.03 [-0.19, 0.14] |  | 1.29 (30.00) | .71 | 0.09 [-0.11, 0.29] |
| Accuracy (Hit+CR - Miss+FA) | 0.50 (28.00) | .58 | 0.11 [-0.21, 0.42] |  | 3.30 (30.00) | .16 | 0.15 [-0.04, 0.34] |  | 0.00 (30.00) | .95 | -0.01 [-0.17, 0.16] |
| Recognition (Hit-FA) | 0.78 (28.00) | .48 | 0.14 [-0.19, 0.47] |  | 1.74 (30.00) | .31 | 0.11 [-0.07, 0.29] |  | 0.07 (30.00) | .91 | 0.02 [-0.15, 0.19] |
| **Delayed recall (50 stimuli)** | | | | | | | | | | | |
| Number of hits | 0.30 (28.00) | .63 | 0.08 [-0.23, 0.40] |  | 6.23 (30.00) | .09 | 0.18 [-0.01, 0.37] |  | 3.15 (30.00) | .71 | 0.13 [-0.08, 0.33] |
| Accuracy (Hit+CR - Miss+FA) | 1.89 (28.00) | .30 | 0.21 [-0.13, 0.55] |  | 6.65 (30.00) | .09 | 0.19 [-0.01, 0.40] |  | 1.97 (30.00) | .71 | 0.11 [-0.09, 0.30] |
| Recognition (Hit-FA) | 2.05 (28.00) | .30 | 0.22 [-0.13, 0.57] |  | 5.49 (30.00) | .10 | 0.17 [-0.02, 0.37] |  | 2.57 (30.00) | .71 | 0.12 [-0.08, 0.32] |
| **Delayed recall (100 stimuli)** | | | | | | | | | | | |
| Number of hits | 0.43 (28.00) | .59 | 0.10 [-0.22, 0.42] |  | 4.36 (30.00) | .14 | 0.15 [-0.03, 0.33] |  | 2.54 (30.00) | .71 | 0.12 [-0.08, 0.31] |
| Accuracy (Hit+CR - Miss+FA) | 1.81 (28.00) | .30 | 0.20 [-0.13, 0.54] |  | 8.67 (30.00) | .09 | 0.20 [-0.01, 0.41] |  | 0.78 (30.00) | .78 | 0.06 [-0.10, 0.22] |
| Recognition (Hit-FA) | 1.95 (28.00) | .30 | 0.21 [-0.13, 0.55] |  | 7.01 (30.00) | .09 | 0.19 [-0.01, 0.38] |  | 1.52 (30.00) | .71 | 0.09 [-0.09, 0.26] |
| **Delayed recall (50 stimuli) – Immediate recall (50 stimuli)** | | | | | | | | | | | |
| Change in number of hits | 0.01 (58.00) | .92 | 0.01 [-0.22, 0.24] |  | 7.30 (58.00) | .09 | 0.31 [-0.02, 0.64] |  | 0.25 (58.00) | .88 | 0.06 [-0.19, 0.30] |
| Change in accuracy (Hit+CR - Miss+FA) | 1.16 (28.00) | .41 | 0.15 [-0.15, 0.46] |  | 0.36 (30.00) | .60 | 0.06 [-0.15, 0.28] |  | 2.46 (30.00) | .71 | 0.17 [-0.12, 0.45] |
| Change in recognition (Hit-FA) | 0.82 (28.00) | .48 | 0.12 [-0.16, 0.41] |  | 0.76 (30.00) | .47 | 0.10 [-0.14, 0.34] |  | 1.65 (30.00) | .71 | 0.15 [-0.14, 0.44] |
| **Low reward** | | | | | | | | | | | |
| **Immediate recall (50 stimuli)** | | | | | | | | | | | |
| Number of hits | 2.02 (28.00) | .30 | 0.20 [-0.12, 0.52] |  | 1.10 (30.00) | .38 | 0.12 [-0.12, 0.36] |  | 0.08 (30.00) | .91 | -0.03 [-0.27, 0.20] |
| Accuracy (Hit+CR - Miss+FA) | 2.26 (28.00) | .30 | 0.22 [-0.12, 0.55] |  | 4.27 (30.00) | .14 | 0.20 [0.04, 0.45] |  | 0.37 (30.00) | .88 | -0.06 [-0.28, 0.16] |
| Recognition (Hit-FA) | 2.28 (28.00) | .30 | 0.21 [-0.12, 0.54] |  | 4.08 (30.00) | .14 | 0.21 [-0.04, 0.46] |  | 0.45 (30.00) | .88 | -0.07 [-0.30, 0.16] |
| **Delayed recall (50 stimuli)** | | | | | | | | | | | |
| Number of hits | 3.06 (28.00) | .30 | 0.27 [-0.12, 0.65] |  | 0.06 (30.00) | .81 | 0.02 [-0.13, 0.16] |  | 0.01 (30.00) | .95 | 0.01 [-0.14, 0.15] |
| Accuracy (Hit+CR - Miss+FA) | 4.37 (28.00) | .30 | 0.33 [-0.11, 0.76] |  | 1.29 (30.00) | .38 | 0.08 [-0.07, 0.22] |  | 0.03 (30.00) | .94 | 0.01 [-0.12, 0.15] |
| Recognition (Hit-FA) | 5.00 (28.00) | .30 | 0.35 [-0.10, 0.80] |  | 0.54 (30.00) | .53 | -0.05 [-0.09, 0.19] |  | 0.16 (30.00) | .88 | 0.03 [-0.12, 0.17] |
| **Delayed recall (100 stimuli)** | | | | | | | | | | | |
| Number of hits | 1.88 (28.00) | .30 | 0.21 [-0.13, 0.55] |  | 1.20 (30.00) | .38 | 0.08 [-0.08, 0.25] |  | 0.76 (30.00) | .78 | 0.07 [-0.11, 0.24] |
| Accuracy (Hit+CR - Miss+FA) | 3.19 (28.00) | .30 | 0.27 [-0.11, 0.66] |  | 3.36 (30.00) | .16 | 0.13 [-0.04, 0.30] |  | 0.15 (30.00) | .88 | 0.03 [-0.13, 0.18] |
| Recognition (Hit-FA) | 4.00 (28.00) | .30 | 0.30 [-0.11, 0.71] |  | 2.81 (30.00) | .19 | 0.13 [-0.05, 0.30] |  | 0.30 (30.00) | .88 | 0.04 [-0.13, 0.21] |
| **Delayed recall (50 stimuli) – Immediate recall (50 stimuli)** | | | | | | | | | | | |
| Change in number of hits | 1.57 (28.00) | .33 | 0.15 [-0.11, 0.42] |  | 1.14 (30.00) | .38 | -0.13 [-0.38, 0.13] |  | 0.19 (30.00) | .88 | 0.05 [-0.20, 0.30] |
| Change in accuracy (Hit+CR - Miss+FA) | 2.67 (58.00) | .30 | 0.20 [-0.10, 0.51] |  | 2.55 (58.00) | .20 | -0.20 [-0.47, 0.08] |  | 0.86 (58.00) | .78 | 0.11 [-0.17, 0.40] |
| Change in recognition (Hit-FA) | 3.11 (28.00) | .30 | 0.22 [-0.10, 0.54] |  | 3.48 (30.00) | .16 | -0.22 [-0.51, 0.06] |  | 1.39 (30.00) | .71 | 0.14 [-0.15, 0.44] |

*Note*. **p* < .05; The reported *p*-values were adjusted for multiple comparisons using the Benjamini-Hochberg False Discovery Rate procedure. Standardized beta coefficients ($\beta$) are reported with 95% confidence intervals ([95% CI]). CR: Correct Rejection; FA: False Alarm.

#### **Figure S4.** Effects of CBTi on within-network resting-state functional connectivity

(A) Resting-state functional connectivity (rs-FC) changes within the default mode network (DMN) following cognitive-behavioural therapy for insomnia (CBTi) or wait-list period

(B) rs-FC changes within the DMN

(C) rs-FC changes within the salience network (SN)

(D) rs-FC changes within the SN

(E) rs-FC changes within the frontoparietal network (FPN)

(F) rs-FC changes within the FPN

Panels A, C, and E include participants with insomnia disorder who did not sleep during the resting-state task, whereas panels B, D, and F include participants with insomnia disorder who did not sleep during the resting-state task. Age and sex were included as covariates. As no connections survived following false discovery rate (FDR) correction set at .05, only uncorrected results are displayed.

**A**	**DMN (participants who did not fall asleep)**

**No interaction**

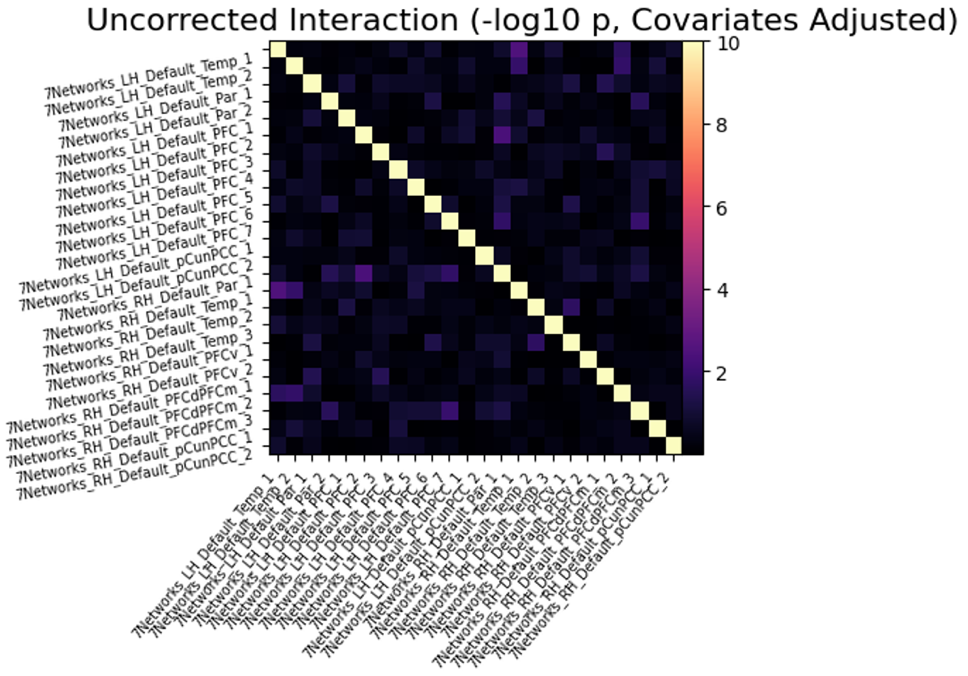

**B**	**DMN (all participants)**

**No interaction**

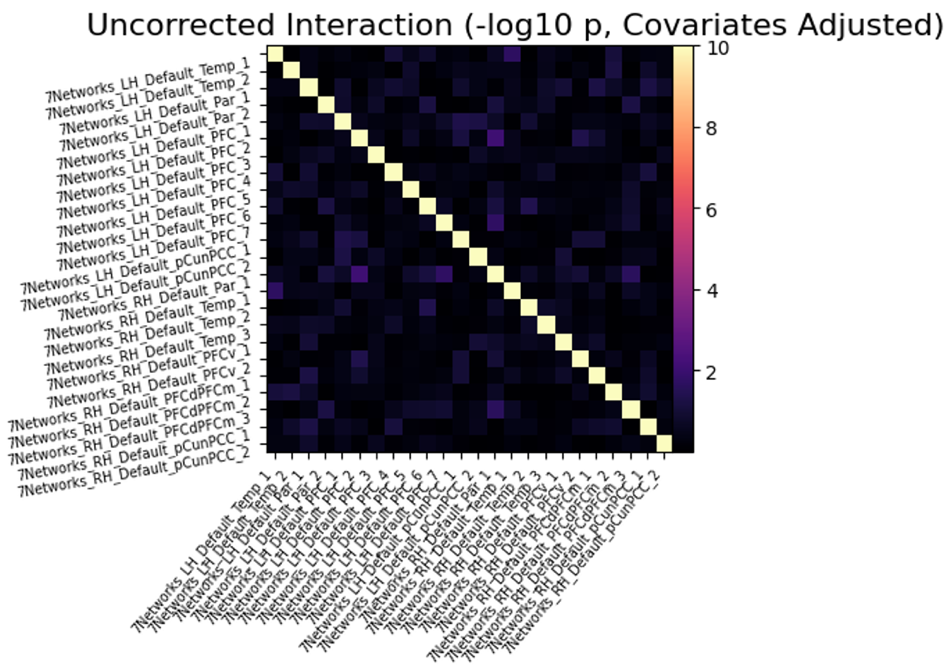

**C**	**SN (participants who did not fall asleep)**

**No interaction**

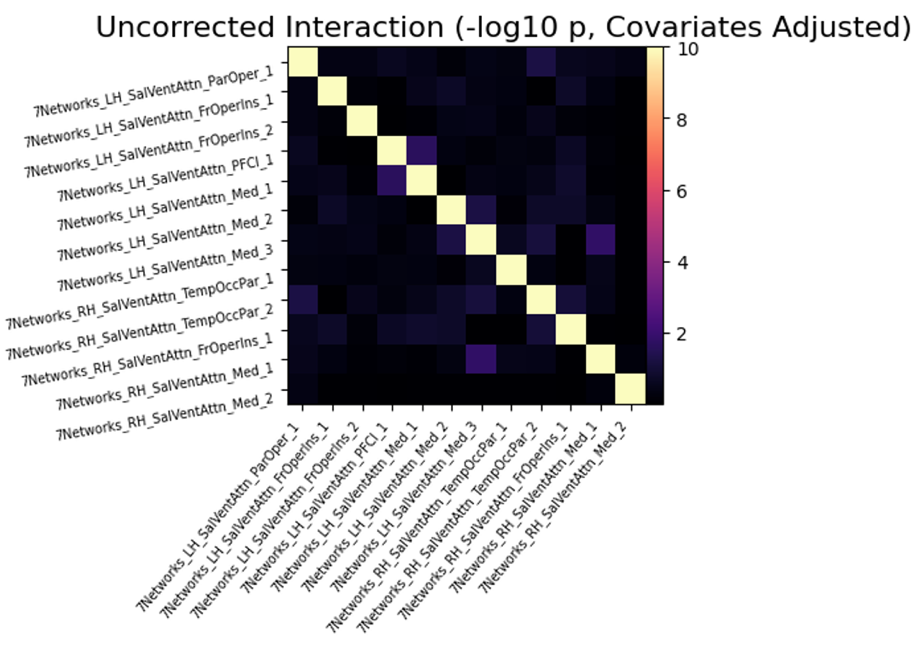

**D**	**SN (all participants)**

**No interaction**

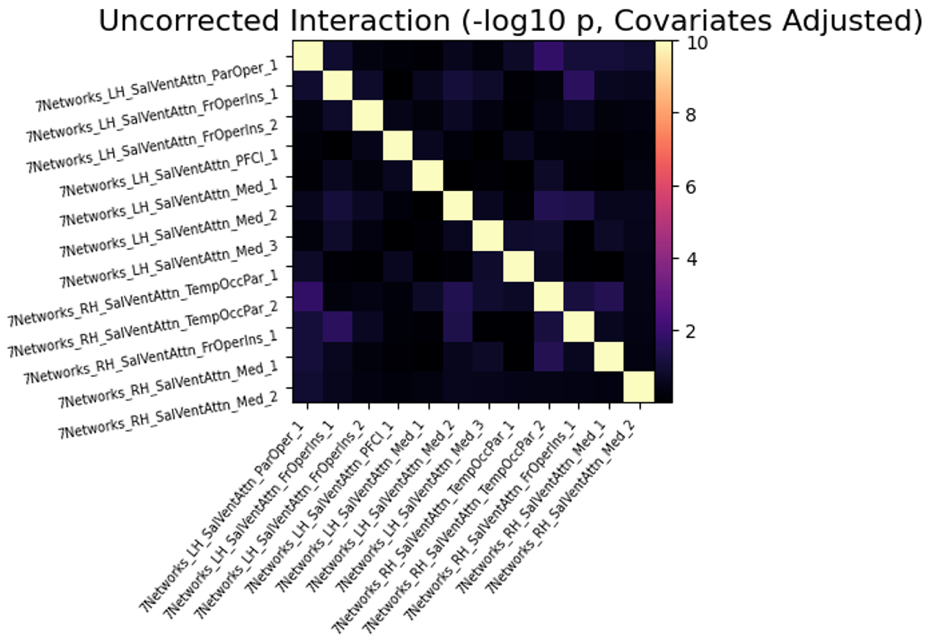

**E**	**FPN (participants who did not fall asleep)**

**No interaction**

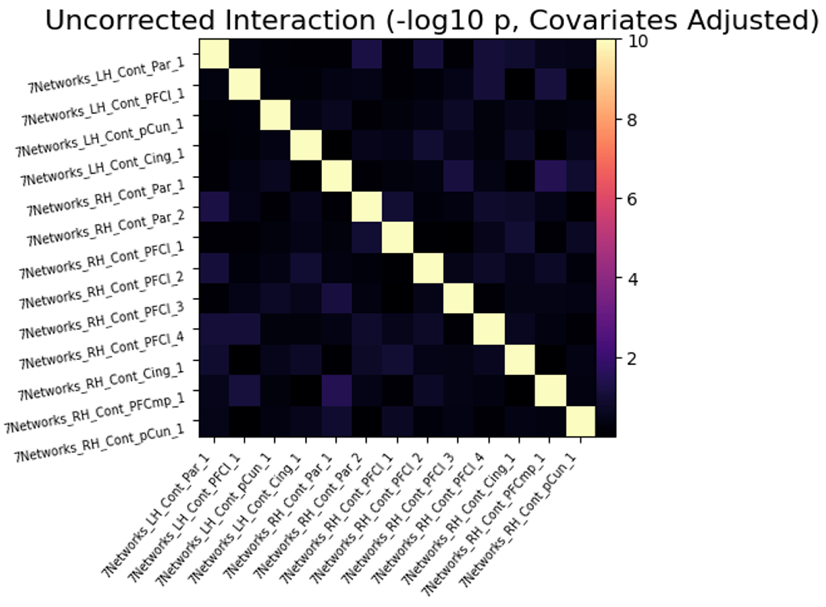

**F**	**FPN (all participants)**

**No interaction**

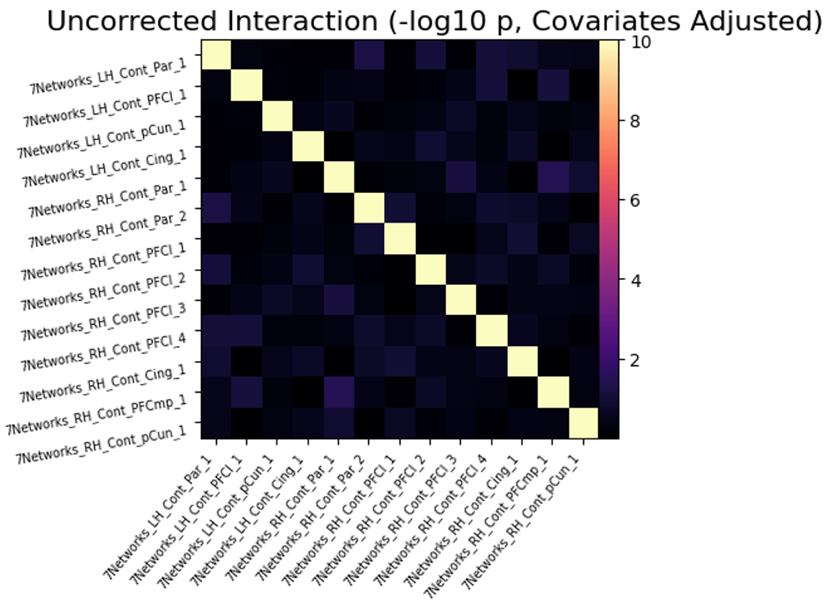

#### **Figure S5.** Effects of CBTi on between-network resting-state functional connectivity

(A) No changes in default mode network (DMN)-salience network (SN) connectivity following cognitive-behavioural therapy for insomnia or wait-list period

(B) No changes in DMN-SN connectivity

(C) No changes in DMN and frontoparietal network (FPN) connectivity

(D) No changes in DMN-FPN connectivity

(E) No SN-FPN connectivity changes

(F) No SN-FPN connectivity changes

(G) Main effect of Group on SN-FPN connectivity

(H) Main effect of Group on SN-FPN connectivity

(I) No main effect of Visit on SN-FPN connectivity

(J) No main effect of Visit on SN-FPN connectivity

Panels A, C, E, G, and I include participants with insomnia disorder who did not sleep during the resting-state task, whereas panels B, D, F, H, and J include all participants who completed the task at V1 and V2. Panels G and H show a statistically significant main effect of group after false discovery rate (FDR) correction set at .05, with the WL group exhibiting lower connectivity between the right dorsal anterior cingulate cortex and right dorsolateral prefrontal cortex, on average, than the TX group (*z* = -3.63, *p_FDR_* = .04).

**A**	**DMN vs. SN (participants who did not fall asleep)	B**	**DMN vs. SN (all participants)**

**No interaction	No interaction**
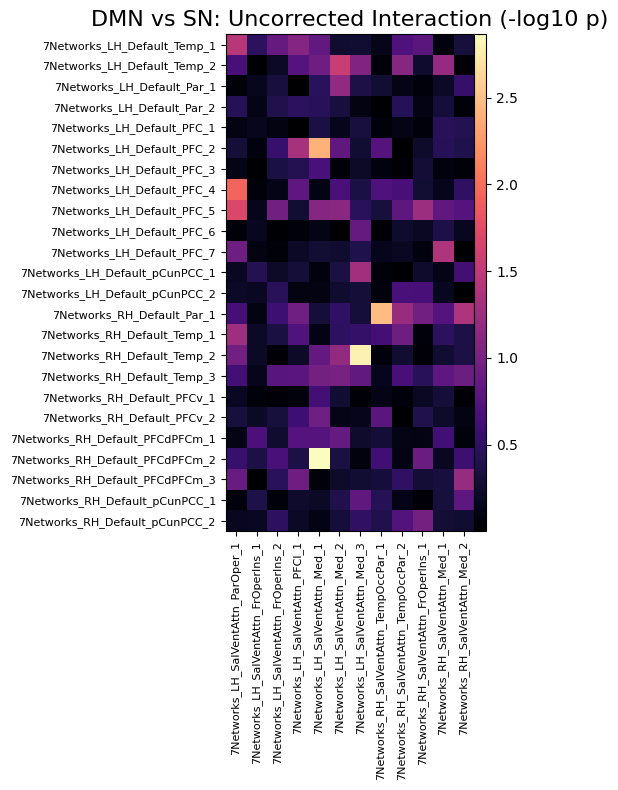

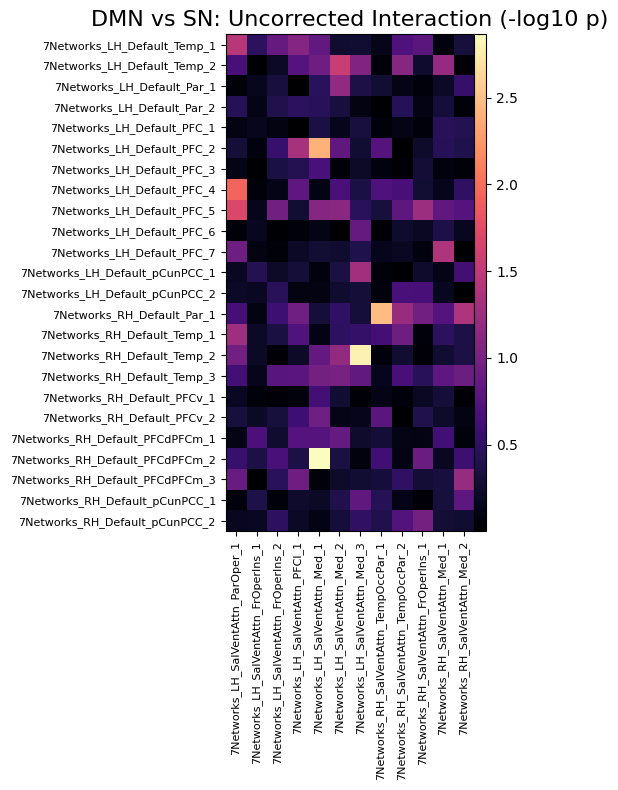

**C**	**DMN vs. FPN (participants who did not fall asleep)	D**	**DMN vs. FPN (all participants)**

**No interaction	No interaction**

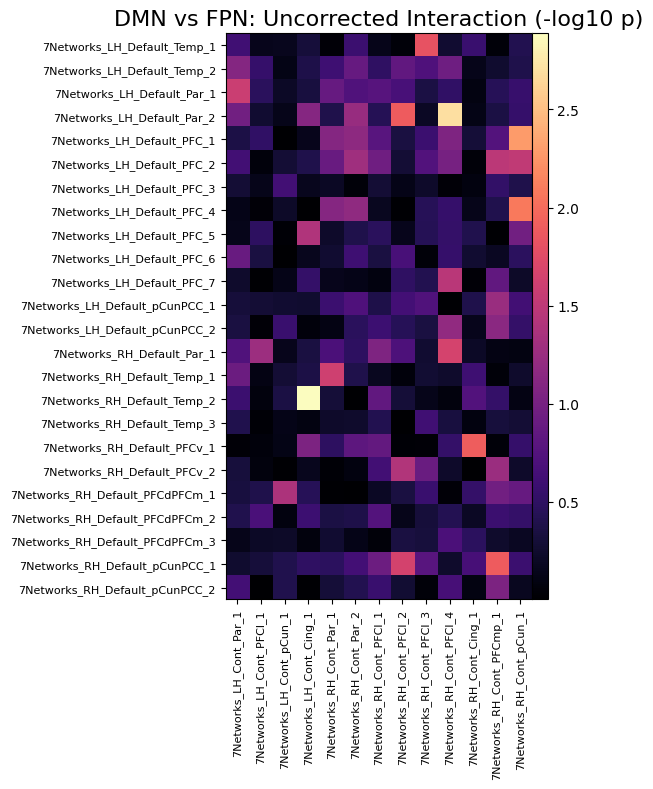

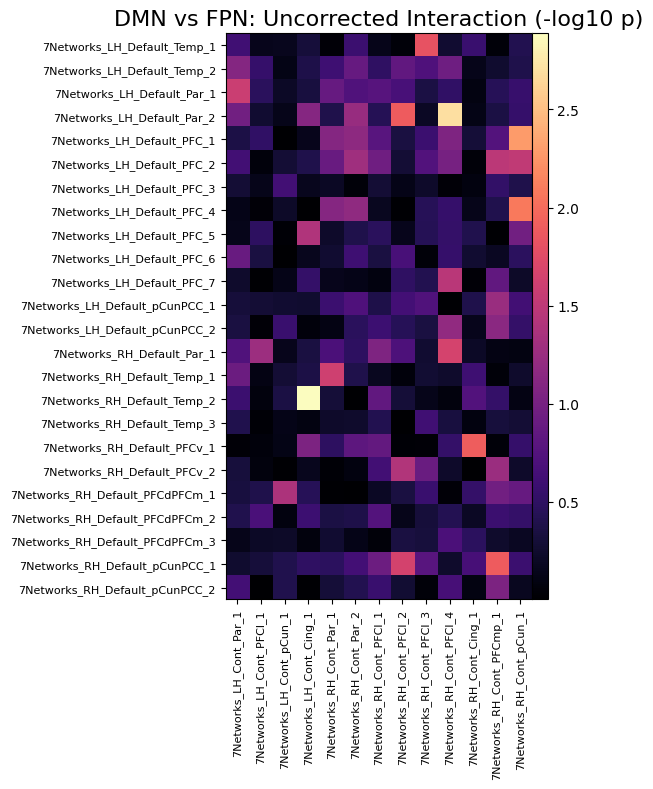

**E**	**SN vs. FPN (participants who did not fall asleep)**

**No interaction**

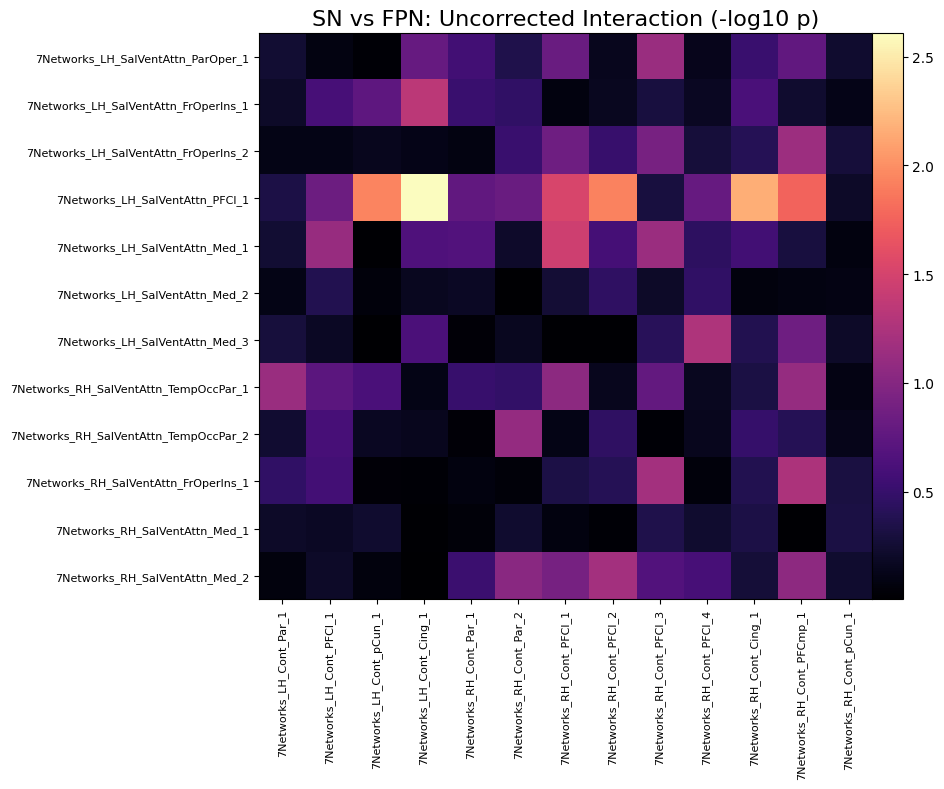

**F**	**SN vs. FPN (all participants)**

**No interaction**

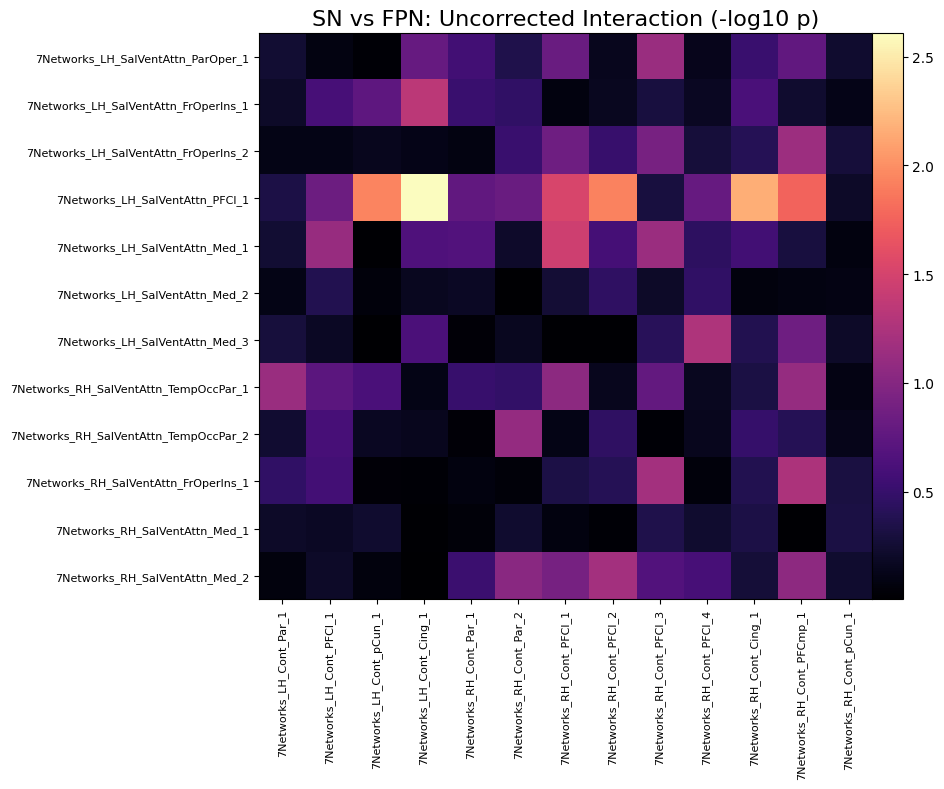

**G**	**SN vs. FPN (participants who did not fall asleep)**

**Main effect of Group**

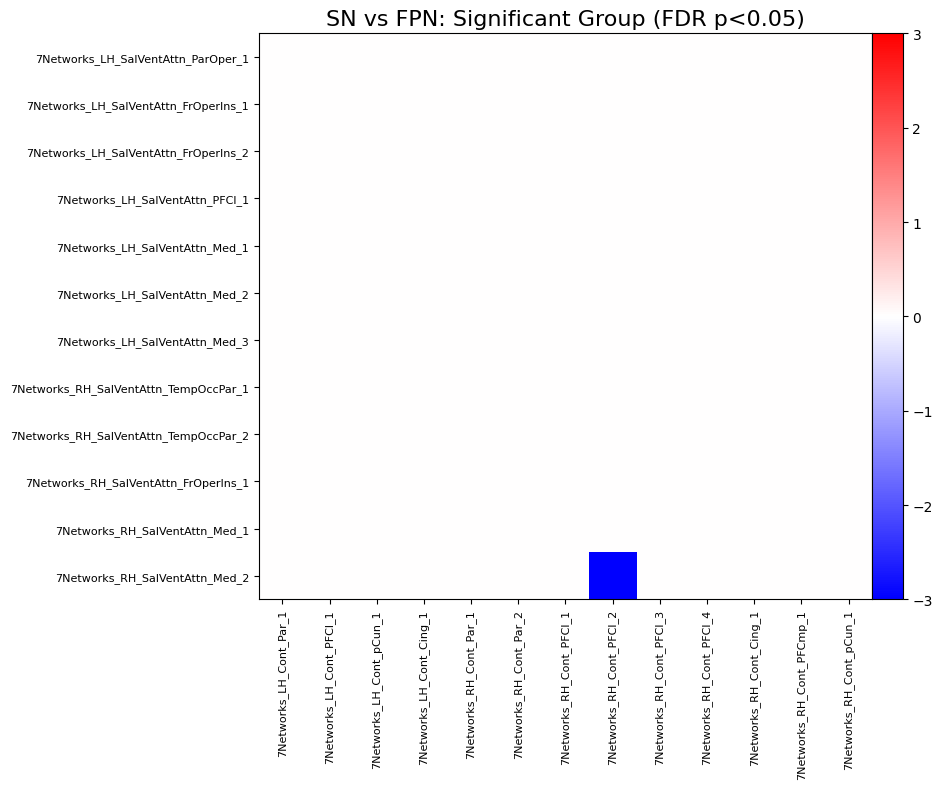

**H**	**SN vs. FPN (all participants)**

**Main effect of Group**

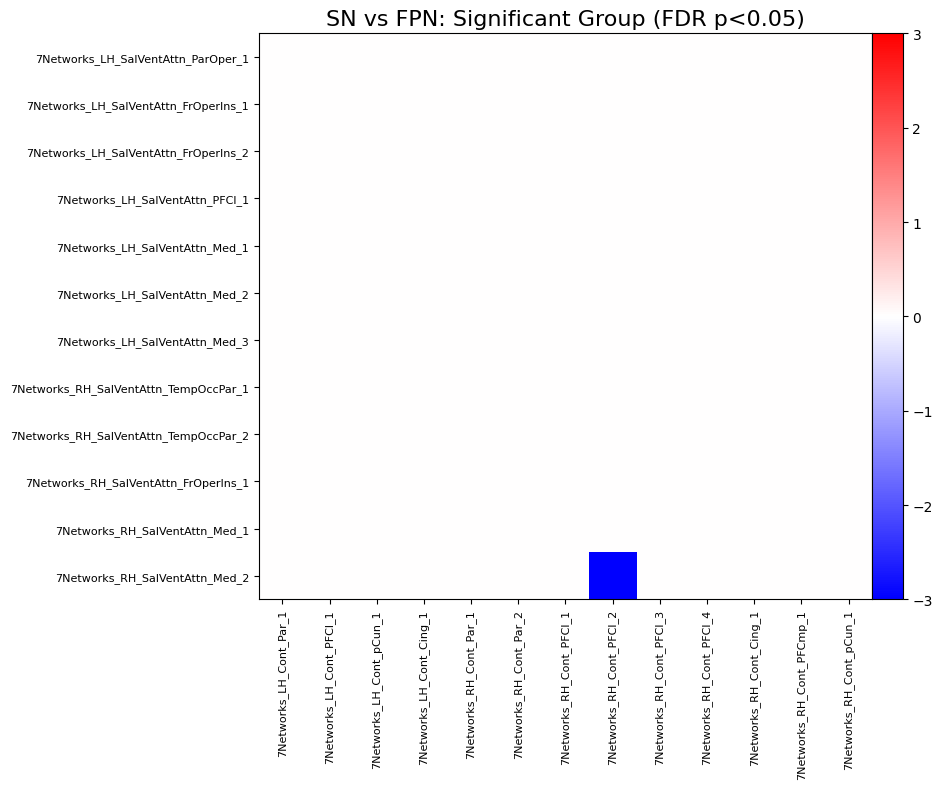

**I**	**SN vs. FPN (participants who did not fall asleep)**

**No main effect of Visit**

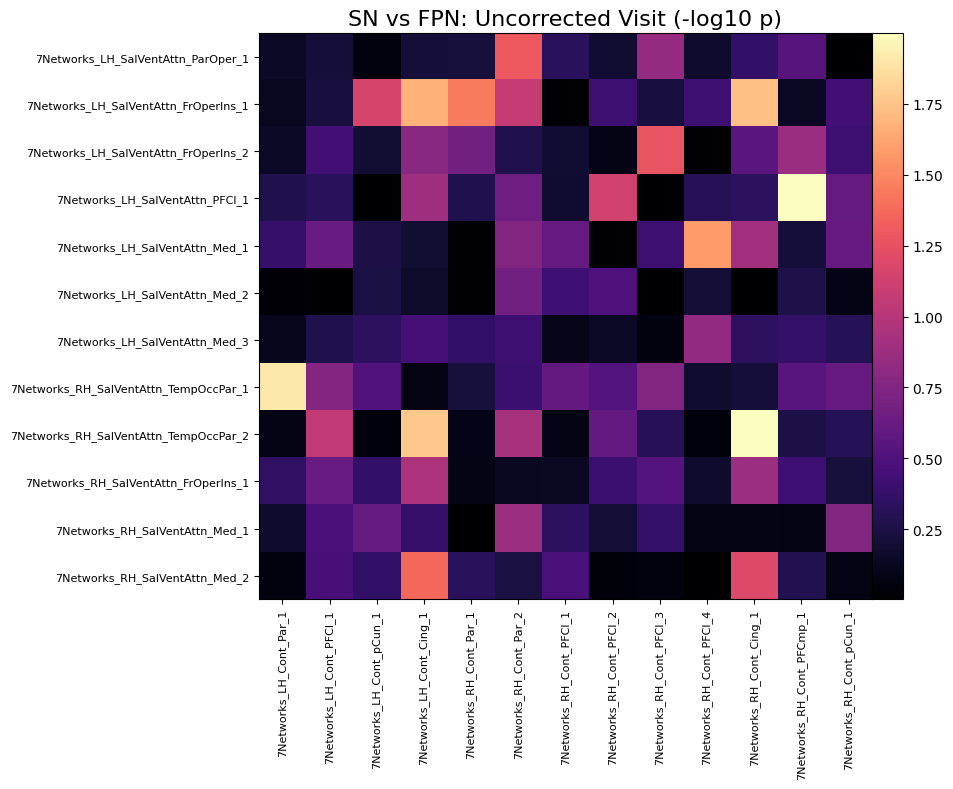

**J**	**SN vs. FPN (all participants)**

**No main effect of Visit**

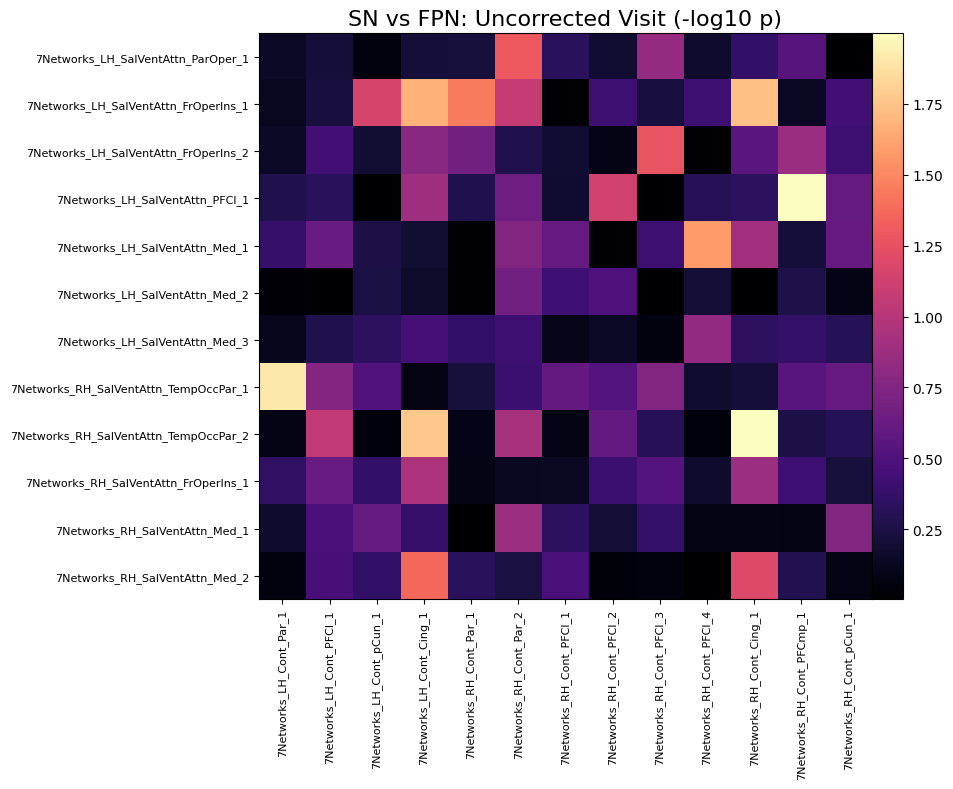

#### **Table S7.** Descriptive statistics for self-reported sleep, emotion, and cognition measures of TX and WL groups at V1, V2, and one-year follow-up

| **Outcome/Visit** | **TX** | |  | **WL** | |
| --- | --- | --- | --- | --- | --- |
|  | *n* | *M* (*SD*) |  | *n* | *M* (*SD*) |
| **Sleep** | | | | | |
| ISI |  |  |  |  |  |
| V1 | 29 | 16.97 (3.94) |  | 27 | 15.67 (3.54) |
| V2 | 28 | 7.46 (5.66) |  | 26 | 13.54 (6.24) |
| 1YFU | 26 | 6.81 (4.60) |  | 22 | 7.82 (4.55) |
| PSQI |  |  |  |  |  |
| V1 | 28 | 9.29 (2.72) |  | 27 | 10.00 (2.72) |
| V2 | 28 | 4.96 (3.02) |  | 26 | 7.77(3.55) |
| 1YFU | 26 | 4.88 (2.25) |  | 22 | 5.86 (2.96) |
| DBAS-16 |  |  |  |  |  |
| V1 | 29 | 5.07 (1.59) |  | 27 | 5.07 (1.52) |
| V2 | 28 | 3.46 (1.84) |  | 26 | 5.38 (1.97) |
| 1YFU | 26 | 3.56 (1.45) |  | 22 | 3.91 (1.60) |
| DISRS |  |  |  |  |  |
| V1 | 29 | 40.17 (10.15) |  | 27 | 42.89 (11.79) |
| V2 | 28 | 30.57 (11.79) |  | 26 | 38.81 (10.74) |
| 1YFU | 26 | 31.58 (9.90) |  | 22 | 32.09 (10.14) |
| TST (min.) |  |  |  |  |  |
| V1 | 26 | 388.78 (65.05) |  | 23 | 394.18 (40.82) |
| V2 | 25 | 407.81 (52.79) |  | 21 | 394.10(46.01) |
| 1YFU | 24 | 429.88 (55.81) |  | 22 | 412.55 (40.89) |
| SOL (min.) |  |  |  |  |  |
| V1 | 26 | 29.93(15.04) |  | 23 | 24.53(18.30) |
| V2 | 25 | 17.73 (11.96) |  | 21 | 25.54 (14.31) |
| 1YFU | 24 | 15.26 (11.18) |  | 22 | 21.60 (17.50) |
| WASO (min.) |  |  |  |  |  |
| V1 | 26 | 47.59 (33.06) |  | 23 | 33.71 (27.30) |
| V2 | 25 | 32.77(23.84) |  | 21 | 39.30 (26.54) |
| 1YFU | 24 | 29.68 (19.97) |  | 22 | 35.39 (26.00) |
| Number of awakenings |  |  |  |  |  |
| V1 | 26 | 2.53 (1.77) |  | 23 | 1.93 (1.82) |
| V2 | 25 | 1.74(1.07) |  | 21 | 1.91(1.77) |
| 1YFU | 24 | 1.72 (1.02) |  | 22 | 1.55 (1.24) |
| SE (%) |  |  |  |  |  |
| V1 | 26 | 76.46 (10.73) |  | 23 | 80.82 (10.88) |
| V2 | 25 | 84.74 (9.98) |  | 21 | 80.01 (10.15) |
| 1YFU | 24 | 88.58 (10.08) |  | 22 | 83.25 (7.73) |
| **Emotions** | | | | | |
| BDI-II |  |  |  |  |  |
| V1 | 29 | 10.62 (7.66) |  | 27 | 11.22 (7.28) |
| V2 | 28 | 6.07 (10.40) |  | 26 | 10.81 (8.02) |
| 1YFU | 26 | 5.65 (5.78) |  | 22 | 7.68 (7.17) |
| PANAS – positive affect |  |  |  |  |  |
| V1 | 28 | 34.00 (6.90) |  | 27 | 28.78 (7.21) |
| V2 | 28 | 35.43(7.80) |  | 26 | 28.77 (8.17) |
| 1YFU | 26 | 33.81 (8.19) |  | 22 | 30.68 (8.12) |
| PANAS – negative affect |  |  |  |  |  |
| V1 | 28 | 17.57 (5.72) |  | 27 | 19.30 (6.05) |
| V2 | 28 | 17.00 (7.36) |  | 26 | 19.15 (7.74) |
| 1YFU | 26 | 17.35 (6.63) |  | 22 | 17.91 (6.62) |
| STICSA – total |  |  |  |  |  |
| V1 | 29 | 34.48 (10.16) |  | 27 | 33.78 (9.41) |
| V2 | 28 | 30.07 (10.58) |  | 26 | 33.08 (8.57) |
| 1YFU | 26 | 29.46 (8.58) |  | 22 | 30.36 (7.39) |
| STICSA – cognitive |  |  |  |  |  |
| V1 | 29 | 18.72 (6.67) |  | 27 | 18.22 (5.70) |
| V2 | 28 | 15.29 (5.73) |  | 26 | 18.00 (5.28) |
| 1YFU | 26 | 15.27 (5.62) |  | 22 | 16.09 (4.92) |
| STICSA – somatic |  |  |  |  |  |
| V1 | 29 | 15.76 (4.70) |  | 27 | 15.56 (4.44) |
| V2 | 28 | 14.79 (5.48) |  | 26 | 15.08 (4.68) |
| 1YFU | 26 | 14.19 (3.48) |  | 22 | 14.27 (2.78) |
| **Cognition** | | | | | |
| SASCI-Q |  |  |  |  |  |
| V1 | 28 | 86.11 (26.54) |  | 27 | 80.44 (27.69) |
| V2 | 28 | 74.21 (29.04) |  | 26 | 82.27 (32.56) |
| 1YFU | 26 | 72.85 (29.13) |  | 22 | 72.32 (32.20) |

*Note*. Continuous variables are presented as means (*M*) and *standard deviations* (*SD*); 1YFU: one-year follow-up; BDI-II: Beck Depression Inventory-II; DBAS-16: Dysfunctional Beliefs and Attitudes about Sleep; DISRS: Daytime Insomnia Symptom Response Scale; ISI: Insomnia Severity Index; PANAS: Positive Affect Negative Affect Scale; PSQI: Pittsburgh Sleep Quality Index; SASCI-Q: Sahlgrenska Academy Self-reported Cognitive Impairment questionnaire; SE: Sleep Efficiency; SOL: Sleep Onset Latency; STICSA: State-Trait Inventory for Cognitive and Somatic Anxiety; TST: Total Sleep Time; TX: Immediate CBTi group; WASO: Wake After Sleep Onset; WL: Wait-list control group; SOL, TST, and WASO are reported in minutes (min.).

#### **Table S8.** Results for the effects of CBTi on self-reported sleep, emotion, and cognition measures of TX and WL groups at V1, V2, and one-year follow-up

| Outcome | Group effect | | |  | Visit effect | | |  | Group*Visit interaction | | |
| --- | --- | --- | --- | --- | --- | --- | --- | --- | --- | --- | --- |
|  | *F (df)* | *p* | $\beta$ [95% CI] |  | *F (df)* | *p* | $\eta_{p}^{2}$ [95% CI] |  | *F (df)* | *p* | $\eta_{p}^{2}$ [95% CI] |
| **Cognition** |  |  |  |  |  |  |  |  |  |  |  |
| SASCI-Q | 0.05 (1, 51.10) | .91 | -0.03 [-0.27, 0.22] |  | 5.85 (2, 97.05) | .01* | 0.11 [0.02, 1.00] |  | 4.29 (2, 97.07) | .04* | 0.08 [0.01, 1.00] |
| **Emotions** |  |  |  |  |  |  |  |  |  |  |  |
| BDI-II | 1.41 (1, 51.72) | .44 | 0.12 [-0.08, 0.32] |  | 7.18 (2, 99.94) | .002* | 0.13 [0.03, 1.00] |  | 1.93 (2, 99.98) | .24 | 0.04 [0.00, 1.00] |
| PANAS – positive affect | 7.32 (1, 52.50) | .10 | -0.28 [-0.49, -0.08] |  | 0.85 (2, 99.51) | .43 | 0.02 [0.00, 1.00] |  | 1.63 (2, 99.54) | .28 | 0.03 [0.00, 1.00] |
| PANAS – negative affect | 0.13 (1, 50.61) | .91 | 0.04 [-0.20, 0.28] |  | 1.22 (2, 96.79) | .33 | 0.03 [0.00, 1.00] |  | 1.22 (2, 96.81) | .33 | 0.03 [0.00, 1.00] |
| STICSA – total | 0.00 (1, 49.69) | 1.00 | 0.00 [-0.23, 0.23] |  | 6.80 (2, 96.92) | .003* | 0.12 [0.03, 1.00] |  | 1.36 (2, 96.95) | .32 | 0.03 [0.00, 1.00] |
| STICSA – cognitive | 0.08 (1, 50.07) | .91 | 0.03 [-0.19, 0.26] |  | 7.44 (2, 97.48) | .002* | 0.13 [0.04, 1.00] |  | 2.83 (2, 97.52) | .12 | 0.06 [0.00, 1.00] |
| STICSA – somatic | 0.10 (1, 49.41) | .91 | -0.04 [-0.26, 0.19] |  | 2.40 (2, 97.24) | .12 | 0.05 [0.00, 1.00] |  | 0.03 (2, 97.28) | .97 | 0.00 [0.00, 1.00] |
| **Sleep** |  |  |  |  |  |  |  |  |  |  |  |
| ISI | 1.85 (1, 51.14) | .40 | 0.11 [-0.05, 0.28] |  | 95.48 (2, 99.41) | < .001* | 0.66 [0.57, 1.00] |  | 17.65 (2, 99.45) | < .001* | 0.26 [0.14, 1.00] |
| PSQI | 3.98 (1, 50.13) | .24 | 0.18 [0.00, 0.36] |  | 60.00 (2, 97. 94) | < .001* | 0.55 [0.44, 1.00] |  | 5.10 (2, 97.97) | .03* | 0.09 [0.02, 1.00] |
| DBAS-16 | 3.56 (1, 52.49) | .24 | 0.20 [-0.01, 0.41] |  | 18.93 (2, 100.06) | < .001* | 0.28 [0.15, 1.00] |  | 12.55 (2, 100.09) | < .001* | 0.20 [0.09, 1.00] |
| DISRS | 2.10 (1, 51.39) | .40 | 0.14 [-0.05, 0.32] |  | 24.52 (2, 99.55) | < .001* | 0.33 [0.21, 1.00] |  | 3.68 (2, 99.59) | .06 | 0.07 [0.00, 1.00] |
| TST | 0.82 (1, 48.91) | .56 | -0.11 [-0.34, 0.13] |  | 11.93 (2, 84.65) | < .001* | 0.22 [0.10, 1.00] |  | 0.99 (2, 84.77) | .38 | 0.02 [0.00, 1.00] |
| SOL | 0.88 (1, 47.74) | .56 | 0.11 [-0.12, 0.34] |  | 8.46 (2, 84.99) | < .001* | 0.17 [0.05, 1.00] |  | 3.63 (2, 85.11) | .08 | 0.08 [0.00, 1.00] |
| WASO | 0.00 (1, 48.53) | .99 | 0.00 [-0.23, 0.23] |  | 2.07 (2, 86.77) | .13 | 0.05 [0.00, 1.00] |  | 3.12 (2, 86, 89) | .08 | 0.07 [0.00, 1.00] |
| Number of awakenings | 0.87 (1, 47.36) | .56 | -0.13 [-0.39, 0.14] |  | 5.01 (2, 81.85) | .01* | 0.11 [0.02, 1.00] |  | 2.45 (2, 81.94) | .12 | 0.06 [0.00, 1.00] |
| SE | 0.58 (1, 48.15) | .56 | -0.08 [-0.30, 0.14] |  | 9.67 (2, 86.31) | < .001* | 0.18 [0.07, 1.00] |  | 4.85 (2, 86.43) | .05 | 0.10 [0.02, 1.00] |

*Note*. **p* < .05; The reported *p*-values were adjusted for multiple comparisons using the Benjamini-Hochberg False Discovery Rate procedure. Standardized beta coefficients ($\beta$) and partial eta-squared ($\eta_{p}^{2}$) are reported with 95% confidence intervals ([95% CI]). BDI-II: Beck Depression Inventory-II; DBAS-16: Dysfunctional Beliefs and Attitudes about Sleep; DISRS: Daytime Insomnia Symptom Response Scale; ISI: Insomnia Severity Index; PANAS: Positive Affect Negative Affect Scale; PSQI: Pittsburgh Sleep Quality Index; SASCI-Q: Sahlgrenska Academy Self-reported Cognitive Impairment questionnaire; SE: Sleep Efficiency; SOL: Sleep Onset Latency; STICSA: State-Trait Inventory for Cognitive and Somatic Anxiety; TST: Total Sleep Time; WASO: Wake After Sleep Onset.

#### **Figure S6.** Effects of CBTi on self-reported cognition, emotion, and sleep measures of TX and WL groups at V1, V2, and one-year follow-up

(A) Cognition measures

(B) Emotion measures

(C) Sleep Measures

The TX group is shown in grey with a dashed line, whereas the WL group is shown in blue with a solid line. Error bars represent standard error of the mean (SEM). 1YFU: one-year follow-up; BDI-II: Beck Depression Inventory-II; DBAS-16: Dysfunctional Beliefs and Attitudes about Sleep; DISRS: Daytime Insomnia Symptom Response Scale; ISI: Insomnia Severity Index; PANAS: Positive Affect Negative Affect Scale; PSQI: Pittsburgh Sleep Quality Index; SASCI-Q: Sahlgrenska Academy Self-reported Cognitive Impairment questionnaire; STICSA: State-Trait Inventory for Cognitive and Somatic Anxiety.

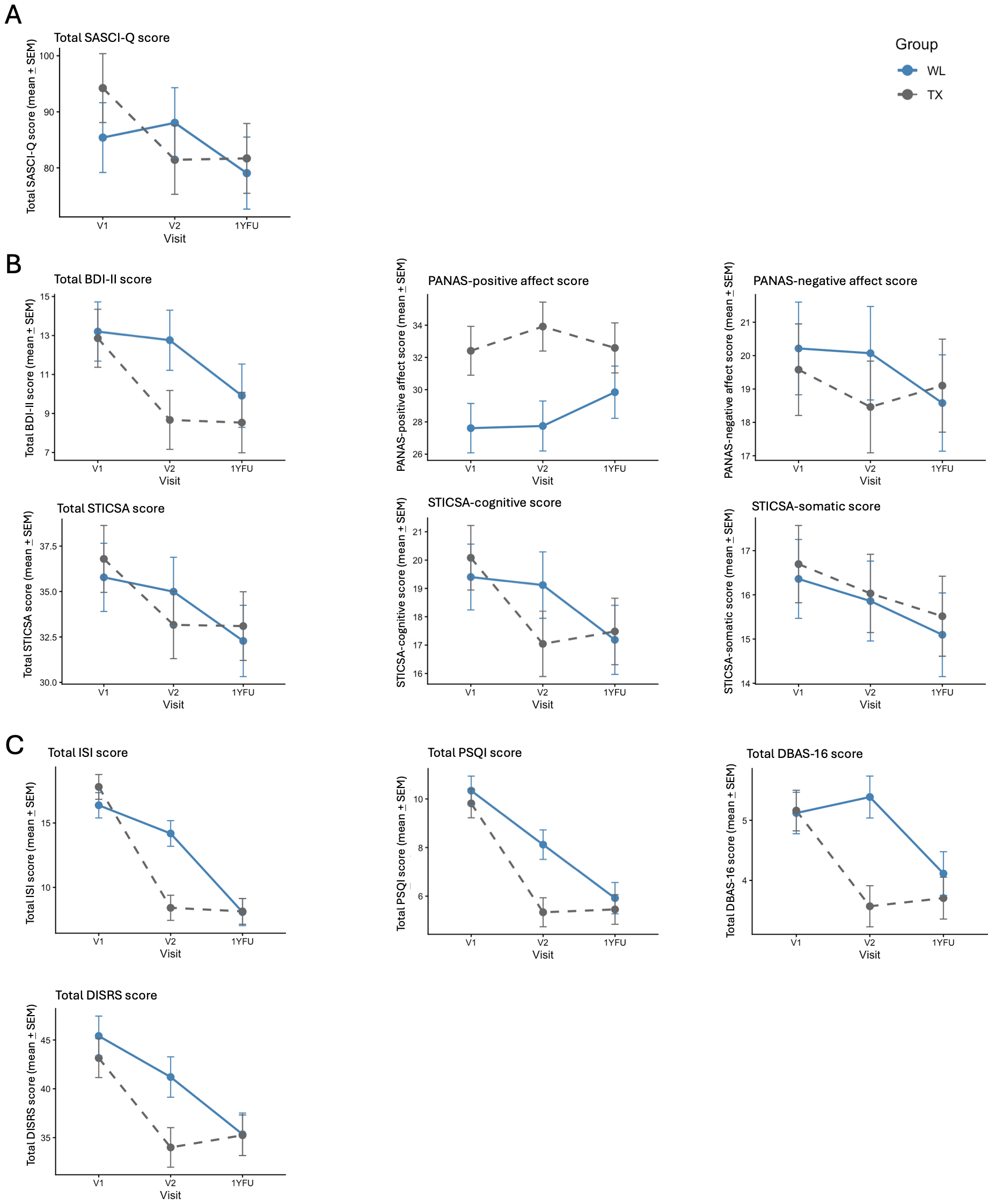

#### **Figure S7.** Effects of CBTi on total ISI scores

The TX group is shown in grey with a dashed line, whereas the WL group is shown in blue with a solid line. Error bars represent standard error of the mean (SEM). Asterisks represent statistical significance (*p*): * <.05. ISI: Insomnia Severity Index; TX: Immediate CBTi group; WL: Wait-list control group.

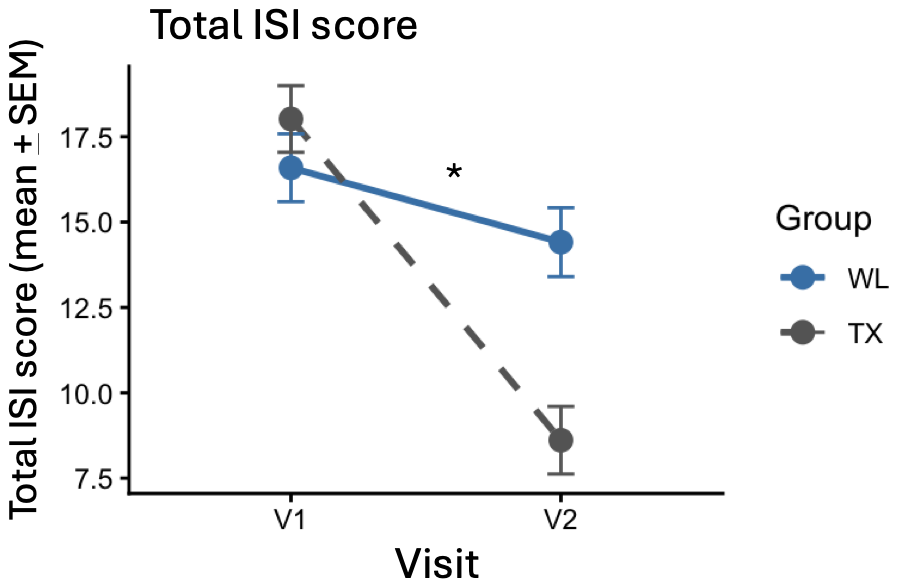

#### **Table S9.** Effects of CBTi on cognitive performance on the Sternberg working memory, emotion reactivity, and associative memory tasks (CBTi responders vs. non-responders)

| Outcome | Group effect | | |  | Visit effect | | |  | Group*Visit interaction | | |
| --- | --- | --- | --- | --- | --- | --- | --- | --- | --- | --- | --- |
|  | *F (df)* | *p* | $\beta$ [95% CI] |  | *F (df)* | *p* | $\beta$ [95% CI] |  | *F (df)* | *p* | $\beta$ [95% CI] |
| **Sternberg working memory** | | | | | | | | | | | |
| Accuracy (% correct) | 0.00 (21.00) | .99 | 0.01 [-0.34, 0.35] |  | 1.44 (23.00) | .48 | -0.16 [-0.46, 0.15] |  | 1.98 (23.00) | .44 | 0.19 [-0.13, 0.51] |
| **Emotion reactivity** | | | | | | | | | | | |
| Valence rating for neutral images | 0.02 (21.00) | .90 | -0.03 [-0.41, 0.36] |  | 1.23 (23.00) | .28 | 0.11 [-0.09, 0.30] |  | 0.68 (23.00) | .90 | 0.08 [-0.15, 0.31] |
| Valence rating for negative images | 0.02 (21.00) | .90 | -0.03 [-0.40, 0.36] |  | 1.95 (23.00) | .26 | 0.15 [-0.09, 0.38] |  | 0.15 (23.00) | .90 | 0.04 [-0.19, 0.27] |
| Valence rating for sleep images | 0.98 (21.00) | .90 | 0.20 [-0.29, 0.69] |  | 4.52 (23.00) | .13 | 0.18 [-0.03, 0.39] |  | 0.02 (23.00) | .90 | -0.01 [-0.18, 0.16] |
| **Face-name associative memory** | | | | | | | | | | | |
| Number of hits | 0.19 (10.00) | .98 | 0.13 [-0.54, 0.80] |  | 0.12 (12.00) | .80 | 0.03 [-0.15, 0.21] |  | 0.03 (12.00) | .92 | 0.02 [-0.17, 0.20] |
| Accuracy (Hit+CR - Miss+FA) | 0.00 (10.00) | .98 | -0.01 [-0.62, 0.60] |  | 2.20 (12.00) | .41 | 0.12 [-0.09, 0.33] |  | 0.86 (12.00) | .84 | 0.08 [-0.13, 0.28] |
| Recognition (Hit-FA) | 0.01 (10.00) | .98 | -0.02 [-0.65, 0.60] |  | 0.97 (12.00) | .65 | 0.08 [-0.12, 0.28] |  | 1.09 (12.00) | .84 | 0.09 [-0.13, 0.30] |

*Note*. The reported *p*-values were adjusted for multiple comparisons using the Benjamini-Hochberg False Discovery Rate procedure. Standardized beta coefficients ($\beta$) are reported with 95% confidence intervals ([95% CI]). CR: Correct Rejection; FA: False Alarm.

#### **Table S10.** Brain activity during the Sternberg working memory, emotion reactivity, and face-name associative memory tasks at V1 and V2 (CBTi responders vs. non-responders)

|  |  |  |  |  |  | Cluster-level | |  | Peak-level | | |
| --- | --- | --- | --- | --- | --- | --- | --- | --- | --- | --- | --- |
| Visit | Group | Contrast | Brain region  (BA) | Coordinates  (x, y, z) |  | Cluster size (voxels) | *p_FWE-corrected_* |  | *t* | *z_E_* | *p_FWE-corrected_* |
|  | **Sternberg working memory task** | | | | | | | | | | |
| V1 | Responder | Correct encoding |  |  |  |  |  |  |  |  |  |
|  |  |  | Left anterior insula (BA13) | (-34, 20, 2) |  | 224 | .000 |  | 8.31 | 6.46 | .000 |
|  |  |  | Right anterior insula (BA13) | (32, 24, -2) |  | 90 | .000 |  | 6.68 | 5.55 | .001 |
|  |  |  | Left precentral gyrus (BA6) | (-38, -4, 50) |  | 71 | .000 |  | 6.10 | 5.19 | .003 |
|  |  |  | Right middle frontal gyrus (BA46) | (40, 34, 18) |  | 15 | .010 |  | 5.64 | 4.89 | .012 |
|  |  |  | Right lingual gyrus (BA18) | (12, -74, 2) |  | 35 | .003 |  | 5.64 | 4.89 | .012 |
|  |  |  | Left medial frontal gyrus (BA6) | (-2, 20, 46) |  | 39 | .002 |  | 5.44 | 4.75 | .021 |
|  |  |  | Left lingual gyrus (BA18) | (-10, -72, 2) |  | 1 | .038 |  | 5.14 | 4.54 | .049 |
| V1 | Non-responder | Correct encoding |  |  |  |  |  |  |  |  |  |
|  |  |  | Right anterior insula (BA13) | (32, 26, -2) |  | 39 | .002 |  | 6.27 | 5.30 | .002 |
|  |  |  | Right lingual gyrus (BA18) | (8, -74, 0) |  | 25 | .005 |  | 6.10 | 5.19 | .003 |
|  |  |  | Left anterior insula (BA13) | (-32, 22, 2) |  | 31 | .004 |  | 5.91 | 5.07 | .005 |
| V2 | Responder | Correct encoding |  |  |  |  |  |  |  |  |  |
|  |  |  | Left anterior insula (BA13) | (-34, 20, 4) |  | 150 | .000 |  | 7.02 | 5.76 | .000 |
|  |  |  | Right anterior insula (BA13) | (32, 24, -4) |  | 39 | .002 |  | 5.93 | 5.08 | .005 |
| V2 | Non-responder | Correct encoding |  |  |  |  |  |  |  |  |  |
|  |  |  | Right anterior insula (BA13) | (32, 26, 0) |  | 7 | .019 |  | 5.45 | 4.76 | .020 |
|  |  |  | Left anterior insula (BA13) | (-32, 20, 4) |  | 2 | .033 |  | 5.25 | 4.62 | .036 |
|  | **Emotion reactivity task** | | | | | | | | | | |
| V1 | Responder | Sleep > Neutral | No suprathreshold clusters | | | | | | | | |
| V1 | Non-responder | Sleep > Neutral |  |  |  |  |  |  |  |  |  |
|  |  |  | Left middle temporal gyrus (BA39) | (-40, -64, 32) |  | 40 | .004 |  | 5.58 | 4.85 | .011 |
|  |  |  | Left angular gyrus (BA39) | (-42, -74, 38) |  | 1 | .040 |  | 5.14 | 4.54 | .040 |
| V2 | Responder | Sleep > Neutral | No suprathreshold clusters | | | | | | | | |
| V2 | Non-responder | Sleep > Neutral | No suprathreshold clusters | | | | | | | | |
| V1 | Responder | Negative > Neutral | No suprathreshold clusters | | | | | | | | |
| V1 | Non-responder | Negative > Neutral | No suprathreshold clusters | | | | | | | | |
| V2 | Responder | Negative > Neutral |  |  |  |  |  |  |  |  |  |
|  |  |  | Right inferior occipital gyrus (BA18) | (42, -86, 0) |  | 2 | .034 |  | 5.27 | 4.63 | .031 |
|  |  |  | Right cerebellum exterior | (30, -74, -14) |  | 2 | .034 |  | 5.19 | 4.58 | .039 |
| V2 | Non-responder | Negative > Neutral |  |  |  |  |  |  |  |  |  |
|  |  |  | Right cerebellum exterior | (28, -76, -14) |  | 31 | .004 |  | 6.34 | 5.35 | .001 |
|  |  |  | Left inferior frontal gyrus (BA45) | (-58, 26, 10) |  | 2 | .034 |  | 5.17 | 4.57 | .040 |
|  | **Face-name associative memory task** | | | | | | | | | | |
| V1 | Responder | Hit + Miss > Control | No suprathreshold clusters | | | | | | | | |
| V1 | Non-responder | Hit + Miss > Control | No suprathreshold clusters | | | | | | | | |
| V2 | Responder | Hit + Miss > Control | No suprathreshold clusters | | | | | | | | |
| V2 | Non-responder | Hit + Miss > Control |  |  |  |  |  |  |  |  |  |
|  |  |  | Right postcentral gyrus (BA40) | (68, -24, 22) |  | 1 | .036 |  | 6.35 | 4.74 | .030 |
| V1 | Responder | Hit > Control | No suprathreshold clusters | | | | | | | | |
| V1 | Non-responder | Hit > Control | No suprathreshold clusters | | | | | | | | |
| V2 | Responder | Hit > Control | No suprathreshold clusters | | | | | | | | |
| V2 | Non-responder | Hit > Control |  |  |  |  |  |  |  |  |  |
|  |  |  | Right inferior parietal lobule (BA40) | (68, -26, 24) |  | 3 | .027 |  | 6.86 | 4.97 | .011 |

*Note*. Results are presented with *p_FWE-corrected_* set to .05 and extent threshold (voxels) set to 0. Coordinates are presented in mm.

#### **Table S11.** Results for the effects of CBTi on polysomnography-derived sleep measures

| Outcome | Group effect | | |  | Visit effect | | |  | Group*Visit interaction | | |
| --- | --- | --- | --- | --- | --- | --- | --- | --- | --- | --- | --- |
|  | *F (df)* | *p* | $\beta$ [95% CI] |  | *F (df)* | *p* | $\beta$ [95% CI] |  | *F (df)* | *p* | $\beta$ [95% CI] |
| **Sleep** | | | | | | | | | | | |
| TST | 0.54 (50.71) | .56 | -0.08 [-0.15, 0.32] |  | 0.09 (51.84) | .85 | -0.02 [-0.17, 0.13] |  | 0.26 (51.83) | .61 | -0.04 [-0.19, 0.11] |
| SOL | 1.07 (52.61) | .56 | -0.11 [-0.36, 0.13] |  | 0.10 (53.90) | .85 | -0.03 [-0.19, 0.13] |  | 3.82 (53.90) | .07 | -0.15 [-0.32, 0.01] |
| WASO | 0.34 (52.33) | .56 | -0.07 [-0.30, 0.16] |  | 0.04 (53.27) | .85 | 0.01 [-0.12, 0.15] |  | 6.31 (53.26) | .06 | -0.17 [-0.34, 0.00] |
| SE | 0.56 (52.28) | .56 | 0.09 [-0.16, 0.33] |  | 0.31 (53.18) | .85 | -0.04 [-0.18, 0.11] |  | 4.22 (53.18] | .07 | 0.14 [-0.01, 0.29] |

*Note*. The reported *p*-values were adjusted for multiple comparisons using the Benjamini-Hochberg False Discovery Rate procedure. Standardized beta coefficients ($\beta$) are reported with 95% confidence intervals ([95% CI]). SE: Sleep Efficiency; SOL: Sleep Onset Latency; TST: Total Sleep Time; WASO: Wake After Sleep Onset.
